# Under-recognized or Under-reported? A Global Bibliometric Analysis (1994-2025) of Neurological Assessment in Pediatric Intensive Care Units

**DOI:** 10.64898/2026.03.11.26348139

**Authors:** José C. M. Rodrigues, Brenda C. da Conceição, Lucas Villar P. S. Pantoja, Kissila M. Machado-Ferraro, Mary L. F. Maia, Fábio J. C. Souza-Junior, Rafael Rodrigues Lima, Rodrigo A. Cunha, Roberta Esteves Vieira de Castro, Fernando Bezerra, Luanna M. P. Fernandes, Enéas A. Fontes-Junior, Cristiane S. F. Maia

**Author notes:** These authors contributed equally to the work (José C. M. Rodrigues, Brenda C. da Conceição, Lucas Villar P. S. Pantoja). Corresponding author: Cristiane Socorro Ferraz Maia;.

## Abstract

Critically ill children admitted to pediatric intensive care units (PICUs) face significant neurological risks, but it remains unclear whether neurological assessments are adequately integrated into clinical routines. We aimed to evaluate the global scientific landscape regarding neurological examination in PICU, combining bibliometric analysis with clinical-guided critical analysis. A comprehensive search was conducted on the Web of Science Core Collection (WoS-CC) using terms related to pediatric critical care and neurological evaluation. Eligible publications were original research articles approaching neurological assessment during PICU stay. Bibliometric indicators, science mapping, and study design profiling were analyzed. In a separate, clinically guided interpretative layer, reporting patterns related to timing, tools, and strategies of neurological assessment were synthesized. From 359 records, 128 articles met inclusion criteria. The United States accounted for over half of the publications, while most studies employed retrospective designs and focused on traumatic brain injury or cardiac arrest. Despite the relevance of clinical neurological examination – especially using the Glasgow Coma Scale (GCS) and Pediatric Cerebral Performance Category (PCPC), advanced neuromonitoring tools, e.g., EEG, intracranial pressure monitoring, and biomarkers were inconsistently applied. Notably, neurological evaluations were often underreported at admission and discharge and rarely extended to non-neurological PICU conditions. Our findings reveal a critical gap between the neurological vulnerability of PICU patients, and the limited, inconsistent assessment strategies reported in the literature. Expanding structured neurological evaluation to all critically ill children, not only those with overt neurological diagnoses, seems essential to promote brain health and long-term recovery.

## 1 Introduction

Children admitted to intensive care units represent a particularly vulnerable population that often face significant neurological sequelae^1^. Epidemiological data indicate that up to 90% of deaths among previously healthy children admitted to pediatric intensive care units (PICUs) in the United States are attributed to brain complications^1^. Moreover, the morphophysiological changes that occur throughout childhood are essential for proper neurodevelopment and may be closely linked to the emergence of various neuropsychiatric disorders^2^. The PICU environment *per se* encompasses multiple harmful factors – including continuous exposure to noise and light stimuli, invasive procedures, sedative and analgesic medications, and underlying comorbidities. These negative persistent noxious stimulations may potentially disrupt neural networks in the developing brain and contribute to long-term neurological impairments^3^.

It is well established that PICUs require specialized care and precise techniques, delivered by a qualified multidisciplinary team, encompassing all aspects of care from admission to discharge^4,5^. However, it remains unclear whether the assessment of neurological conditions and alterations is adequately integrated into routine clinical practice. Brown et al.^6^ reported that neurological injuries account for 16.2% of PICU admissions in the United States and Europe and identified key tools for neonatal neurological assessment. Despite notable contributions from Brown et al.^6^ and Hovart et al.^1^ to address this topic, a significant gap in the literature persists regarding studies that explore the management of critically ill pediatric patients with neurological conditions throughout the PICU stay.

Our central hypothesis suggests that the neurological assessment of critically ill pediatric patients remains underemphasized in current clinical research. To investigate this issue, we conducted a bibliometric-type analysis. Using bibliometric techniques, we analyzed publication trends, contributing authors, global distribution of studies, keywords, citation patterns, and the types of studies addressing this topic. Additionally, we incorporated a critical appraisal to map clinical practices, emerging trends, and key findings, with particular emphasis on aspects related to neurological evaluation and management.

## 2 Materials and methods

We performed a global bibliometric analysis, following the BIBLIO guidelines (Guideline for Reporting Bibliometric Reviews of the Biomedical Literature) and an approach previously employed by our research group to tackle a different research question^7–10^. We examined publications about neurological evaluation within pediatric critical care using the Web of Science Core Collection (WoS-CC), selected due to its compatibility with standardized bibliometric tools and extensive coverage of research articles, covering the period from 1945 to the present day. A search was conducted on June 10, 2025, combining terms related to pediatric critical care (e.g., PICU, critically ill children) with neurological assessment terms (e.g., neurological examinations, neurodevelopmental outcomes) (Figure 1).

**Figure 1.**
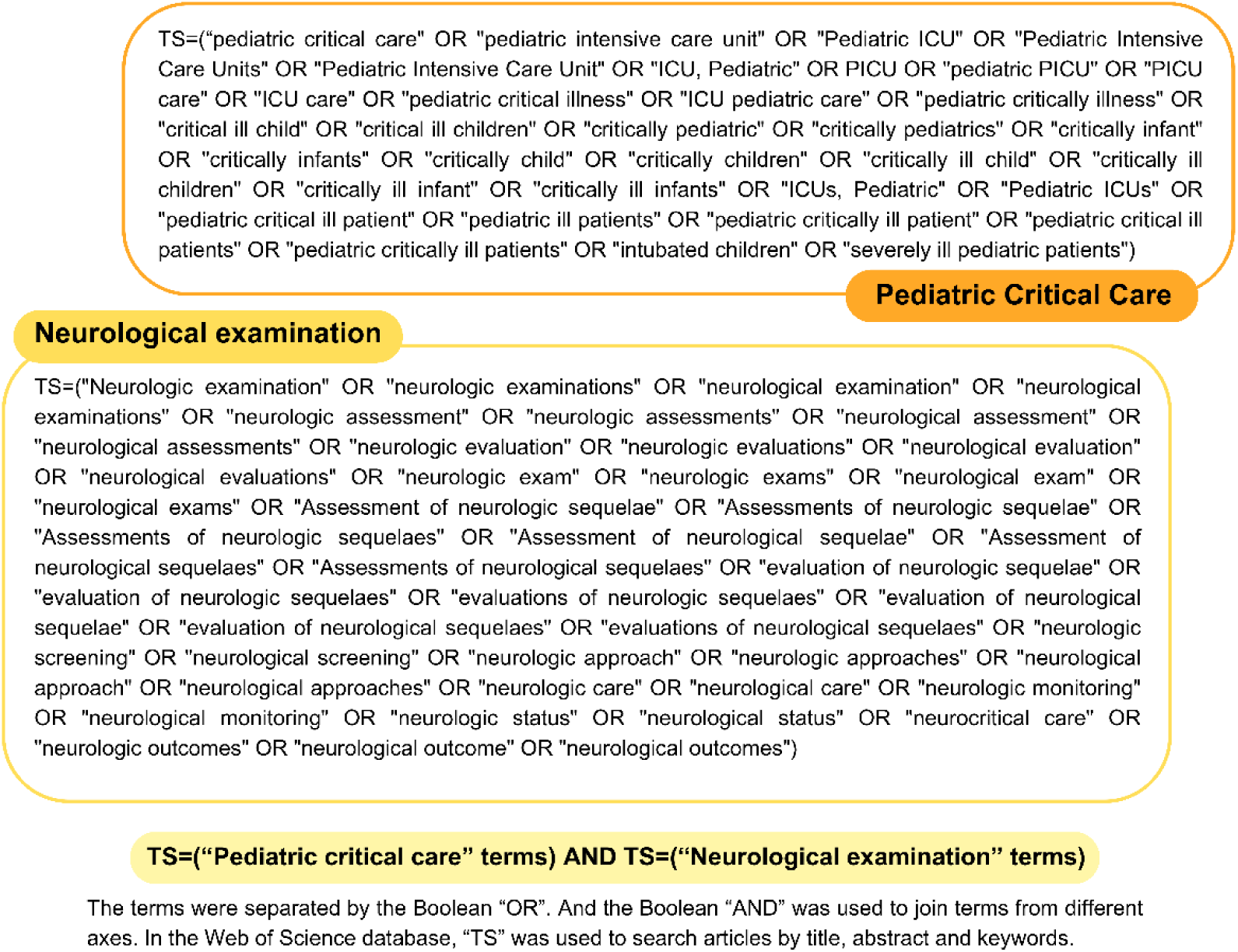
Search strategy.

### 2.1 Inclusion and exclusion criteria

The primary inclusion criterion was the explicit focus on neurological evaluation in critically ill children admitted to the PICU. Eligible studies were original research articles that reported neurological assessments conducted during PICU stay or spanning both PICU and post-PICU periods, regardless of publication language. Only peer-reviewed studies were assessed to ensure methodological rigor.

Exclusion criteria were: (i) review articles, editorials, letters, and conference papers; (ii) publications not directly related to neurological evaluation in PICU; (iii) studies that assessed neurological status exclusively in the post-PICU period; and (iv) survey-based studies (e.g., questionnaires distributed to hospitals or professionals). While surveys provide valuable insights into organizational structures of pediatric neurocritical care, they do not generate patient-level clinical data, which was the central focus of this analysis.

### 2.2 Data selection

To minimize bias during selection process, two independent reviewers screened all retrieved records for eligibility. In cases of disagreement, a senior investigator acted as an arbitrator to reach a consensus on final inclusion. The bibliographic data was extracted directly from the WoS-CC and exported in plain text format, which is compatible with bibliometric analysis tools. Data processing and visualization were performed using VOSviewer software (version 1.6.16; Leiden University, The Netherlands).

### 2.3 Bibliometric Procedures

The comprehensive metadata available in WoS-CC enabled analyses of performance metrics, such as citation trends and author/institution impact, as well as science mapping techniques, including co-authorship and keyword co-occurrence analyses performed via VOSviewer. This study adhered to standard bibliometric principles. Lotka’s Law was applied to analyze author productivity and collaboration patterns; Zipf’s Law was used to assess keyword frequency; and Bradford’s Law helped identify the most influential journals, based on the Journal Citation Reports (JCR) 2024 Edition. Data extracted regarding publication volume, authorship, keywords, geographic distribution of research output across countries and journals of publication were used to generate descriptive statistics and visual representations. Descriptive analyses were performed using Microsoft Excel (365 version), network visualizations were developed with VOSviewer software (version 1.6.16), and geographic mapping of countries was created using MapChart (mapchart.net, accessed on June 16, 2025).

### 2.4 Clinical-guided Critical Analysis

To identify key findings and gaps in neurological assessment practices, we conducted a clinically guided critical appraisal. Information on clinical context, and neurological management strategies was extracted and categorized into three time points: admission, PICU stay, and discharge. The key findings of each article were highlighted and summarized. Given the heterogeneity of study types, we adopted the original classifications provided by the authors to define the study design.

An overview of the methodological workflow, highlighting the sequential bibliometric procedures and the subsequent clinical-guided critical analysis, is summarized in Figure 2.

**Figure 2.**
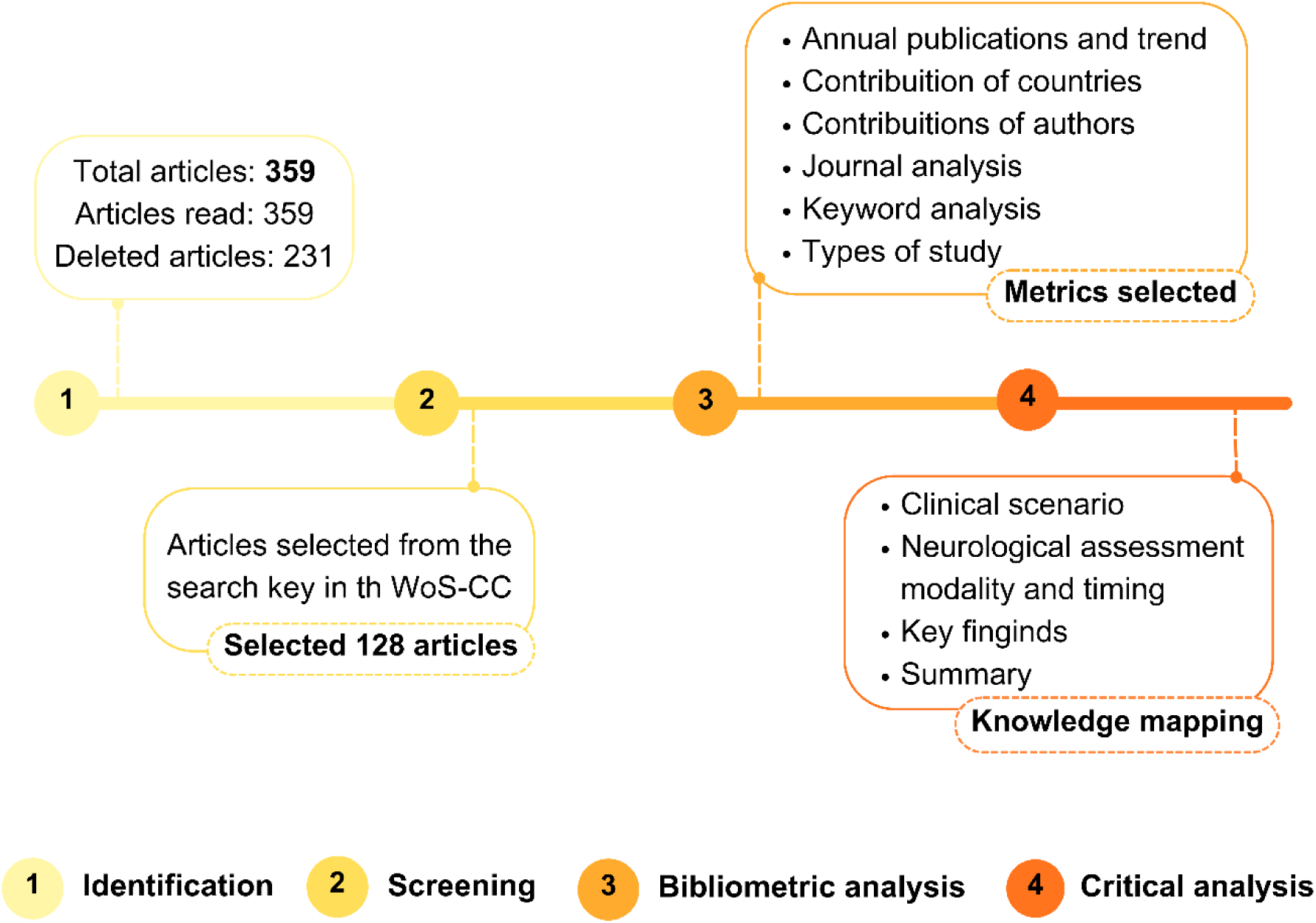
Methodological Strategy Flowchart.

## 3 Results

### 3.1 Article presentation

From an initial pool of 359 documents retrieved from the WoS-CC platform, 128 articles met the inclusion criteria for this bibliometric analysis, which reflects the limited global literature on this topic. The selected publications span a 31-year period, from 1994 to 2025. The earliest study, published in 1994, investigated the utility of serial neurological examinations as predictive tools for post-drowning resuscitation outcomes^11^. In contrast, the most recent article, published in 2025, focused on the use of serum-based brain injury biomarkers to support neurological evaluation in critically ill pediatric patients^1^. Citation counts among the studies included ranged from 0 to 281. The most cited publication was a comprehensive observational study that evaluated neurological assessment techniques, with a particular focus on patterns of neurological decline associated with seizure burden^12^.

### 3.2 Global Landscape of Scientific Output

Scientific literature on critically ill children and neurological examinations was widely distributed across nearly every continent (Figure 3A). The United States dominated global output, accounting for more than half of the publications (n = 70), followed by France (n = 9), Turkey (n = 9), India (n = 8), and Spain (n = 5). Although represented to a lesser extent, the topic’s reach extended to countries such as the United Arab Emirates, Greece, Singapore, Costa Rica, and Taiwan, each contributing one article, underscoring the global recognition of this issue (Figure 3B).

**Figure 3.**
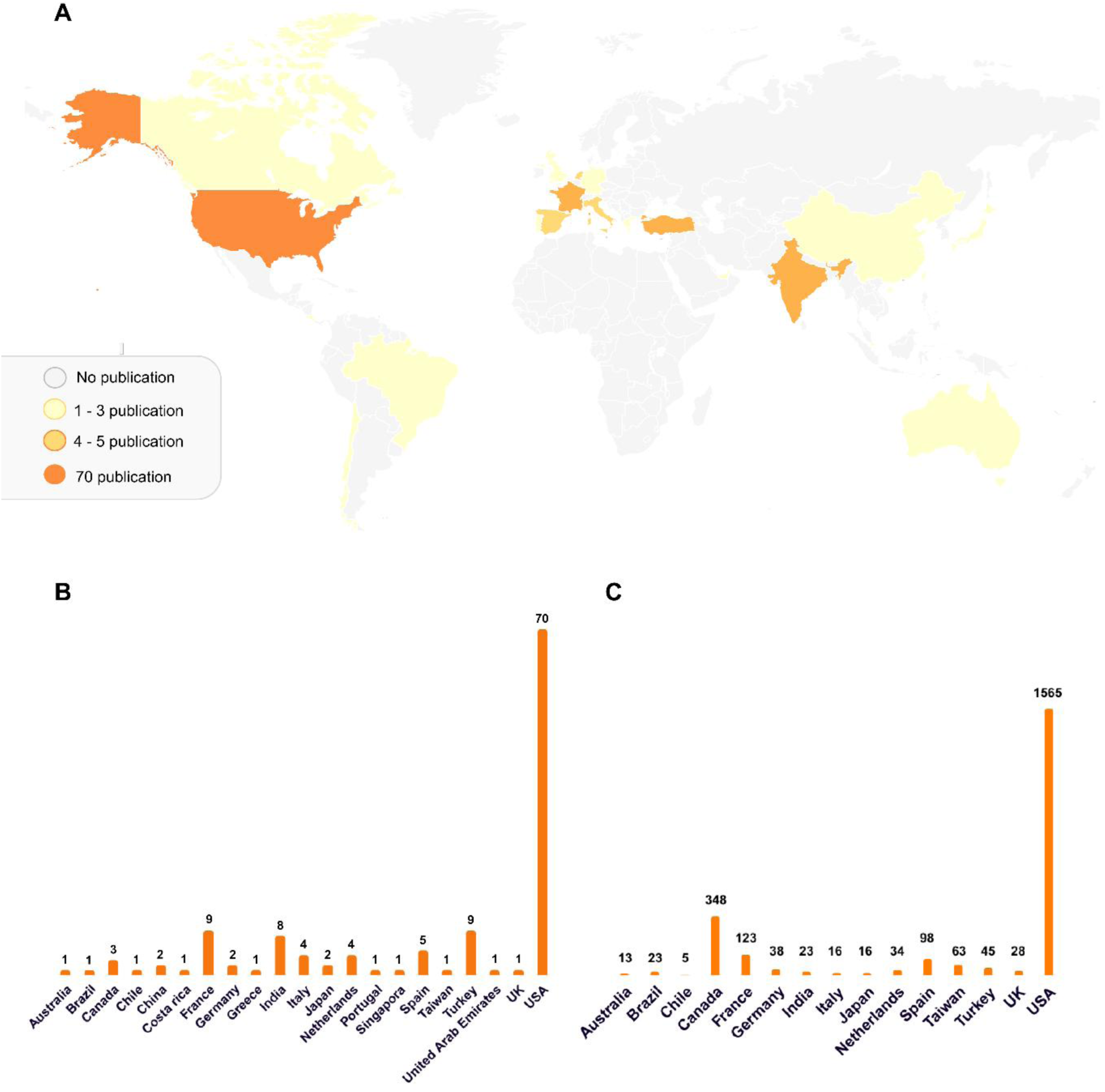
Geographic distribution of all selected articles (A) with number of publications (B) and citations (C).

Regarding citations (considering countries with ≥ 5), the United States also led (n = 1,965), followed by Canada (n = 348), France (n = 123), and Spain (n = 98). Despite publishing more articles, Turkey and India received fewer citations (n = 45 and n = 23, respectively). Notably, Taiwan (n = 63), the United Kingdom (n = 28), and Brazil (n = 23) achieved considerable citation counts despite having only a single publication each (Figure 3C).

### 3.3 Publication Channels: A Journal-Level Analysis

A total of 62 distinct journals contributed to the body of literature on neurological examination in pediatric critical care (Table 2). According to Bradford’s Law, a small core of journals should account for a large proportion of publications. In our analysis, this core was represented primarily by *Pediatric Critical Care Medicine* (n = 20), *Neurocritical Care* (n = 10), and *Resuscitation* (n = 7), which together they accounted for 37 of the 128 articles. These journals also demonstrated high citation counts, particularly *Pediatric Critical Care Medicine* (556 citations) and *Resuscitation* (269 citations), reinforcing their influence in the field. A second line of journals, including *Critical Care Medicine* (n = 6), *Pediatric Neurology* (n = 6), *Frontiers in Pediatrics* (n = 5), and *Journal of Neurosurgery: Pediatrics* (n = 4), also exhibited notable productivity. The remaining 52 journals each contributed only one or two publications.

**Table 1.**
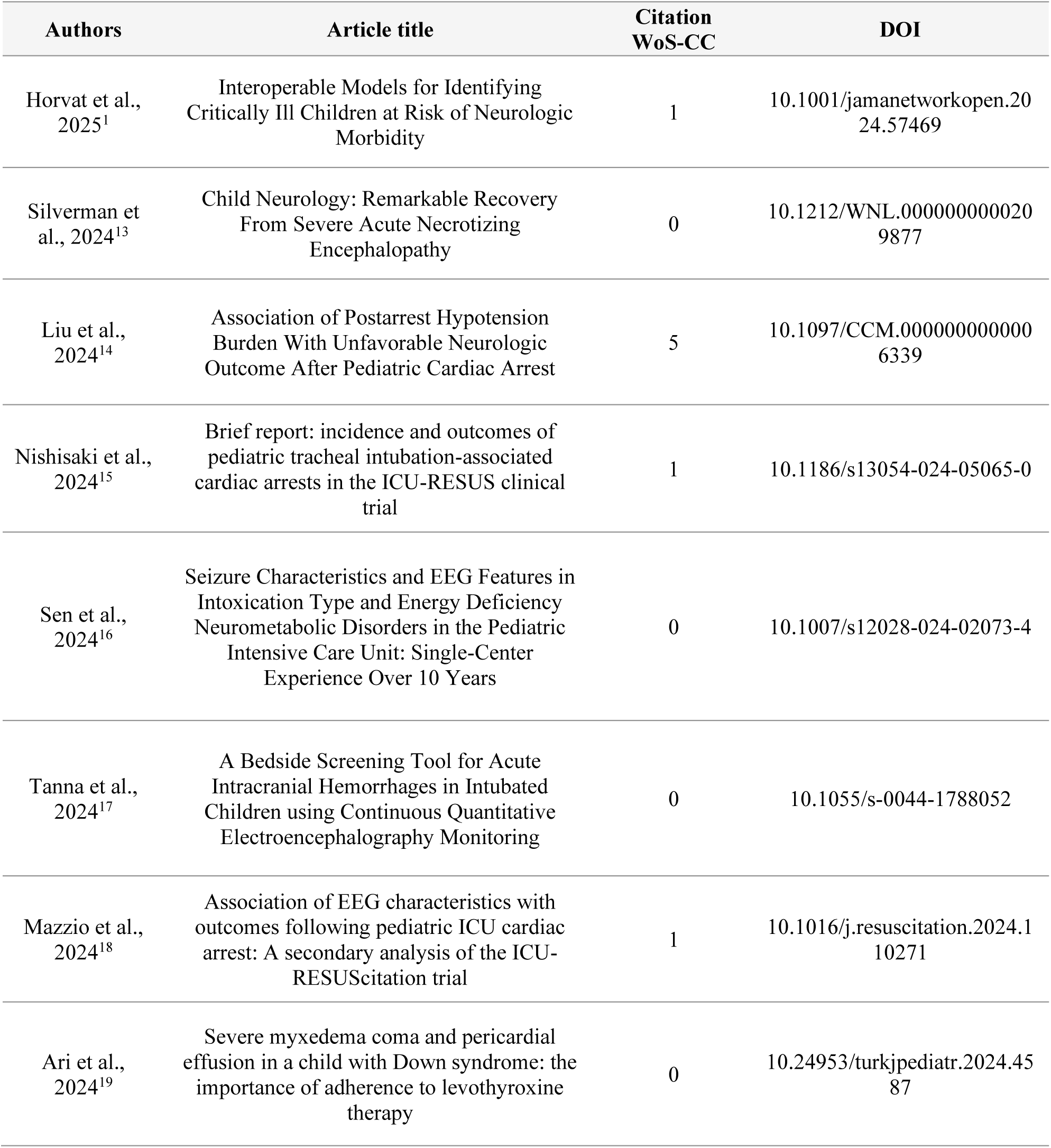
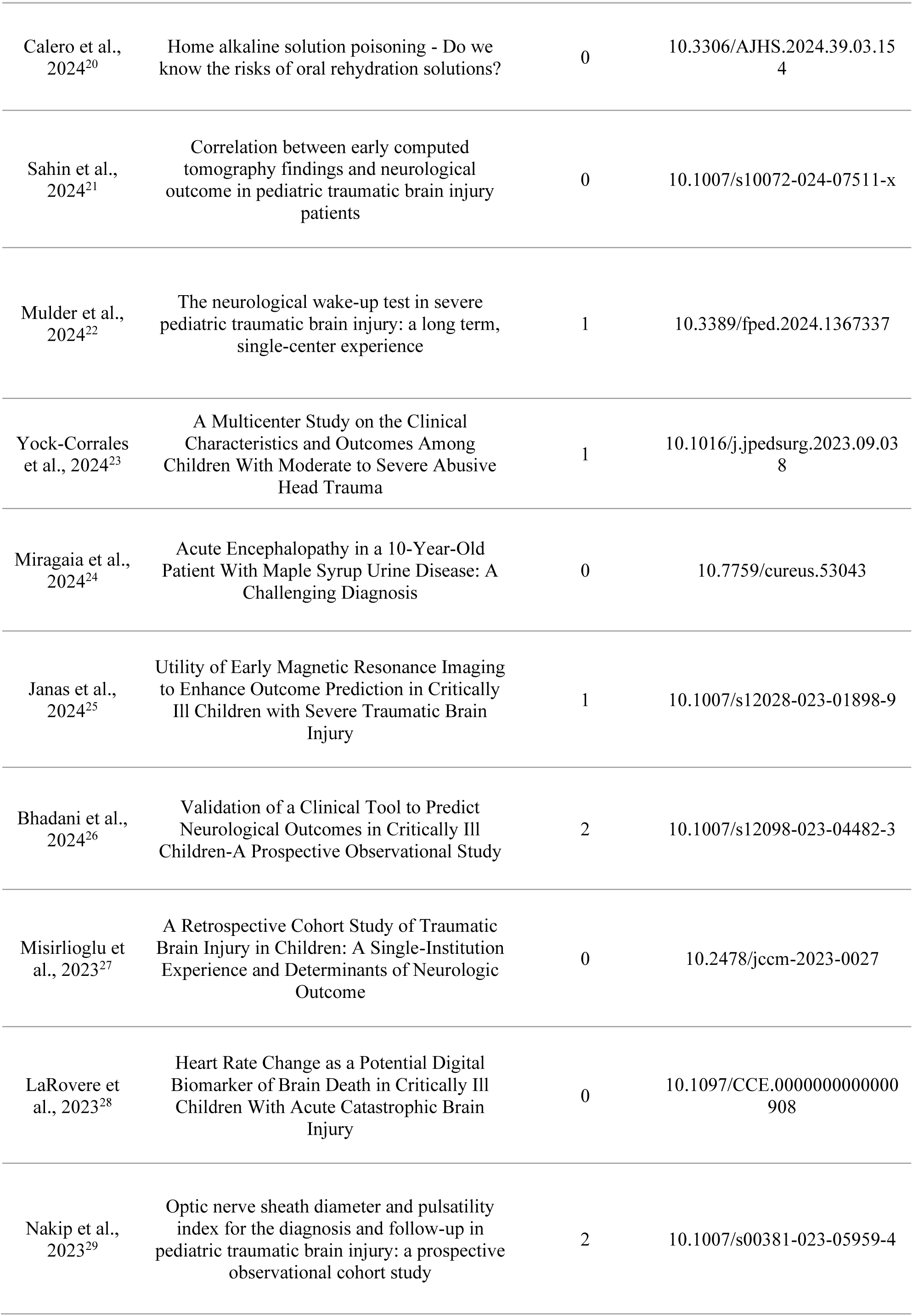
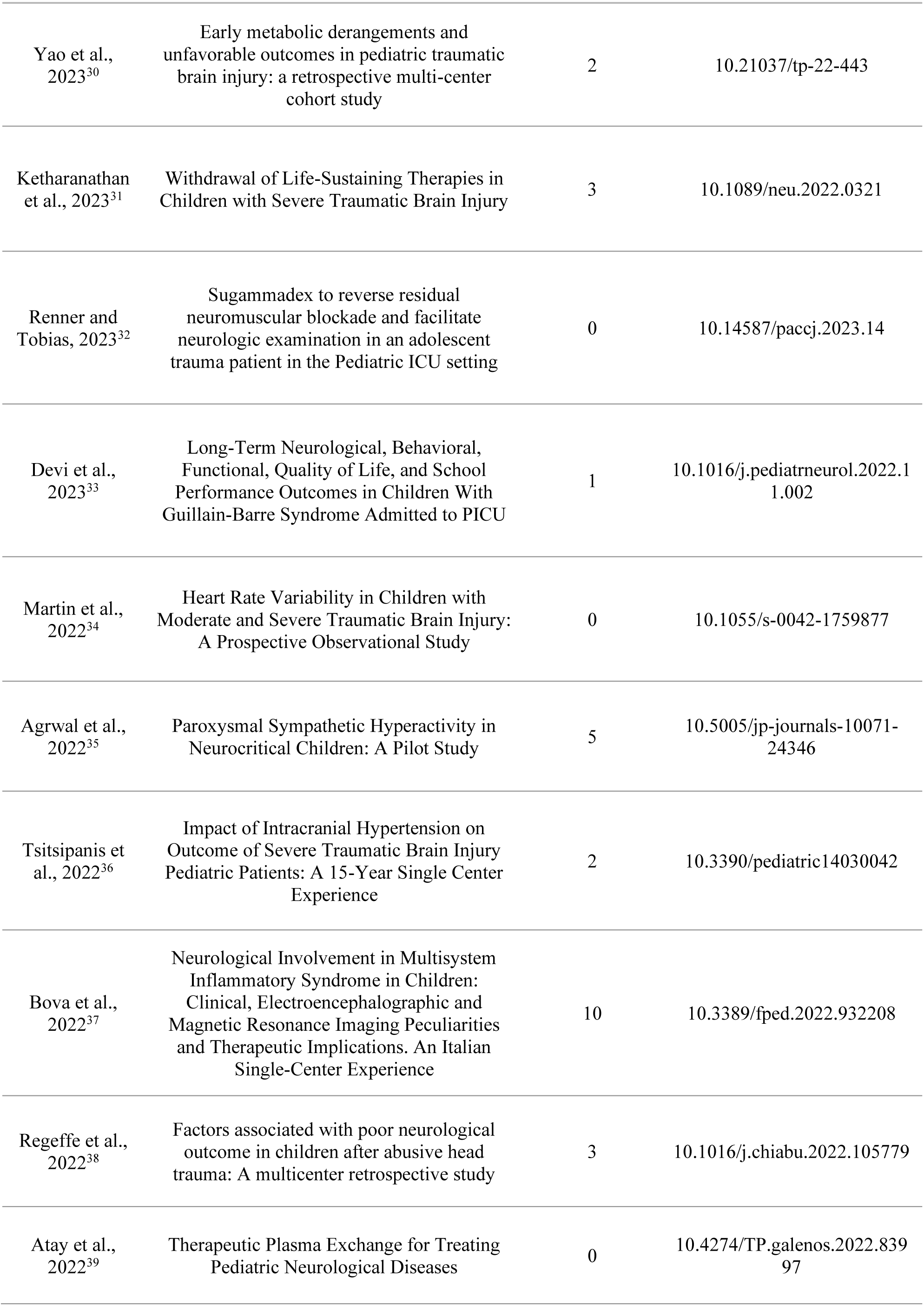
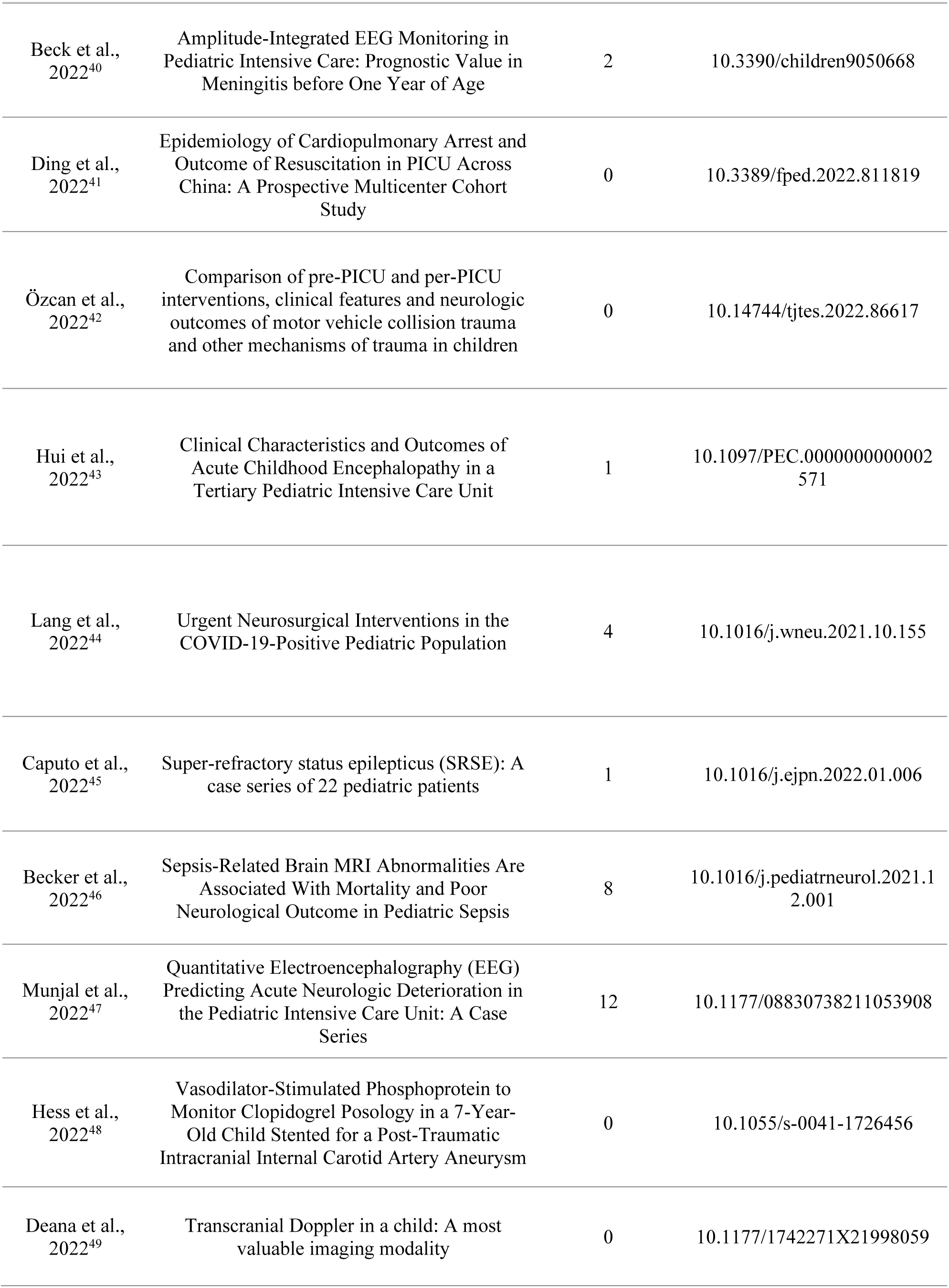
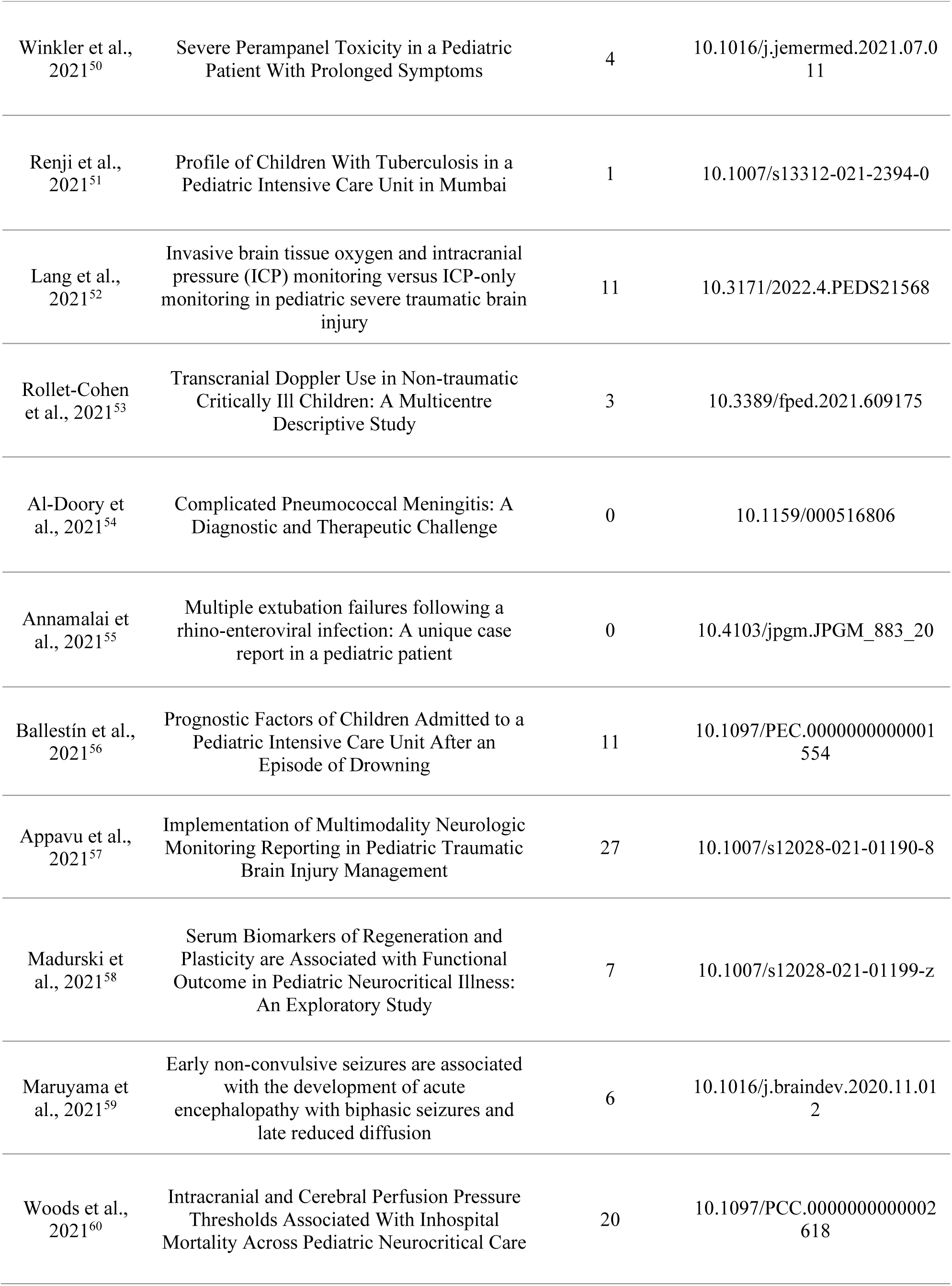
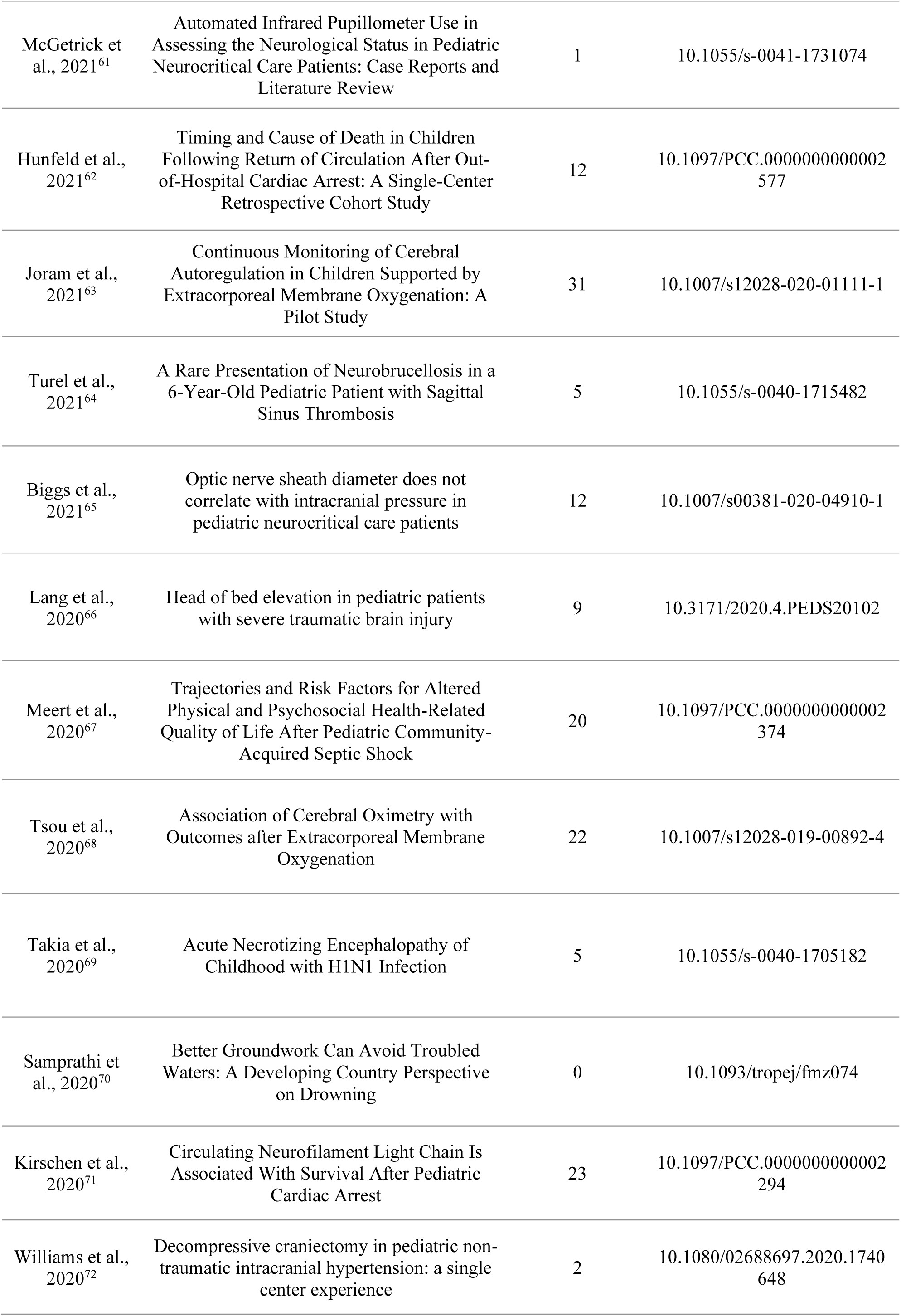
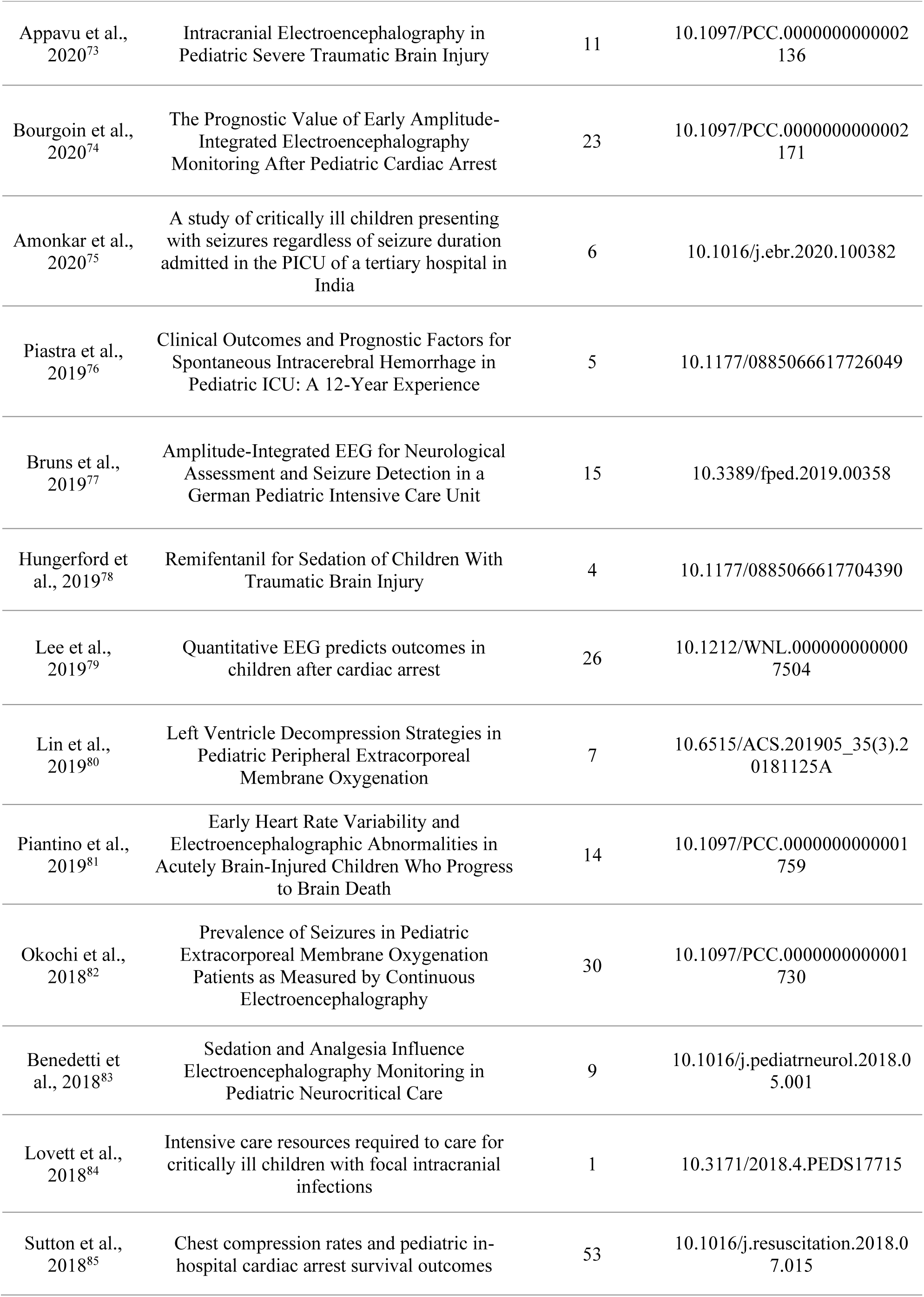
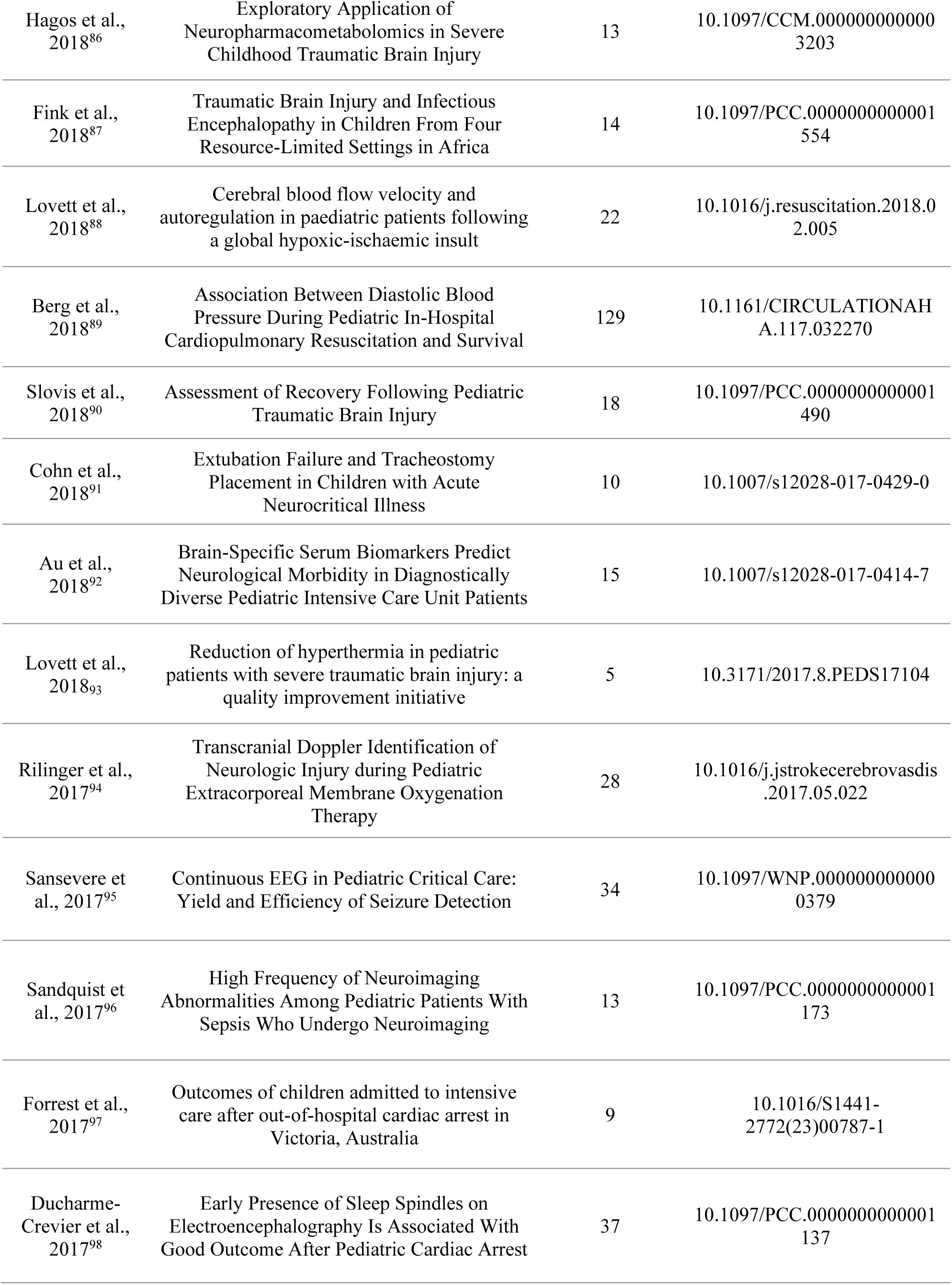
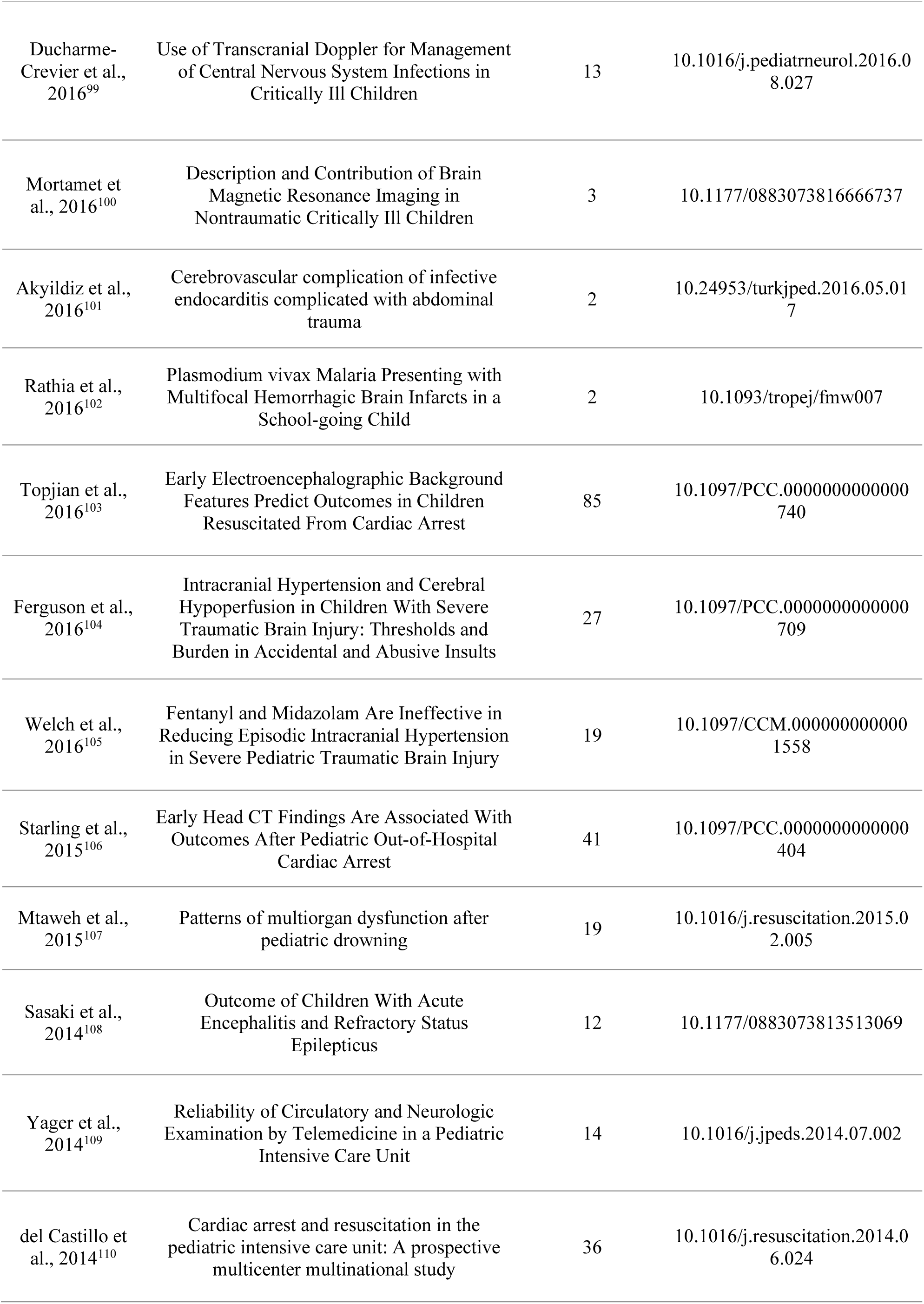
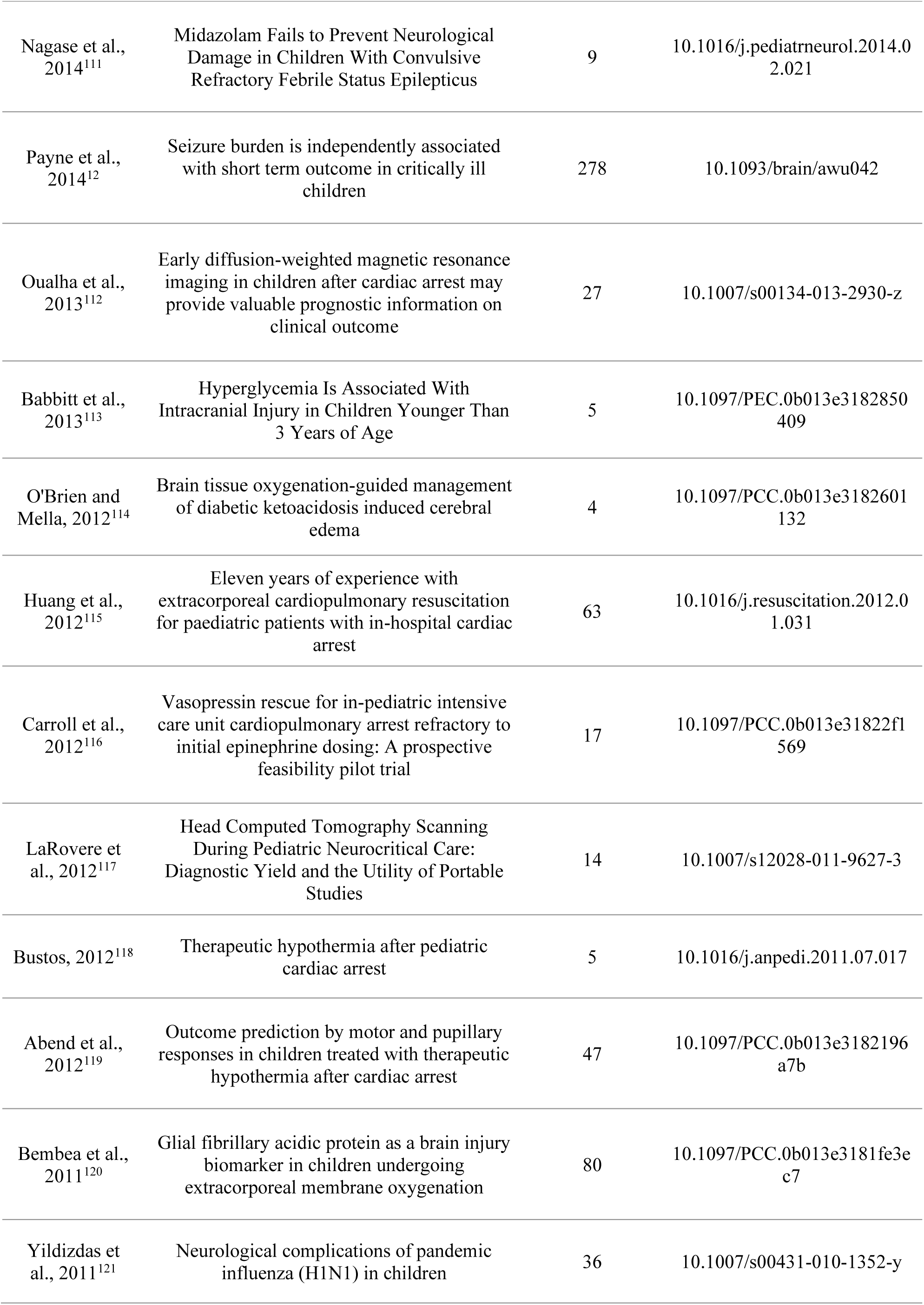
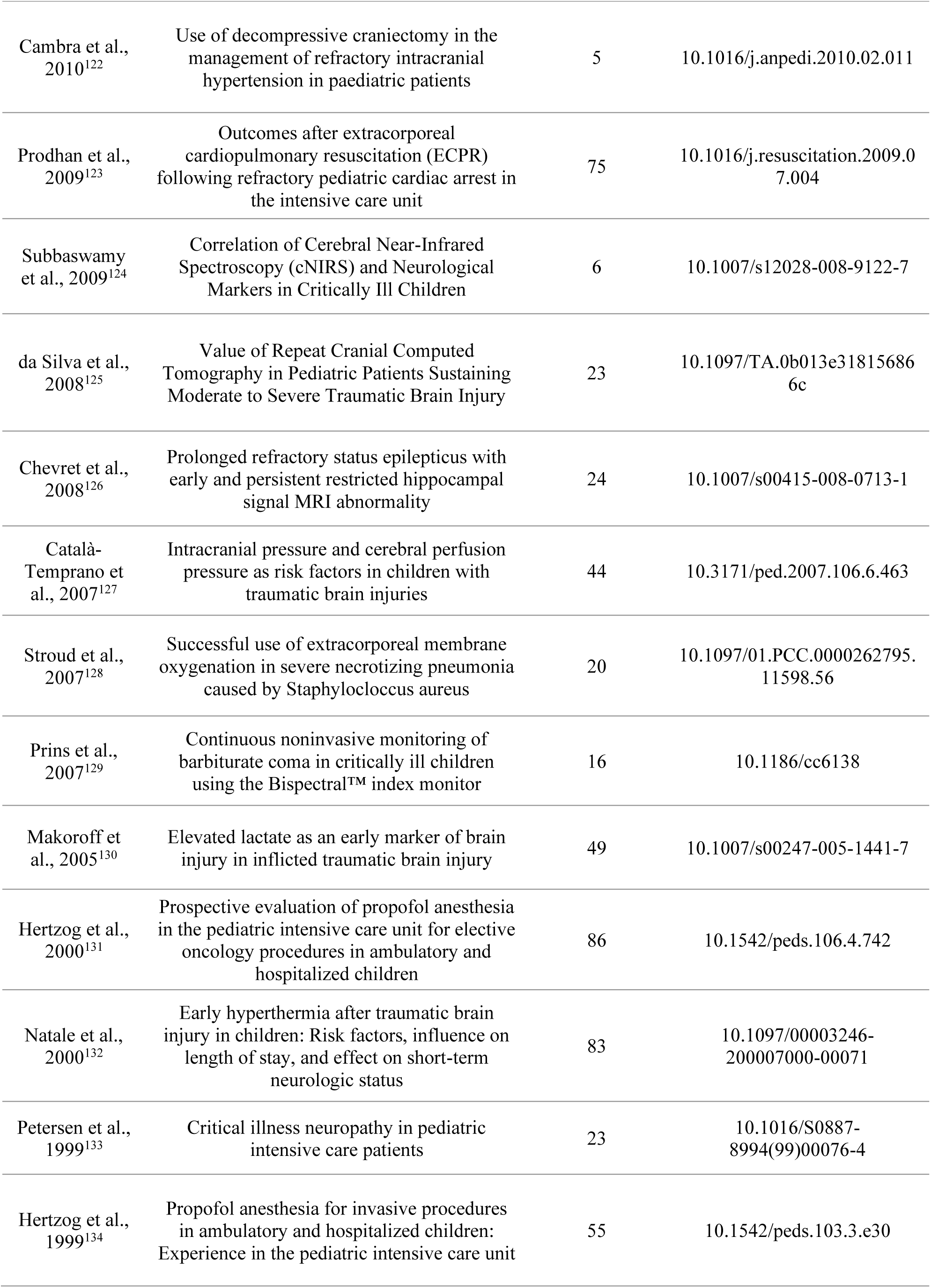
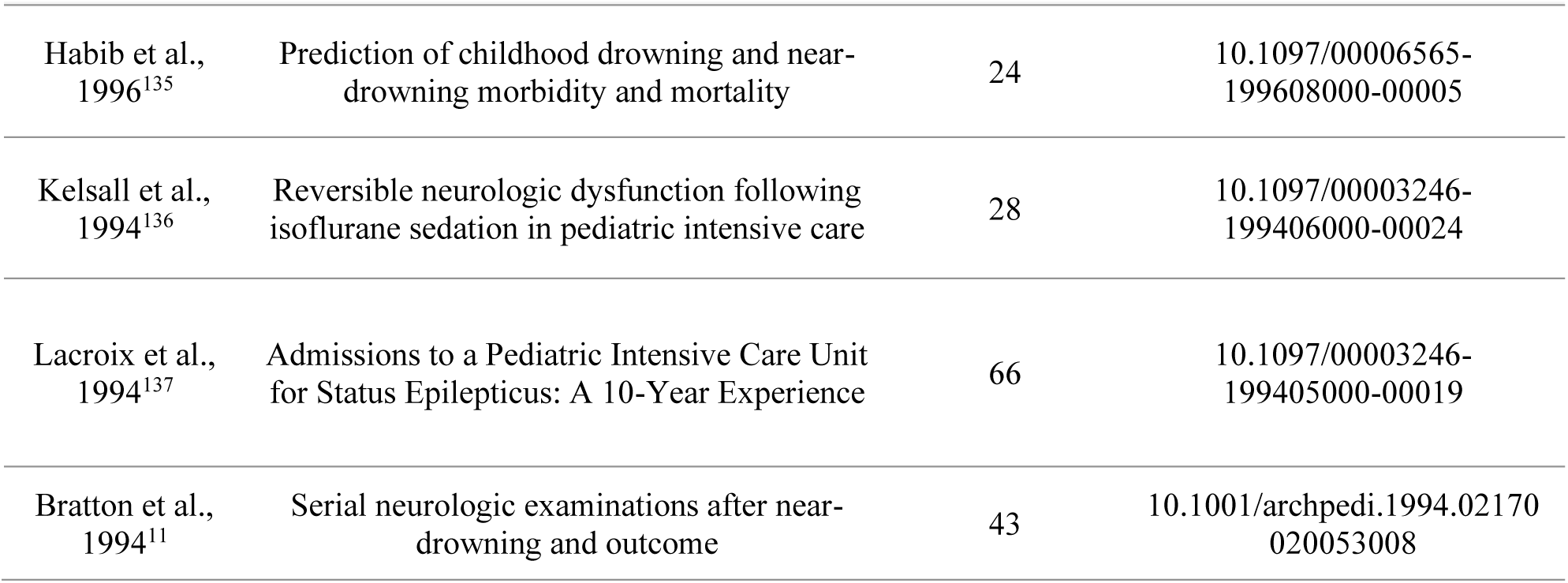
Selected articles approaching neurological examination in critically ill pediatric patients.

**Table 2.** Journals of publication.

| Journals | Publications (n) | Citations (n) | Impact factor |
| --- | --- | --- | --- |
| Acta Cardiologica Sinica | 1 | 7 | 1.8 |
| Anales de Pediatria | 2 | 10 | 2.1 |
| Archives of Pediatrics and Adolescent Medicine | 1 | 43 | 5.7 |
| Brain | 1 | 278 | 11.7 |
| Brain & Development | 1 | 6 | 1.3 |
| British Journal of Neurosurgery | 1 | 2 | 0.8 |
| Child Abuse & Neglect | 1 | 3 | 3.4 |
| Children | 1 | 2 | 2.1 |
| Child's Nervous System | 2 | 14 | 1.2 |
| Circulation | 1 | 129 | 38.6 |
| Critical Care | 2 | 17 | 9.3 |
| Critical Care and Resuscitation | 1 | 9 | 1.7 |
| Critical Care Explorations | 1 | 0 | 2.7 |
| Critical Care Medicine | 6 | 214 | 6.0 |
| Cureus Journal of Medical Science | 1 | 0 | 1.3 |
| Dubai Medical Journal | 1 | 0 | 0.6 |
| Epilepsy & Behavior Reports | 1 | 6 | 1.5 |
| European Journal of Paediatric Neurology | 1 | 1 | 2.3 |
| European Journal of Pediatrics | 1 | 36 | 2.6 |
| Frontiers in Pediatrics | 5 | 29 | 2.0 |
| Indian Journal of Critical Care Medicine | 1 | 5 | 1.5 |
| Indian Journal of Pediatrics | 1 | 2 | 2.0 |
| Indian Pediatrics | 1 | 1 | 1.5 |
| Intensive Care Medicine | 1 | 27 | 21.2 |
| JAMA Network Open | 1 | 1 | 9.7 |
| Journal of Child Neurology | 3 | 27 | 1.6 |
| Journal of Child Science | 1 | 1 | 0.4 |
| Journal of Clinical Neurophysiology | 1 | 34 | 1.7 |
| Journal of Critical Care Medicine | 1 | 0 | 1.7 |
| Journal of Emergency Medicine | 1 | 4 | 1.3 |
| Journal of Intensive Care Medicine | 2 | 9 | 2.1 |
| Journal of Neurology | 1 | 24 | 4.6 |
| Journal of Neurosurgery | 1 | 44 | 3.6 |
| Journal of Neurosurgery - Pediatrics | 4 | 26 | 2.1 |
| Journal of Neurotrauma | 1 | 3 | 3.8 |
| Journal of Pediatric Epilepsy | 1 | 0 | 0.1 |
| Journal of Pediatric Intensive Care | 2 | 5 | 0.3 |
| Journal of Pediatric Neurology | 1 | 0 | 0.2 |
| Journal of Pediatric Surgery | 1 | 1 | 2.5 |
| Journal of Pediatrics | 1 | 14 | 3.5 |
| Journal of Postgraduate Medicine | 1 | 0 | 1.2 |
| Journal of Stroke & Cerebrovascular Diseases1.8 | 1 | 28 | 1.8 |
| Journal of Trauma - Injury, Infection and Critical Care | 1 | 23 | Impact factor not available |
| Journal of Tropical Pediatrics | 2 | 2 | 1.1 |
| Medicina Balear | 1 | 0 | 0.5 |
| Neurocritical Care | 10 | 133 | 3.6 |
| Neurological Sciences | 1 | 0 | 2.4 |
| Neurology | 2 | 26 | 8.5 |
| Neuropediatrics | 1 | 5 | 1.2 |
| Pediatric Anesthesia and Critical Care Journal | 1 | 0 | 0.1 |
| Pediatric Critical Care Medicine | 20 | 556 | 4.5 |
| Pediatric Emergency Care | 4 | 41 | 1.2 |
| Pediatric Neurology | 6 | 63 | 2.1 |
| Pediatric Radiology | 1 | 49 | 2.3 |
| Pediatric Reports | 1 | 2 | 1.4 |
| Pediatrics | 2 | 141 | 6.4 |
| Resuscitation | 7 | 269 | 4.6 |
| Translational Pediatrics | 1 | 2 | 1.7 |
| Trends in Pediatrics | 1 | 0 | 0.2 |
| Turkish Journal of Pediatrics | 2 | 2 | 0.8 |
| Ultrasound | 1 | 0 | 0.7 |
| Ulusal Travma ve Acil Cerrahi Dergisi-Turkish Journal of Trauma & Emergency Surgery | 1 | 0 | 1 |
| World Neurosurgery | 1 | 4 | 2.1 |

### 3.4 Mapping Productivity: Researcher-level analysis

Regarding classical bibliometric parameters, we examined author contributions, keyword frequencies, and the journals of publication. In the author analysis, we identified a total of 791 researchers who contributed to at least one article (Figure 4A). The most prolific author in this field was Berg RA, reaching a total of 10 publications. In this sense, his scientific influence is emphasized by the fact that their research group was also the most prolific (Figure 4B,C), such as Nadkarni VM and Topijan AA, both high-productive researchers belonged to Berg research-network (Figure 4B,C). Considering citation analysis, Berg also stands out as lead researcher in this field, with a total of 405 citations (Figure 4D), followed by Topijan with a total of 248 citations. Interesting, among the most cited articles, there are some authors who published one article, which received higher levels of citation (e.g., Zhao XY et al.) (Figure 4D). In order to identify other cited authors who published more than 2 articles, we evaluated the citation scores range 200 to 405 citations (Figure 4E).

**Figure 4.**
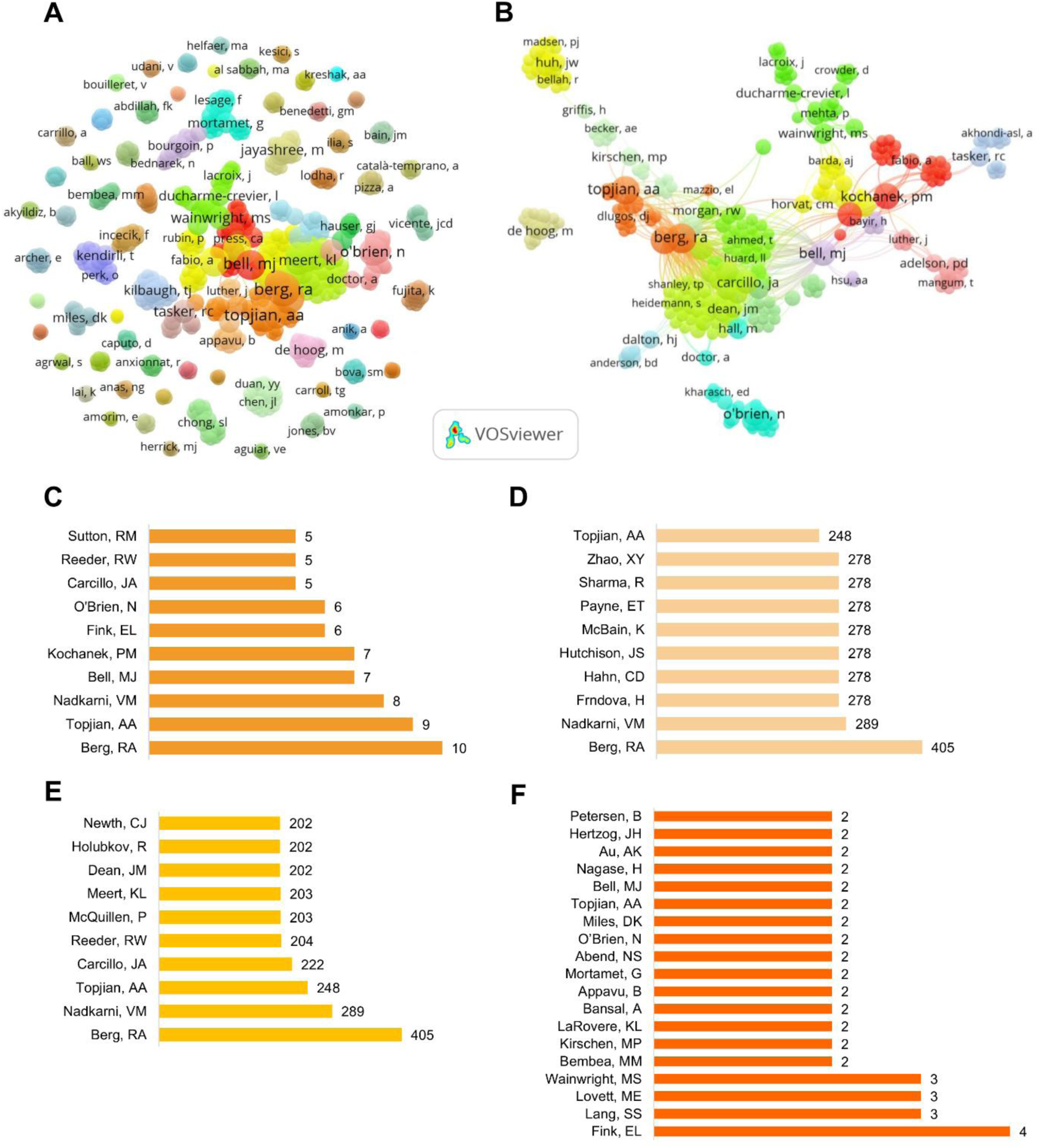
Comprehensive author-level analysis. Co-authorship network visualized by the number of publications (A). Most productive collaborative network is displayed, with a central cluster indicating high interconnectivity among prolific researchers (B). Top 10 most productive authors based on the number of publications (C), and top 10 most cited authors overall (D). To better contextualize sustained scientific impact, we present the top 10 most cited authors who contributed to two or more articles (E). Finally, researchers who served as corresponding authors in at least two studies, reflecting leadership roles in study coordination and manuscript submission (F).

In a more meticulous way, we also mapped corresponding authors to gain insight into institutional leadership and patterns of responsibility for study coordination and manuscript submission (Figure 4F). This analysis revealed a diversified landscape of leading researchers, highlighting researchers such as Fink EL, Lang SS, Kirschen MP, Appavu B, and Au AK as frequent corresponding authors. Their recurrent role in this position suggests consistent engagement in project leadership and editorial responsibility, rather than occasional contribution.

### 3.5 From Words to Research Topics: Co-occurrence keywords-level analysis

Figure 5 presents the most frequently occurring keywords identified across the selected articles. The term “pediatric” was the most prevalent, appearing 22 times, followed closely by “traumatic brain injury” (n = 21) and “children” (n = 18), which underscore the central focus on age group and primary neurological conditions within pediatric critical care. Other frequently cited keywords included “cardiac arrest” (n = 14), “neurocritical care” and “pediatric intensive care unit” (n = 10 each), as well as “cardiopulmonary resuscitation” (n = 9), all indicating the prominence of acute, life-threatening events requiring specialized neurological evaluation (Figure 5B).

**Figure 5.**
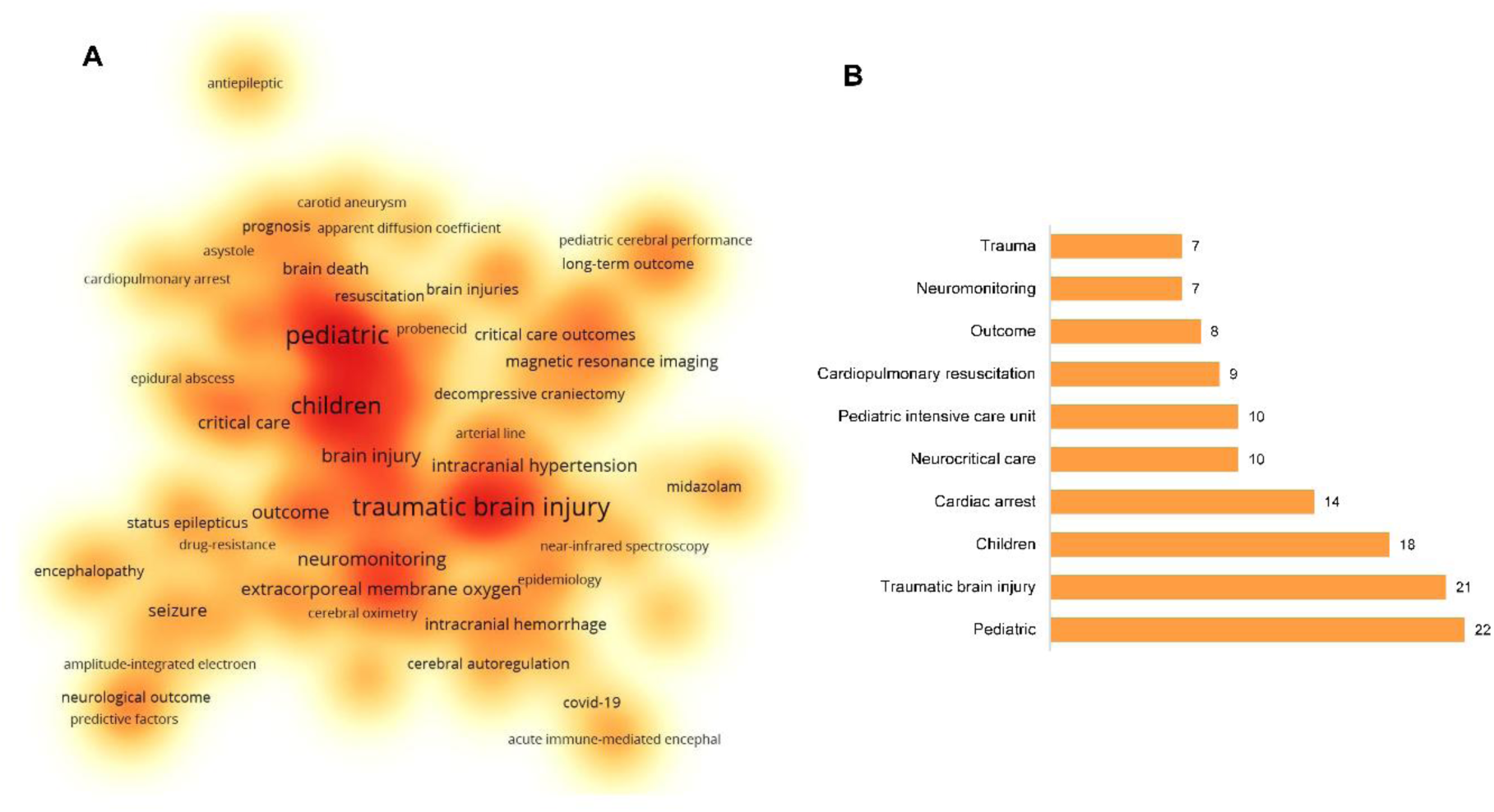
Network visualization of co-occurring keywords. Clusters are color-coded; node size represents keyword frequency, and lines indicate keyword connections (A). Top 10 most frequent keywords (B).

Additionally, keywords such as “outcome” (n = 8), “neuromonitoring” (n = 7), and “trauma” (n = 7) reflect a strong emphasis on diagnostic and prognostic approaches in the context of the PICU. Collectively, these findings suggest a research landscape primarily centered on severe neurological insults and the tools used to monitor and predict recovery in critically ill pediatric populations.

### 3.6 What Has Been Studied? A Study Design-Level Analysis

We categorized study designs according to the classifications provided by the original authors (Table 3). Notably, the period with the highest publication volume was 2011–2020, during which 11 different types of studies were identified. Considering the overall publication history, the most common were retrospective observational studies (n = 56), prospective observational studies (n = 20), case reports (n = 16), and retrospective cohort studies (n = 10). Conversely, less frequent designs included prognostic studies, prospective controlled clinical trials, prospective randomized clinical trials, and prospective controlled cohort studies, represented by a single publication.

**Table 3.** Study designs in the retrieved articles.

| Study design | Decades | Number of articles | Total number of articles | Total number of citations |
| --- | --- | --- | --- | --- |
| Retrospective observational study | 1991 - 2000 | 5 | 56 | 997 |
|  | 2001 - 2010 | 4 |  |  |
|  | 2011 - 2020 | 29 |  |  |
|  | 2021 - 2025 | 18 |  |  |
| Prospective observational study | 1991 - 2000 | 1 | 20 | 967 |
|  | 2001 - 2010 | 3 |  |  |
|  | 2011 - 2020 | 13 |  |  |
|  | 2021 - 2024 | 3 |  |  |
| Retrospective cohort | 1991 - 2000 | 1 | 10 | 183 |
|  | 2011 - 2020 | 3 |  |  |
|  | 2021 - 2025 | 6 |  |  |
| Prospective cohort | 2011 - 2020 | 1 | 7 | 58 |
|  | 2021 - 2025 | 6 |  |  |
| Retrospective and Prospective study | 2011 - 2020 | 2 | 2 | 30 |
| Case report | 1991 - 2000 | 1 | 16 | 56 |
|  | 2011 - 2020 | 4 |  |  |
|  | 2021 - 2025 | 11 |  |  |
| Case control | 2011 - 2020 | 1 | 1 | 14 |
| Cross-sectional | 2021 - 2025 | 2 | 2 | 6 |
| Case series | 2001 - 2010 | 2 | 7 | 85 |
|  | 2011 - 2020 | 1 |  |  |
|  | 2021 - 2025 | 4 |  |  |
| Cohort (A second analysis) | 2021 - 2025 | 2 | 2 | 3 |
| Prognostic study | 2021 - 2025 | 1 | 1 | 1 |
| Prospective cotrolled clinical trial | 2011 - 2020 | 1 | 1 | 17 |
| Prospective randomized trial | 2011 - 2020 | 1 | 1 | 15 |
| Secondary analysis of biospecimens from an Randomized Clinical Trial | 2011 - 2020 | 1 | 2 | 21 |
|  | 2021 - 2025 | 1 |  |  |

Citations were also analyzed by study type. Retrospective observational studies (n = 997), prospective observational studies (n = 967), retrospective cohort studies (n = 183), and case series (n = 85) accounted for the highest citation counts.

### 3.7 From Admission to Discharge: Mapping Neurological Practices in PICU

An overview of the selected studies revealed that neuromonitoring in the PICU has been investigated primarily in a limited set of clinical conditions. The majority of publications focused on traumatic brain injury (TBI), cardiac arrest, central nervous system infections, and encephalopathies, with fewer studies addressing drowning, Guillain-Barré syndrome, multisystem inflammatory syndrome in children (MIS-C), extracorporeal membrane oxygenation (ECMO)-related complications, and metabolic disorders. Across these conditions, the most frequently applied bedside tools were the Glasgow Coma Scale (GCS) at admission and the Pediatric Cerebral Performance Category (PCPC) or Pediatric Overall Performance Category (POPC) at discharge. Continuous EEG (cEEG) and neuroimaging, particularly computed tomography (CT) and magnetic resonance imaging (MRI), were predominantly applied during the PICU stay, while intracranial pressure (ICP) and cerebral perfusion pressure (CPP) monitoring were mainly reported in studies of traumatic brain injury (Table 4).

**Table 4.**
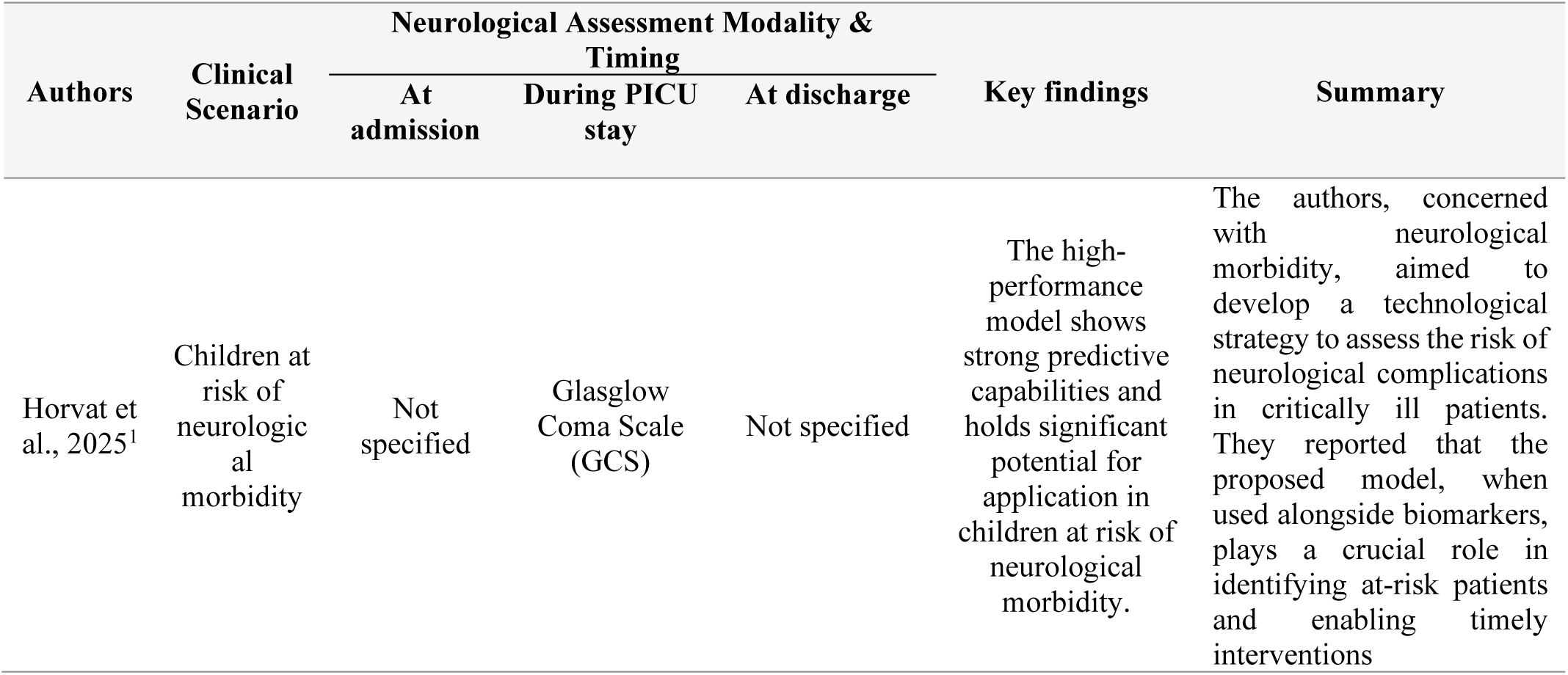
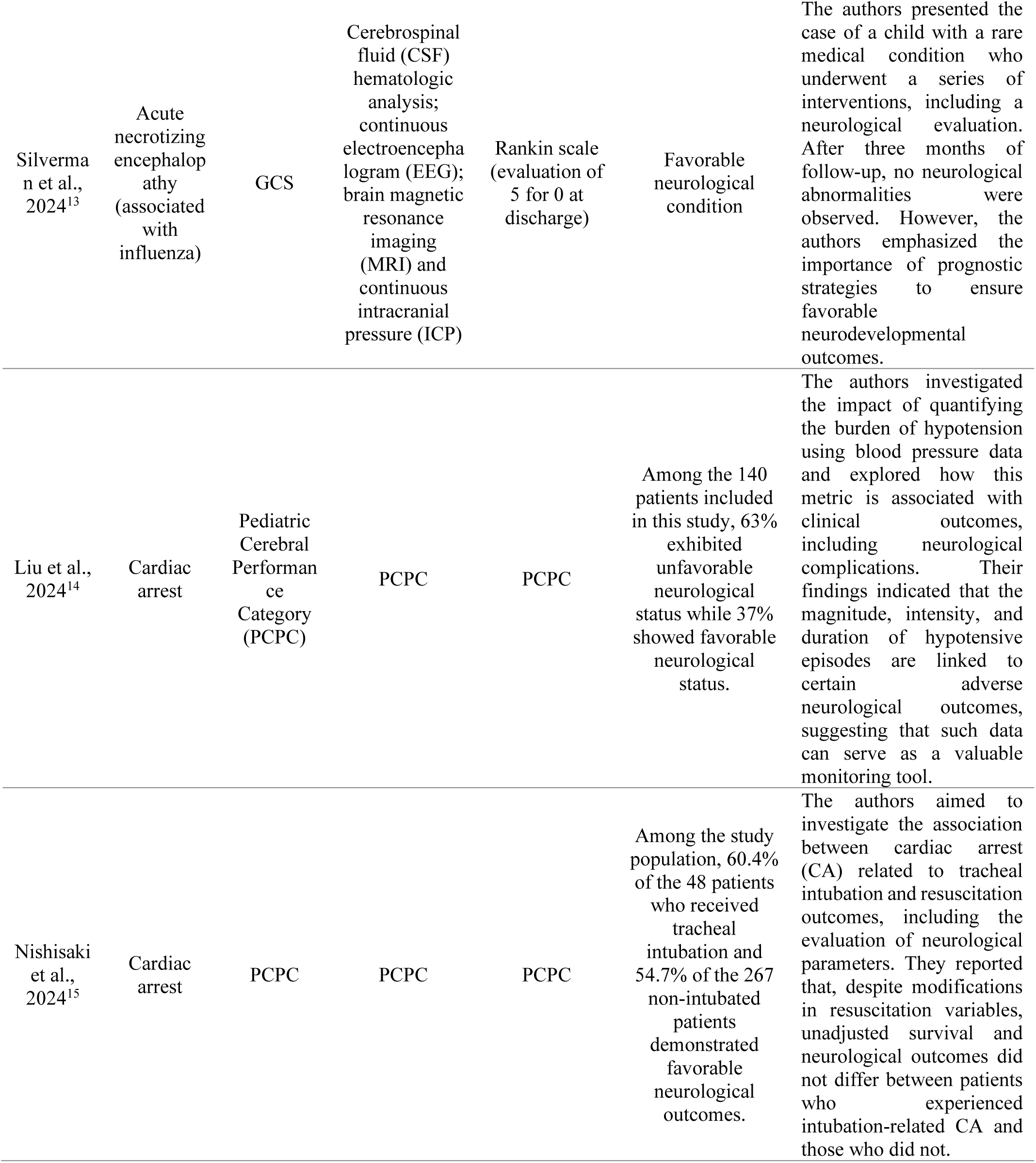
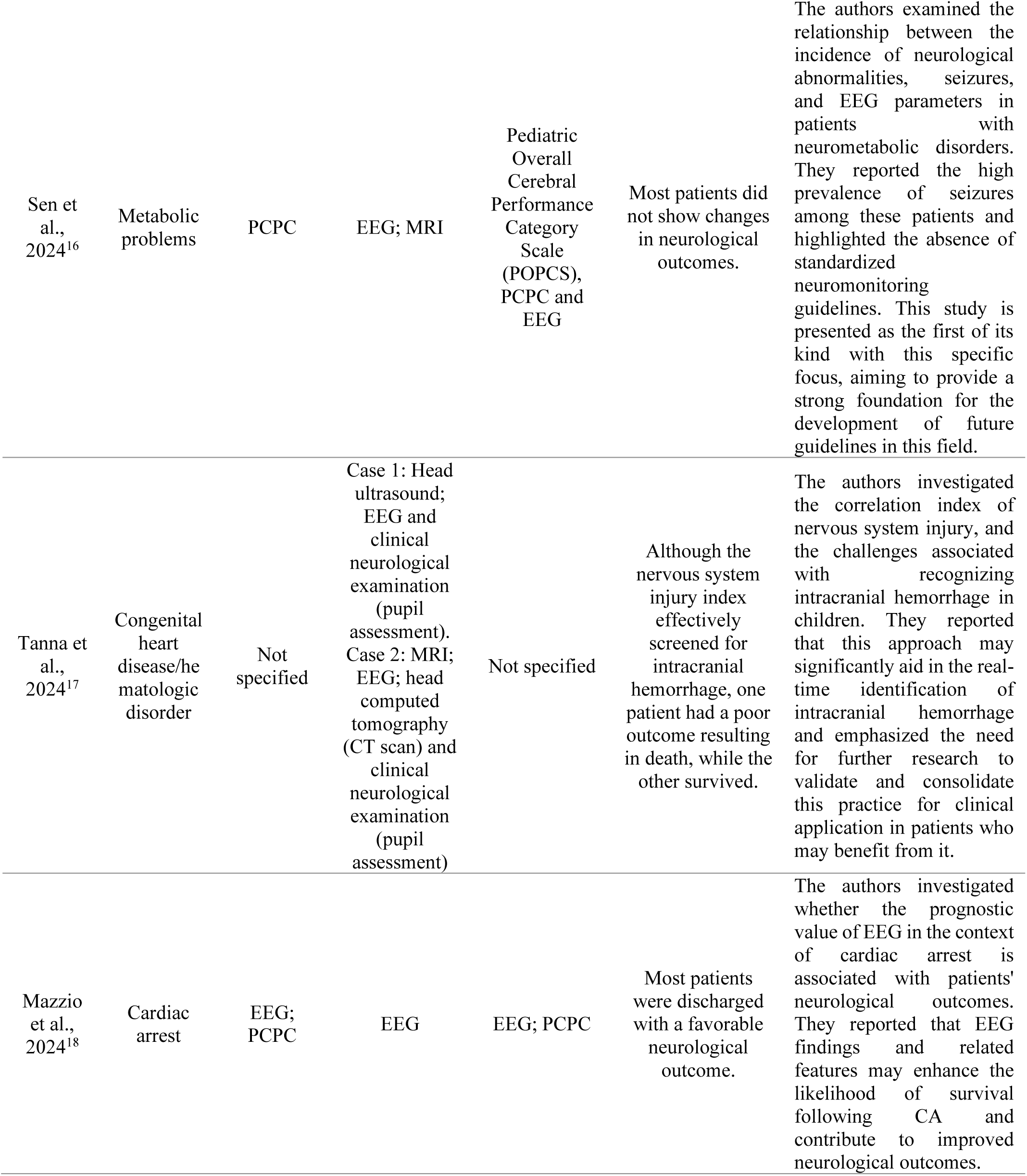
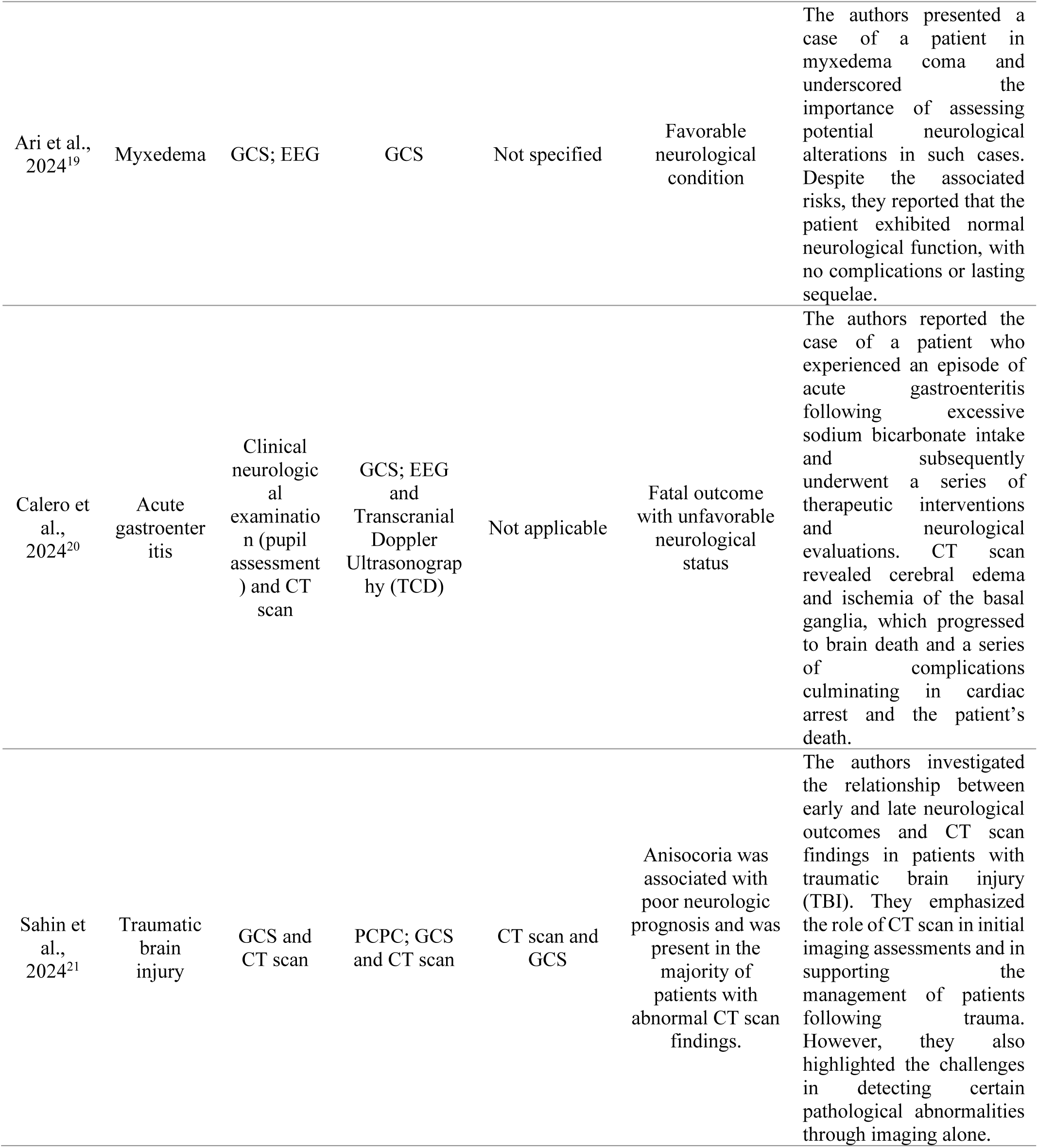
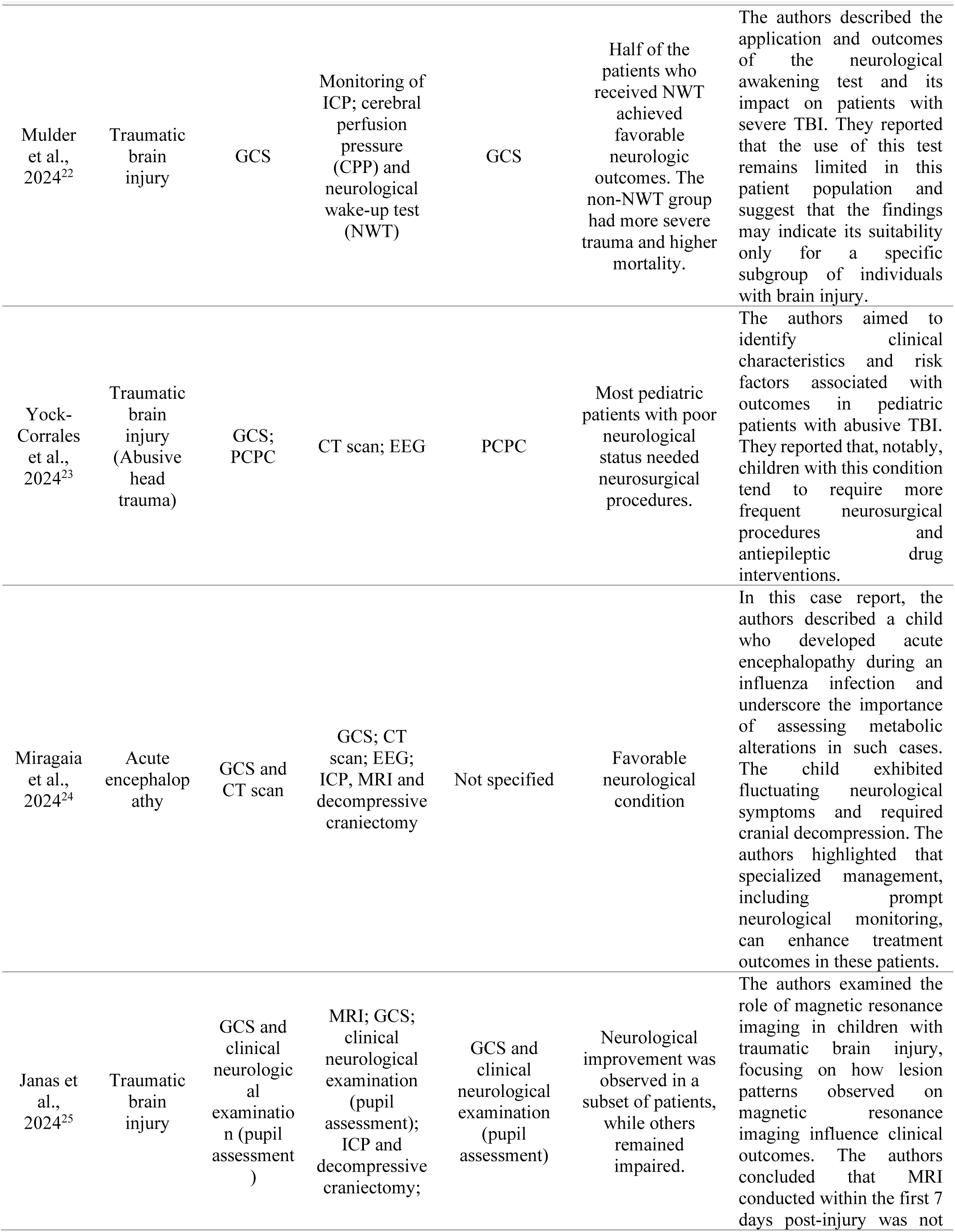
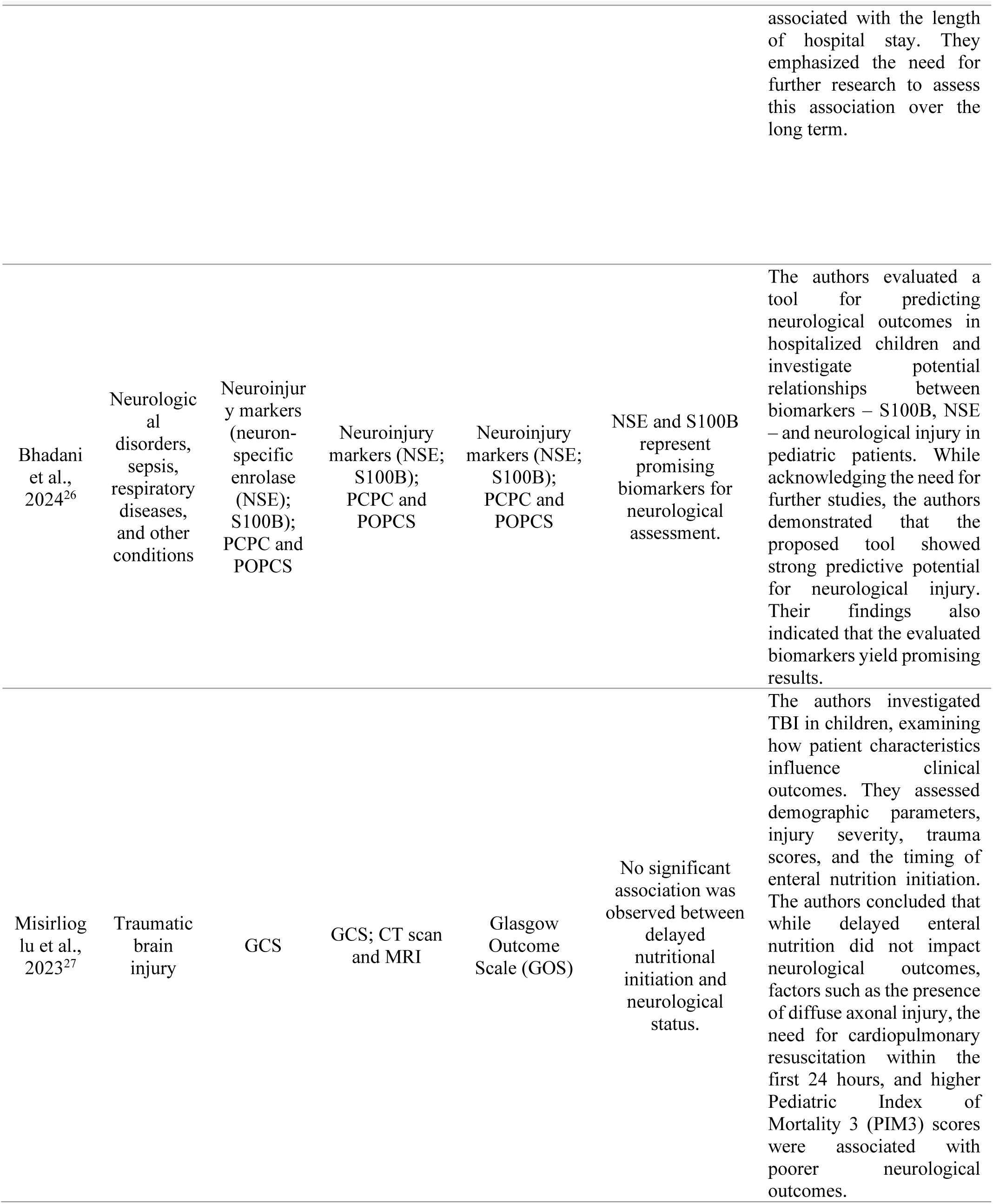
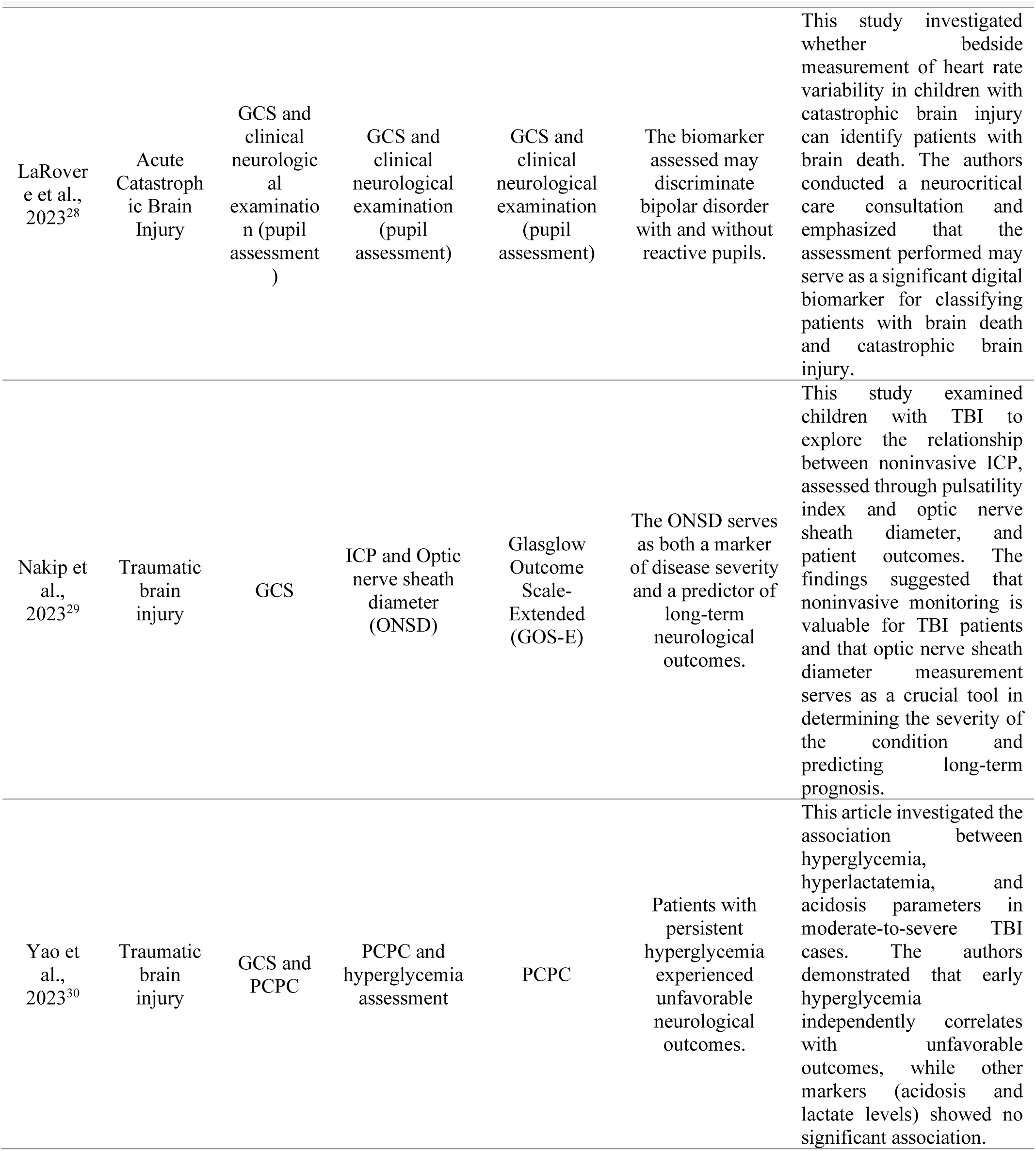
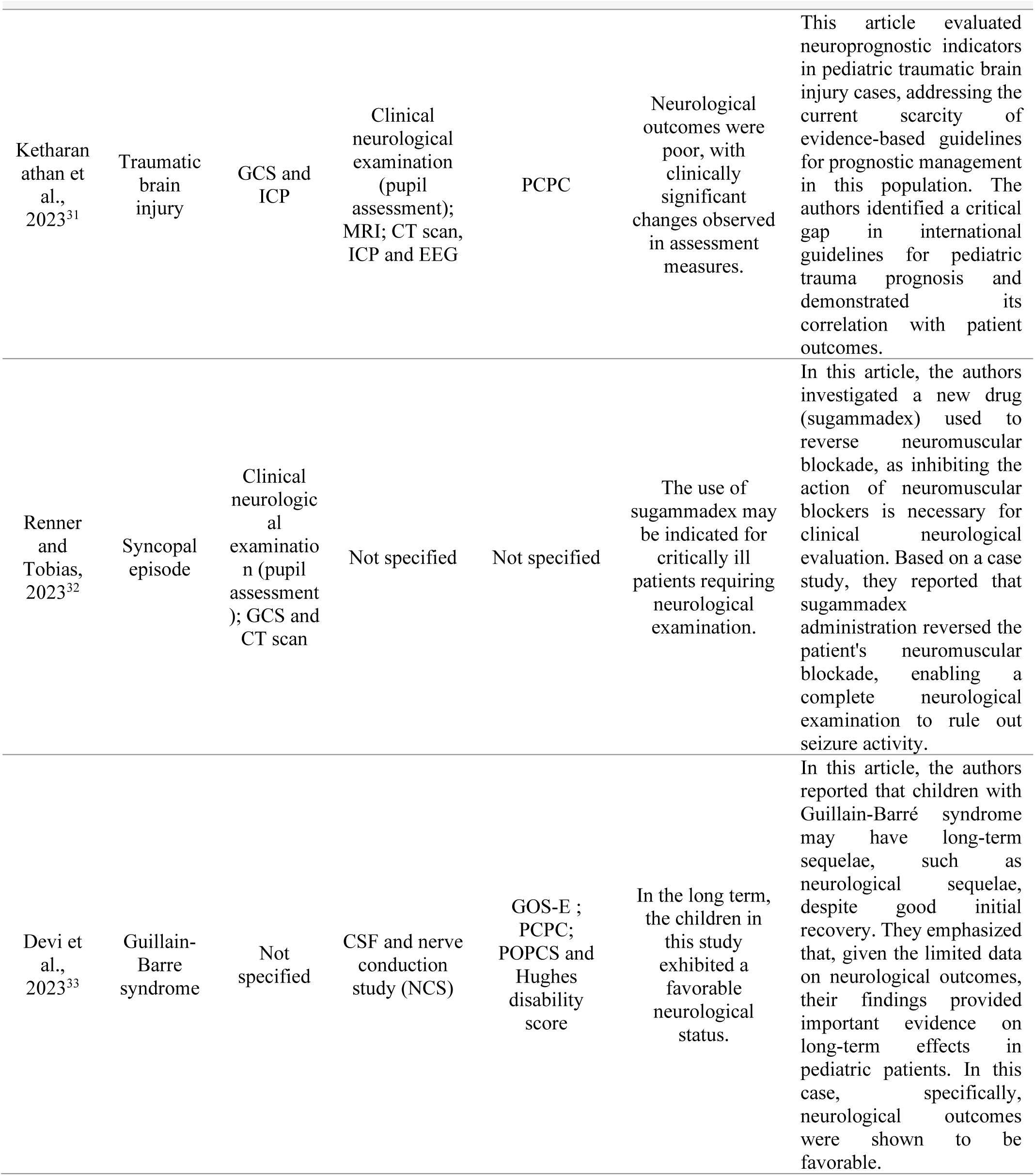
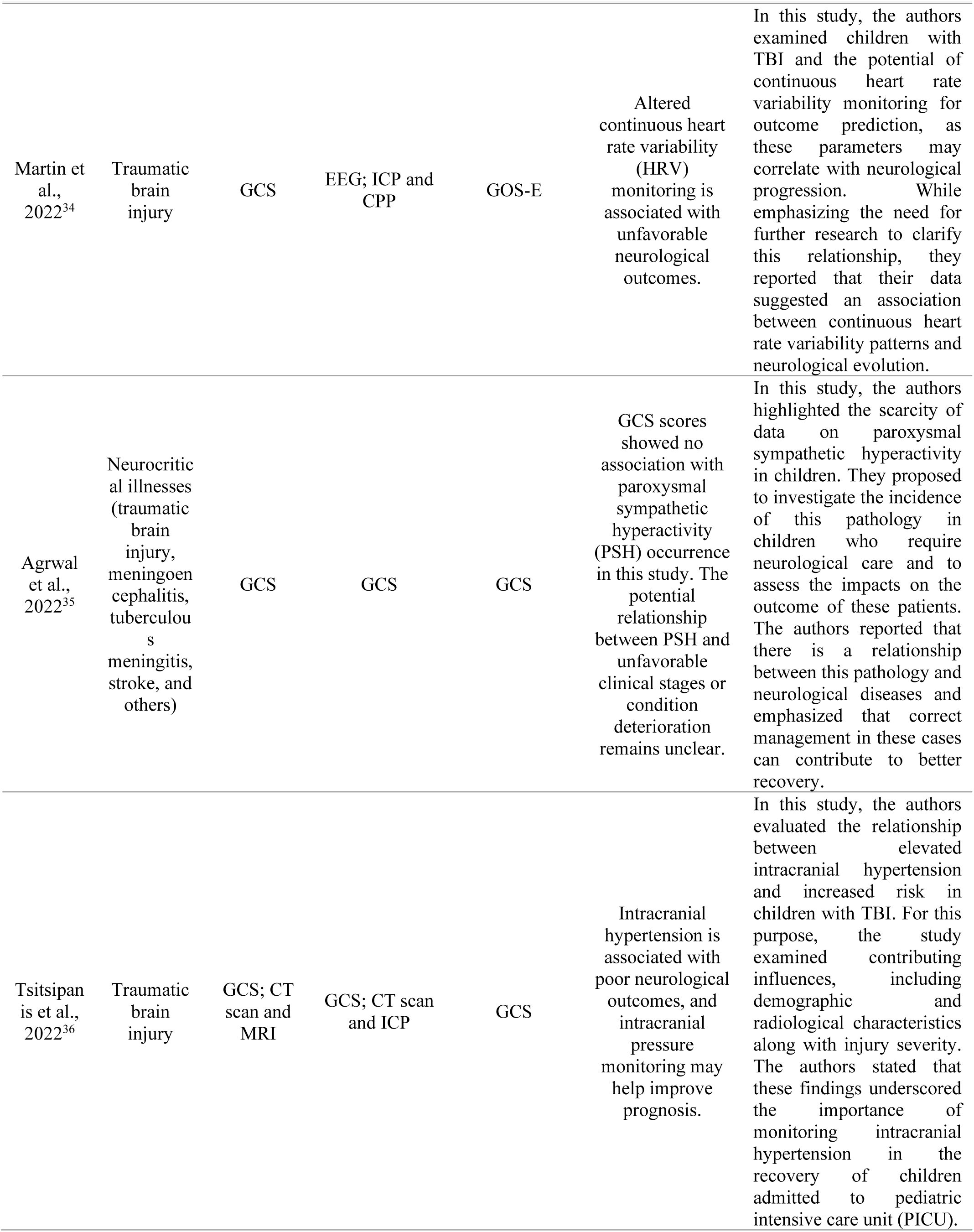
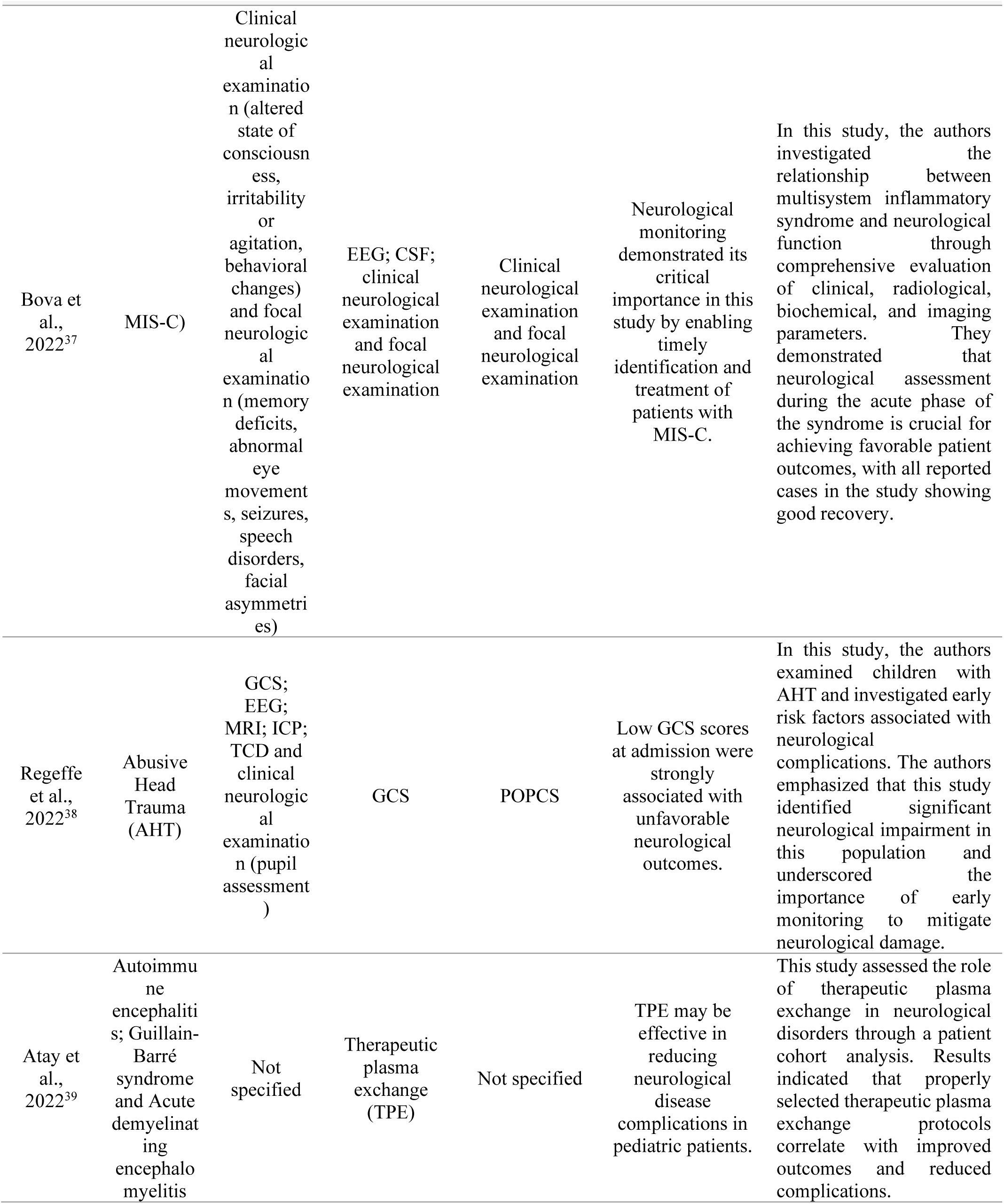
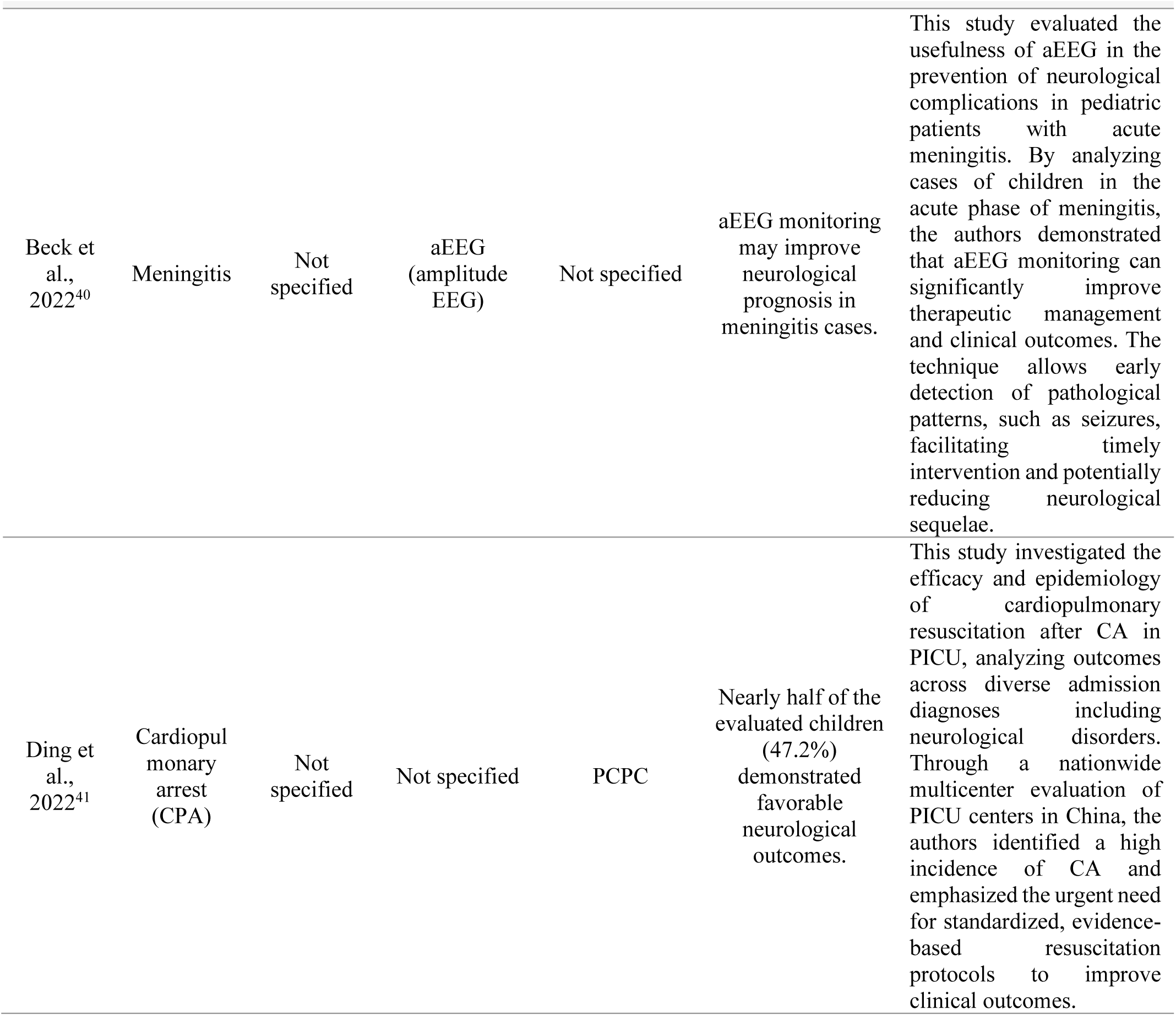
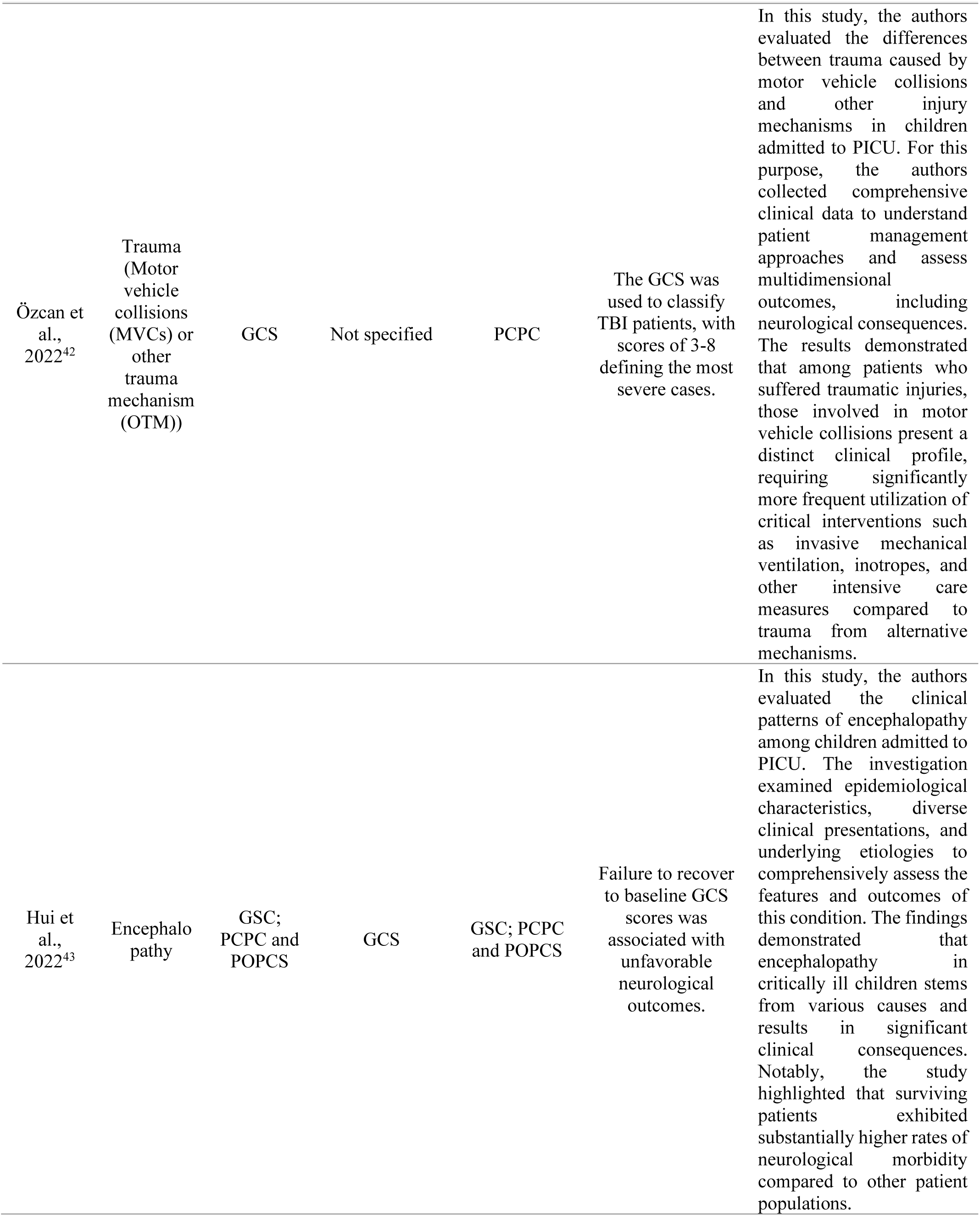
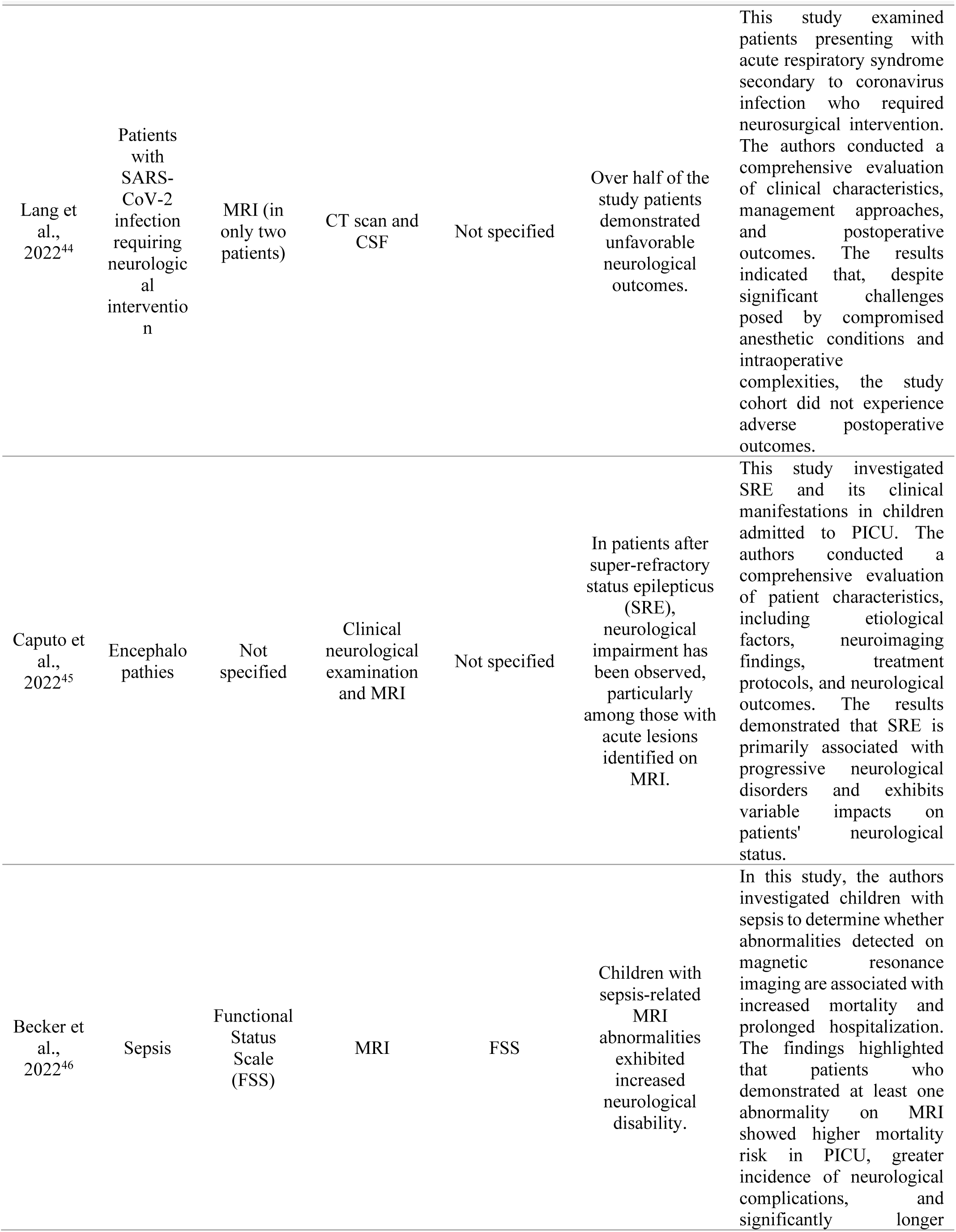
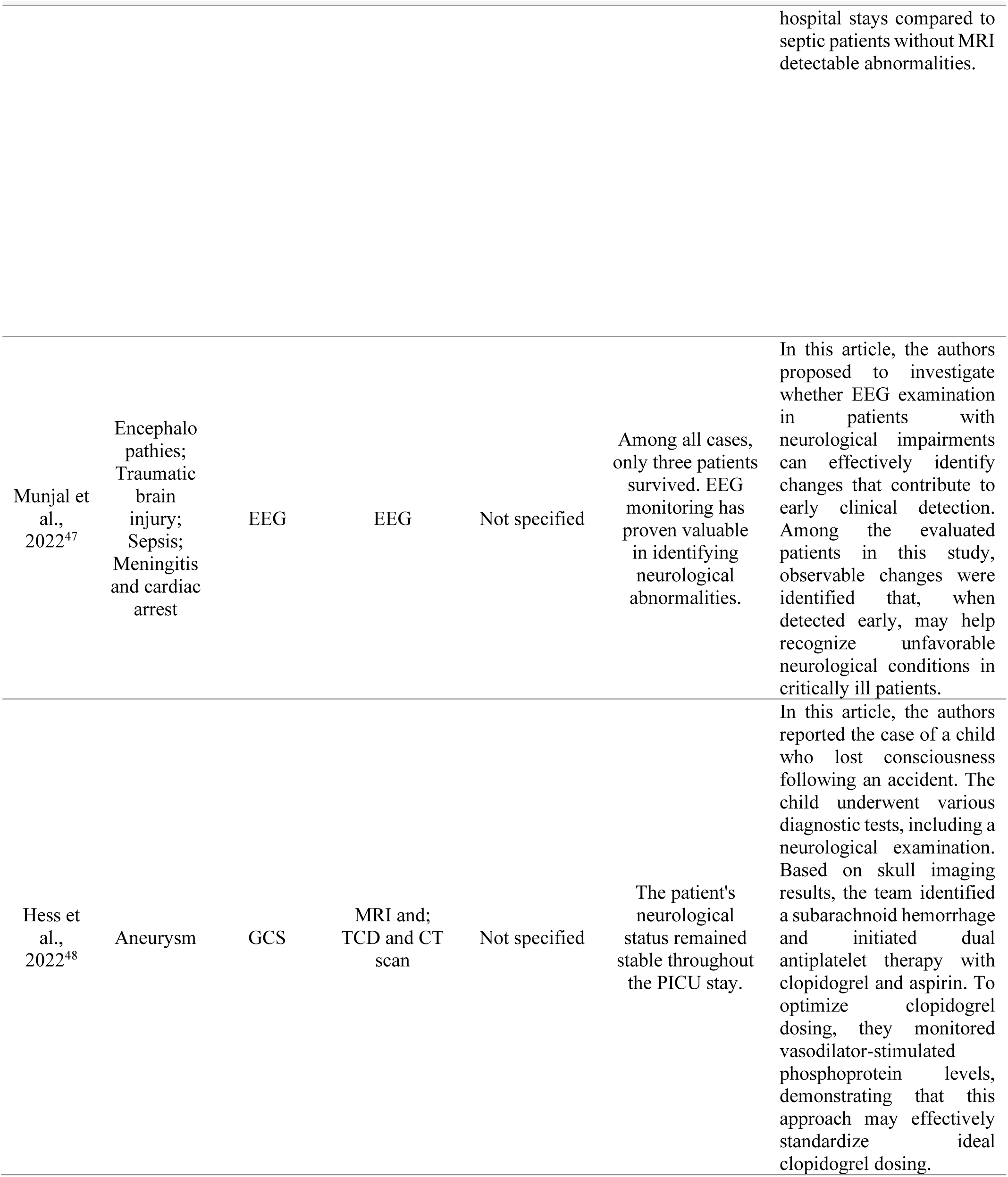
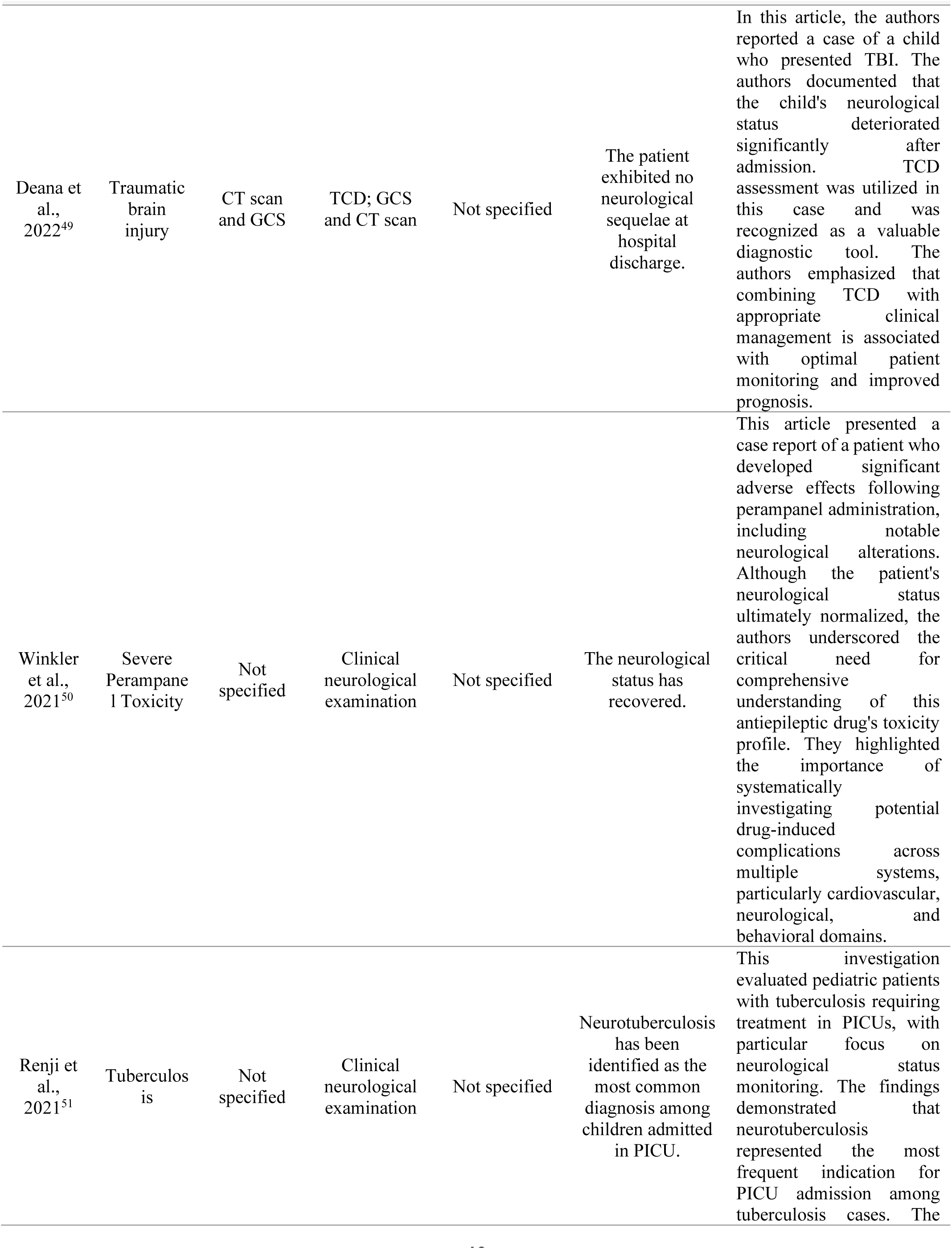
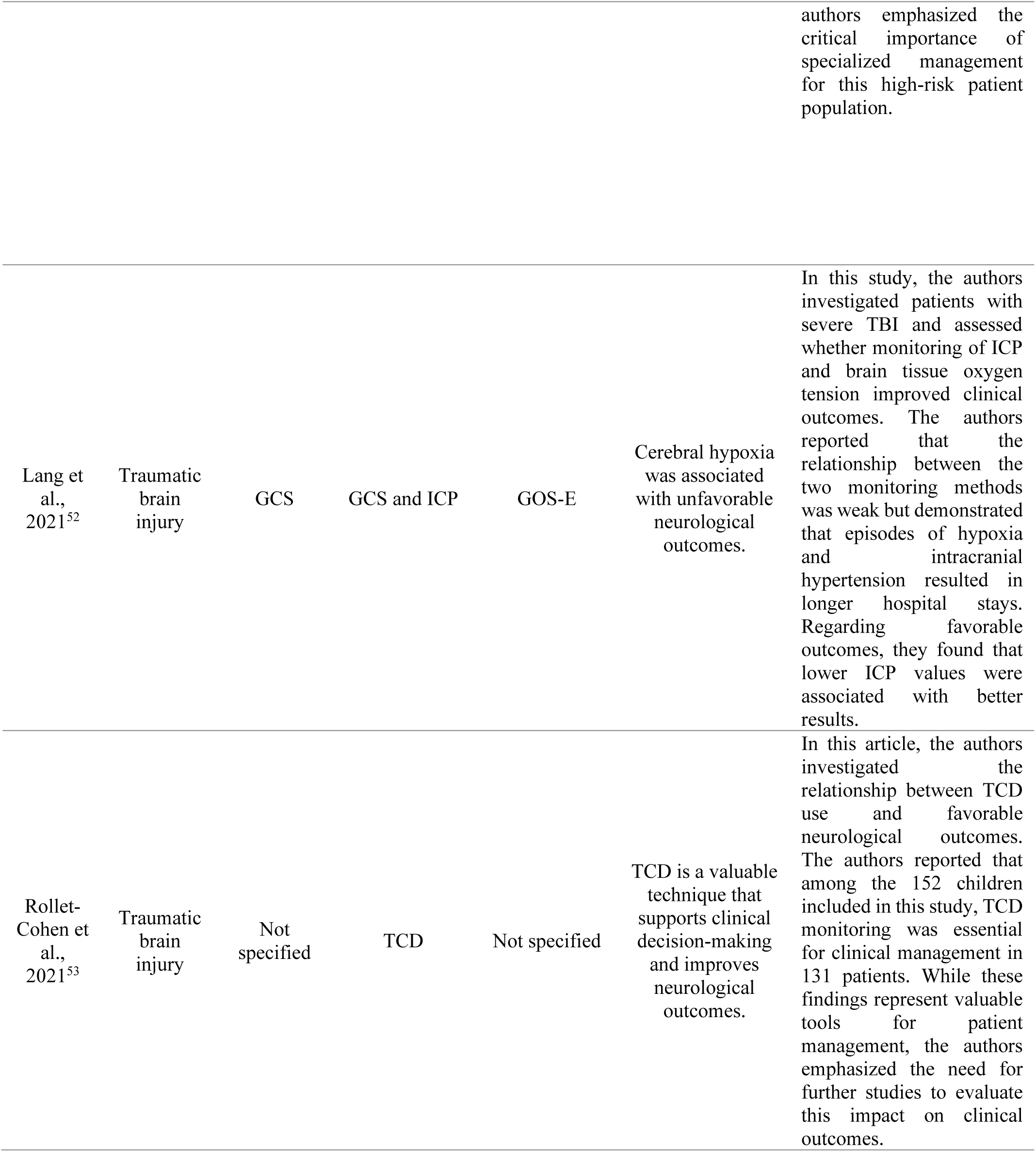
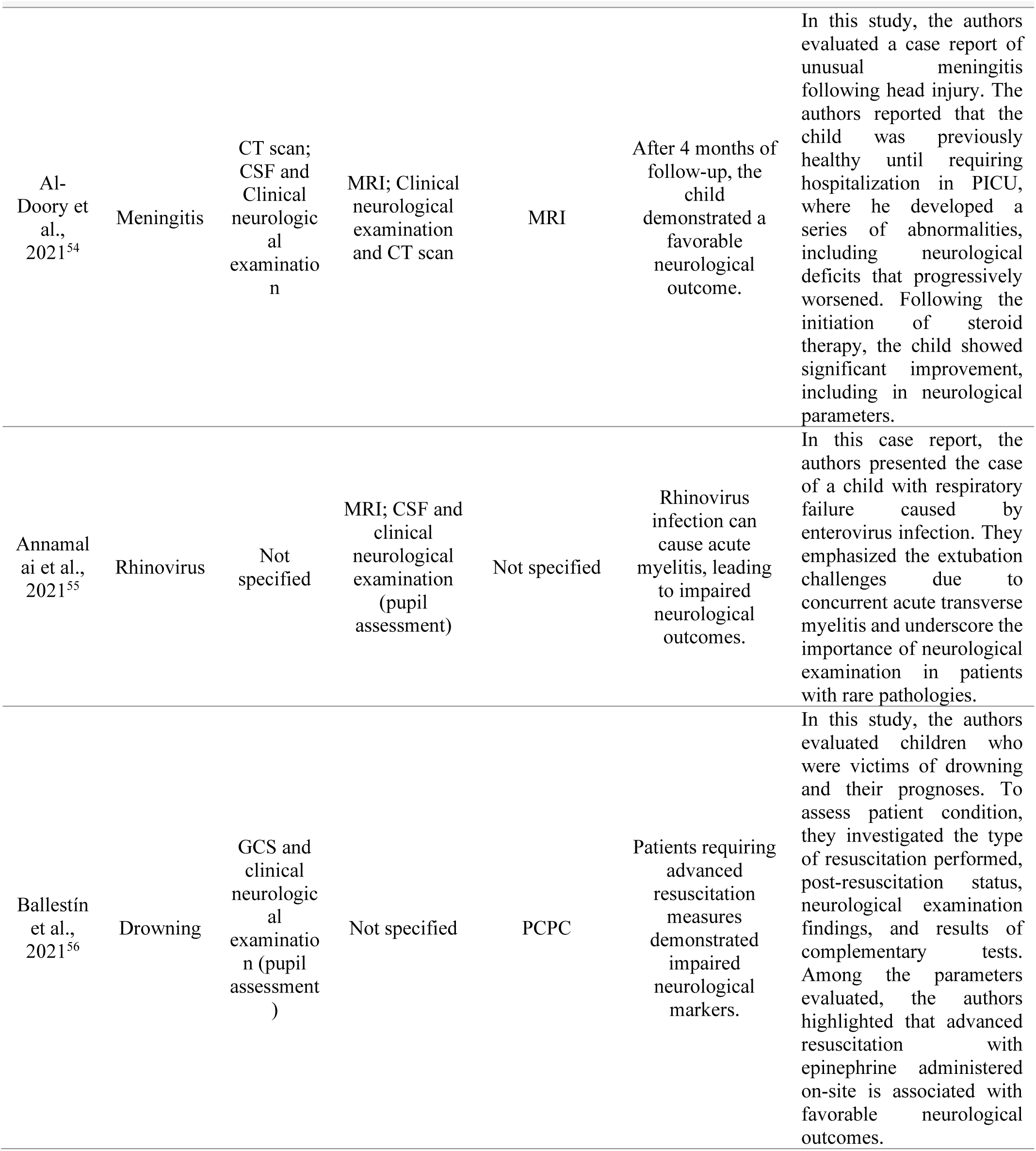
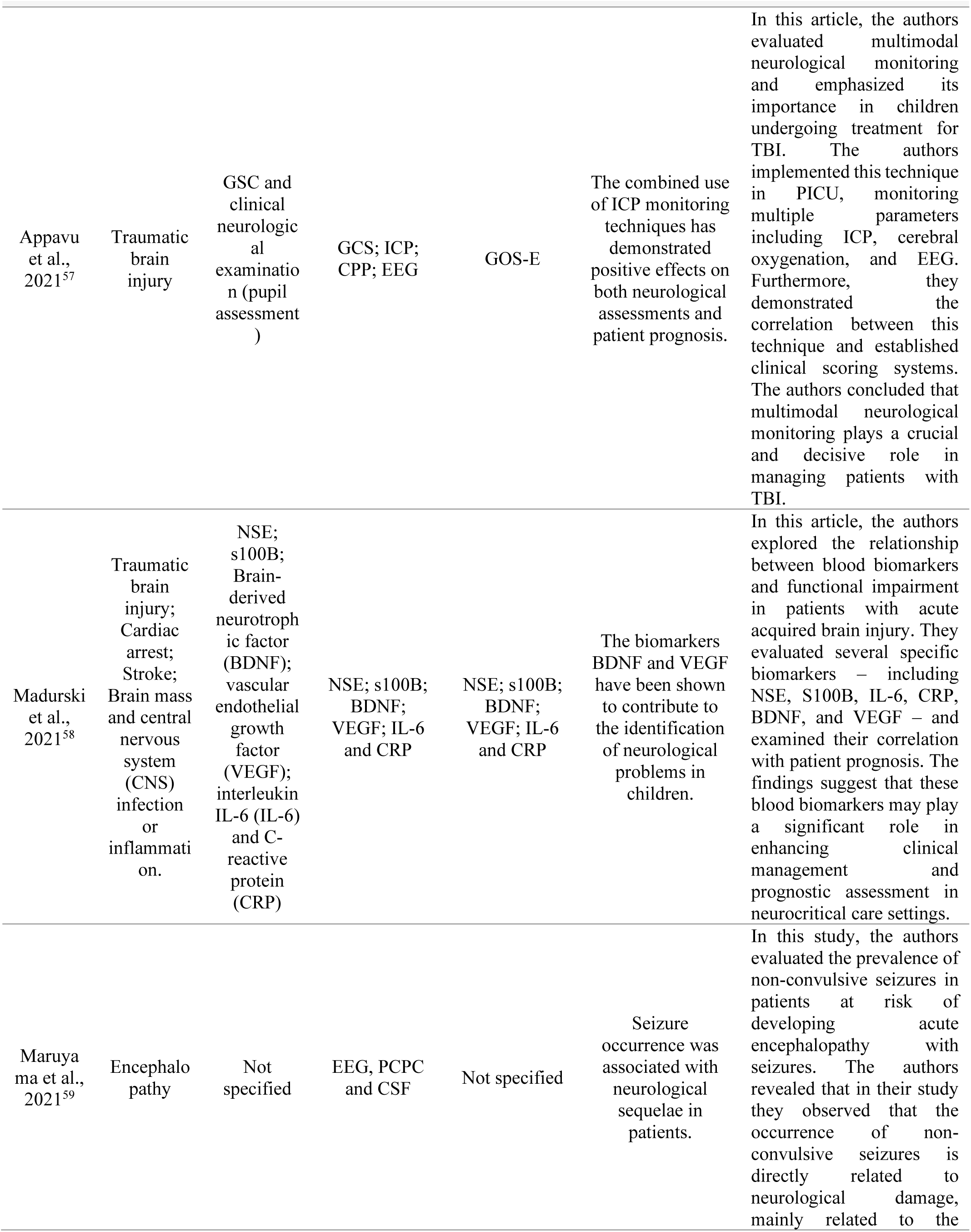
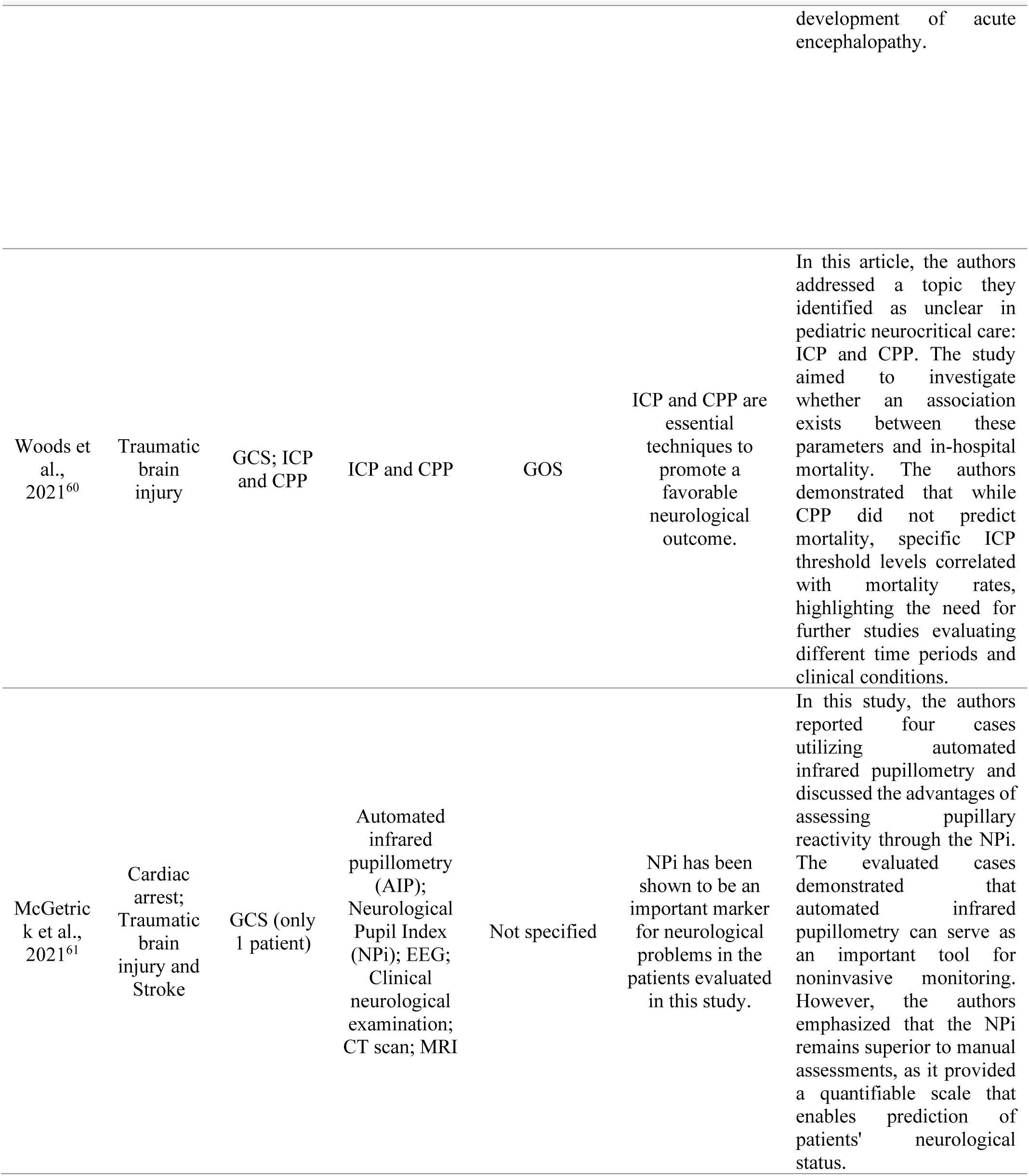
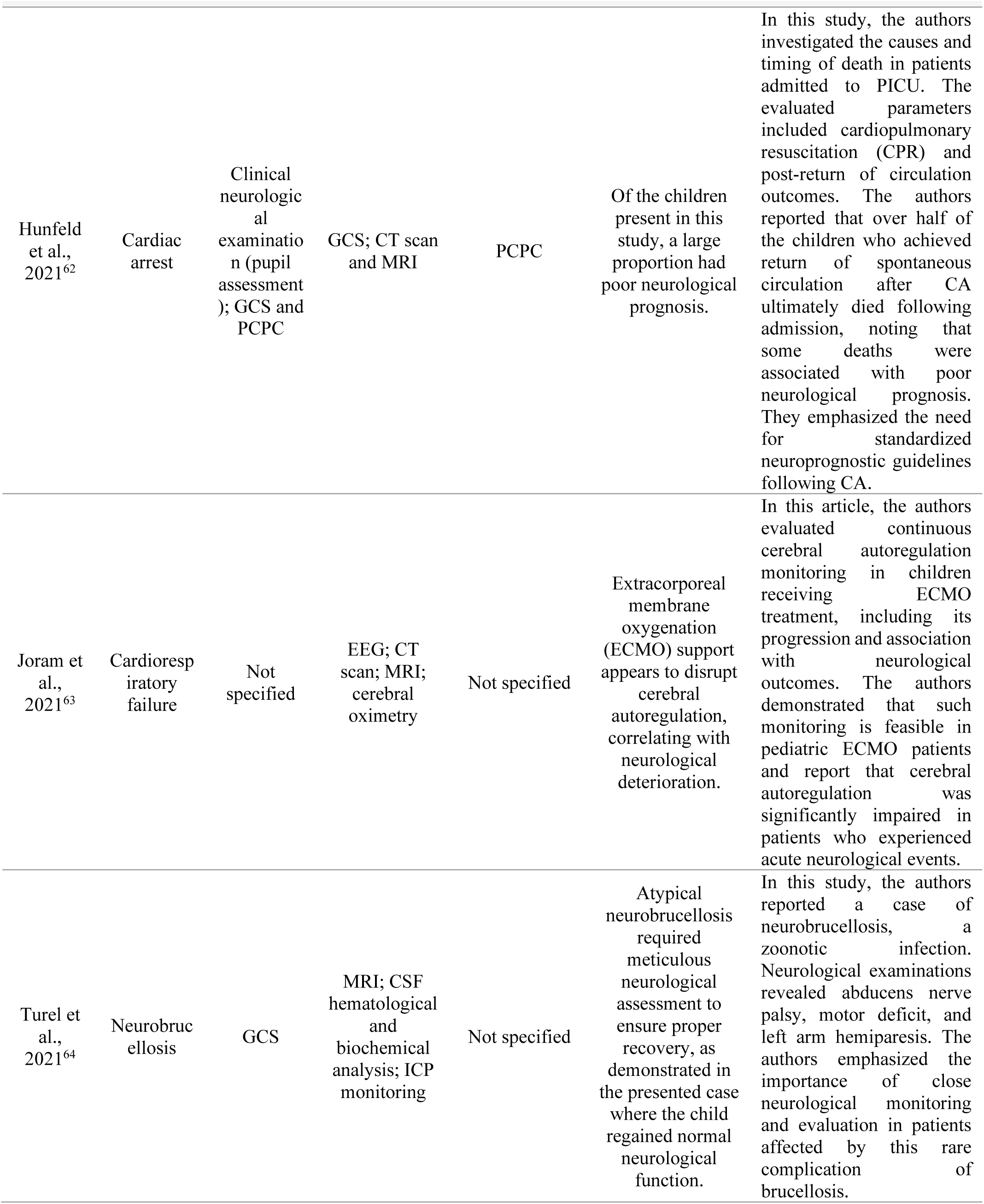
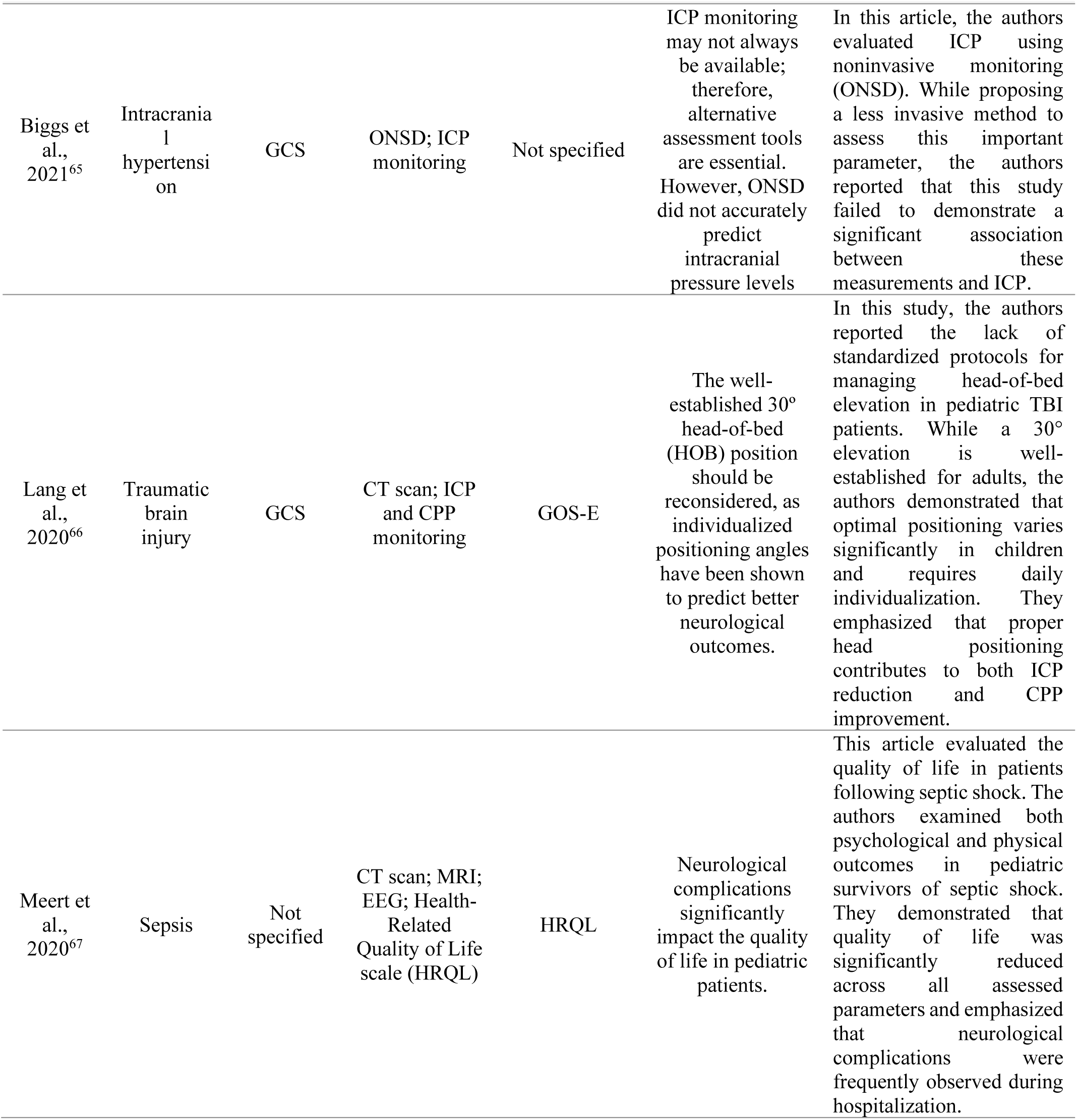
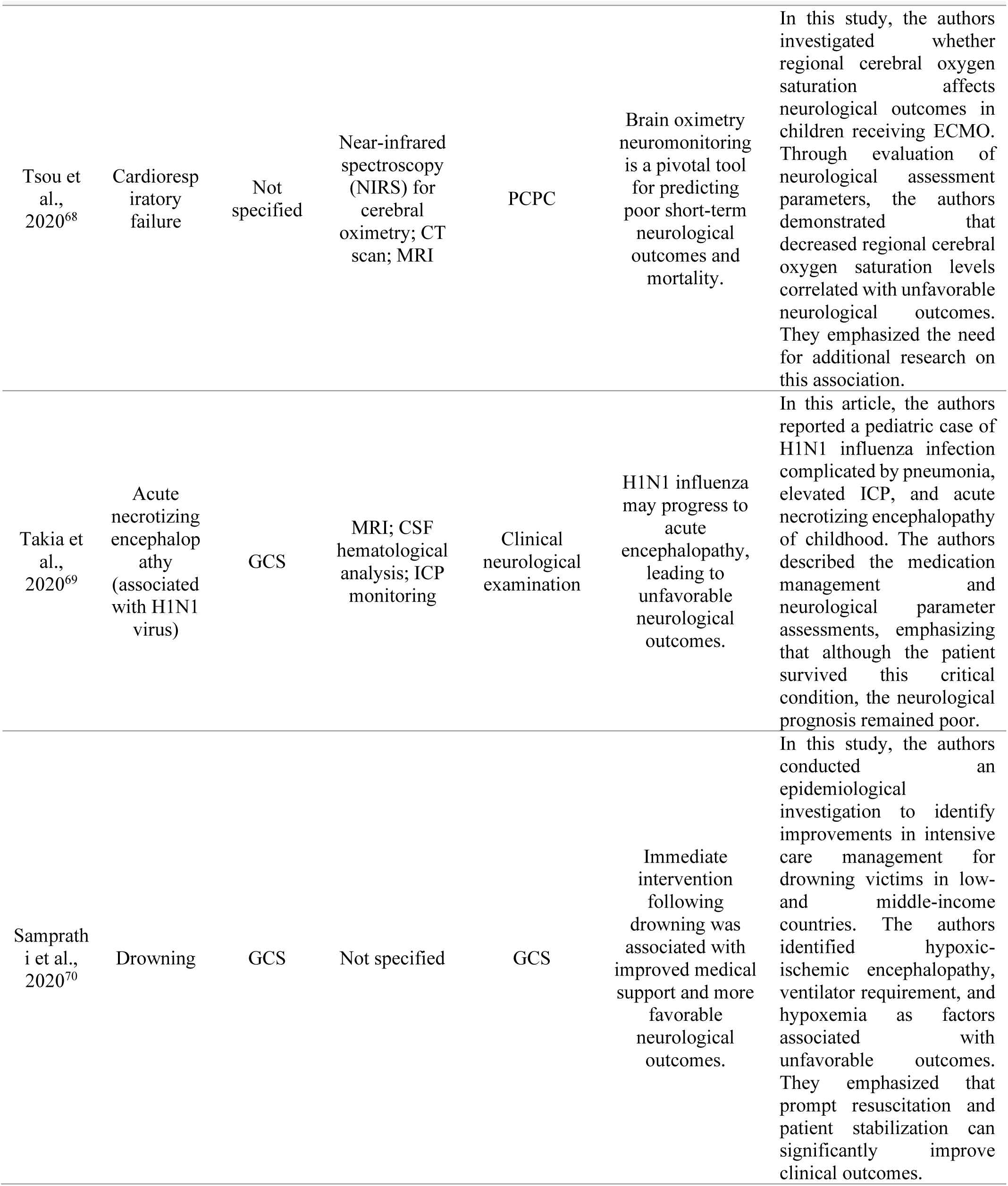
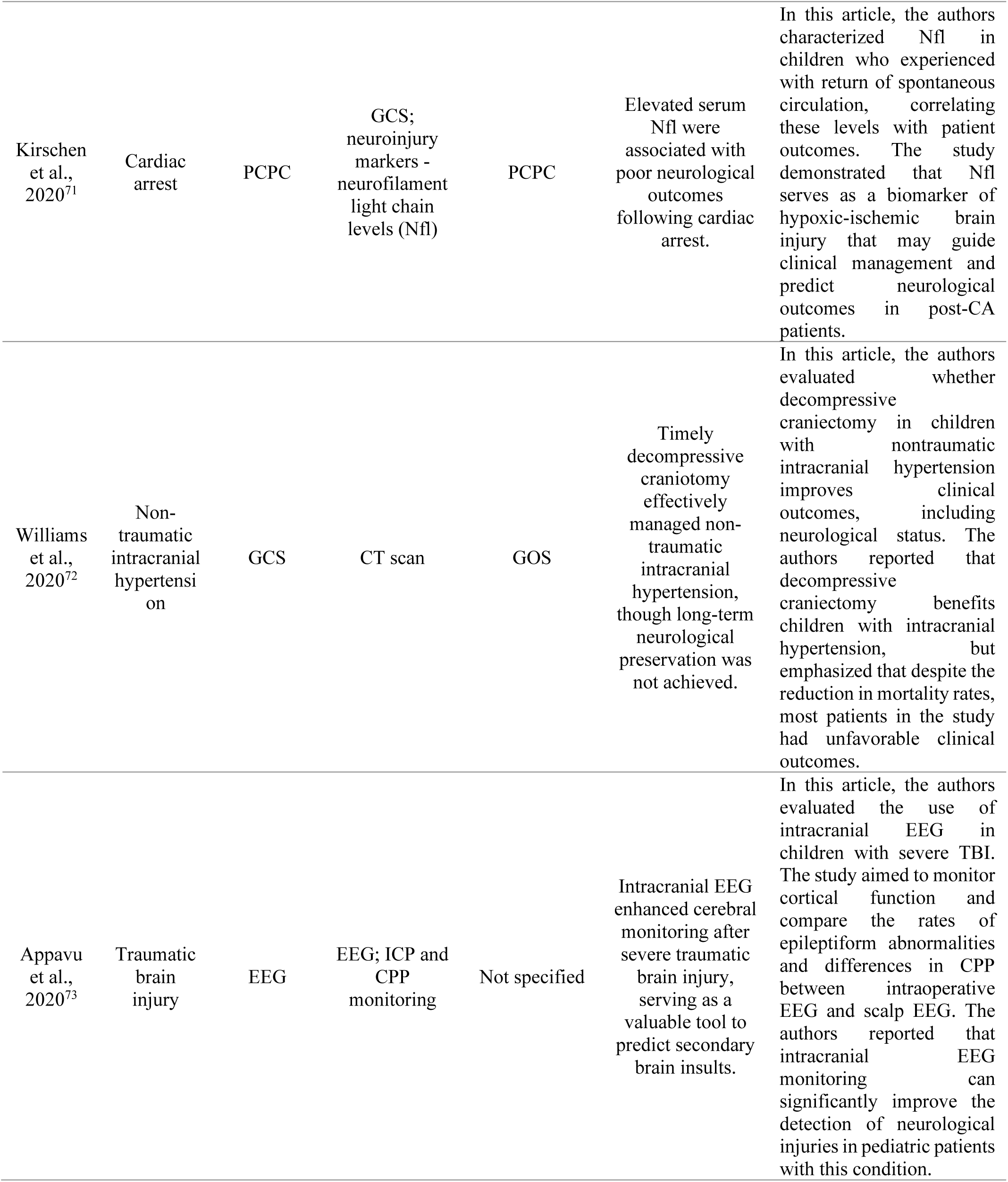
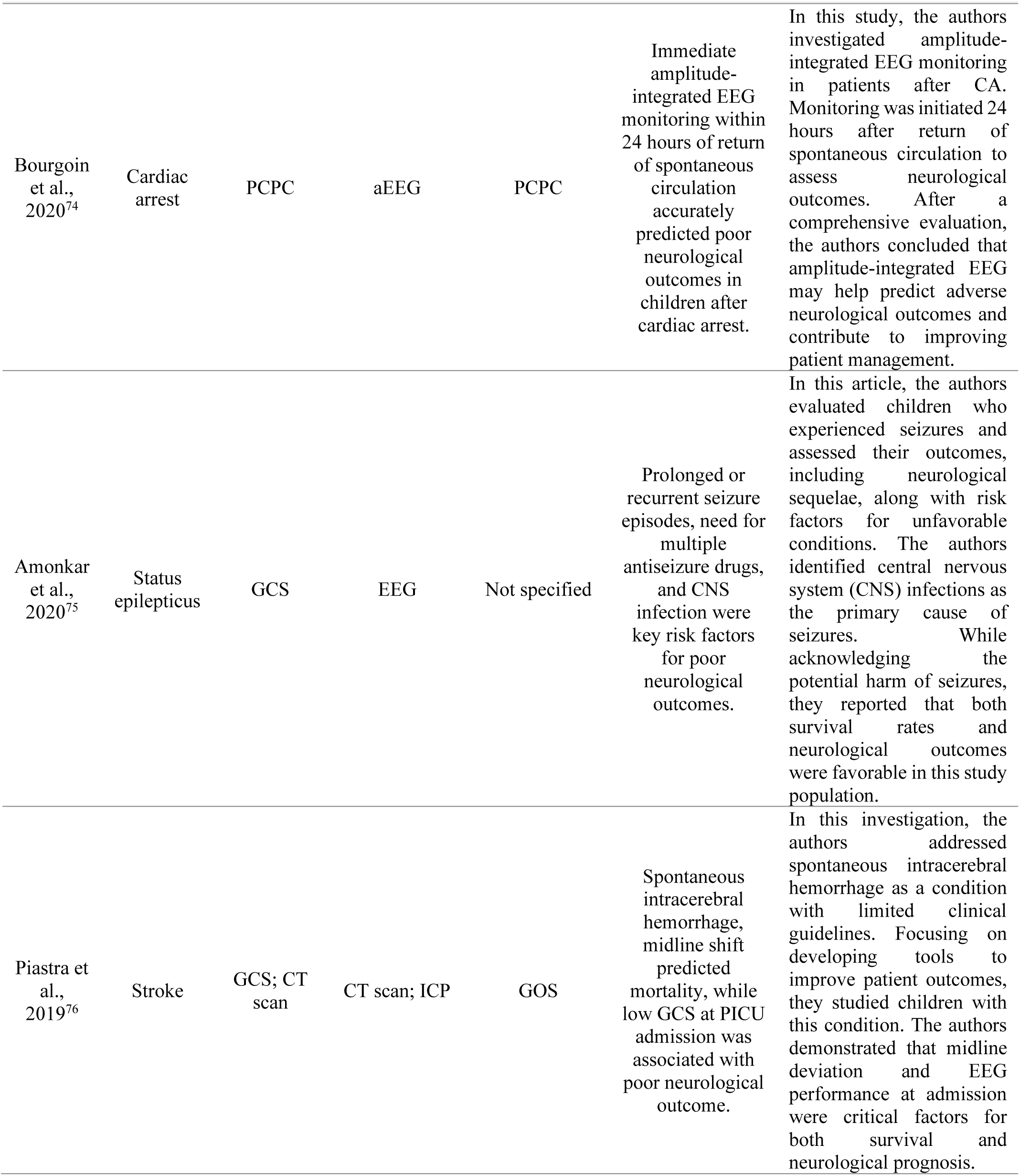
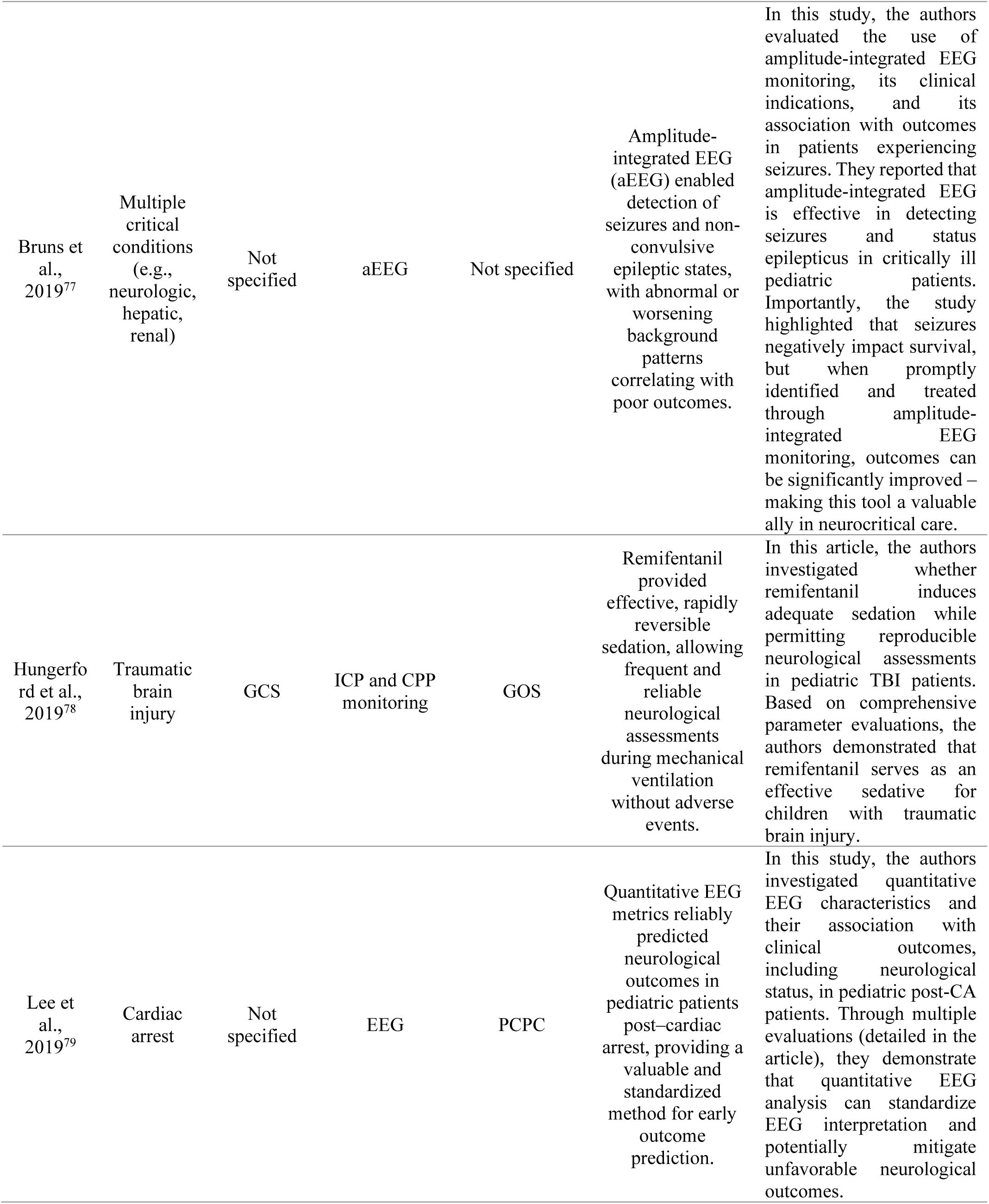
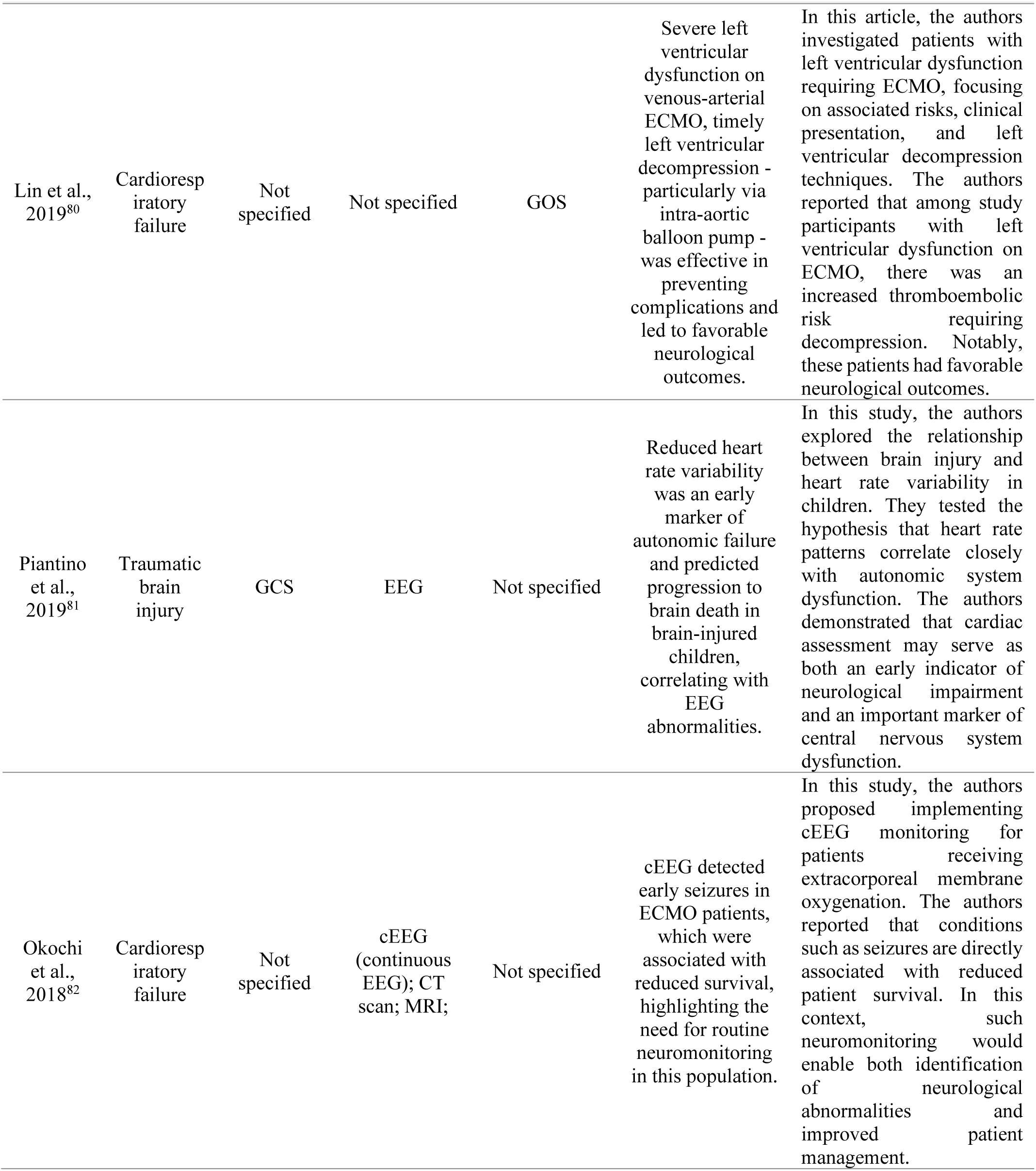
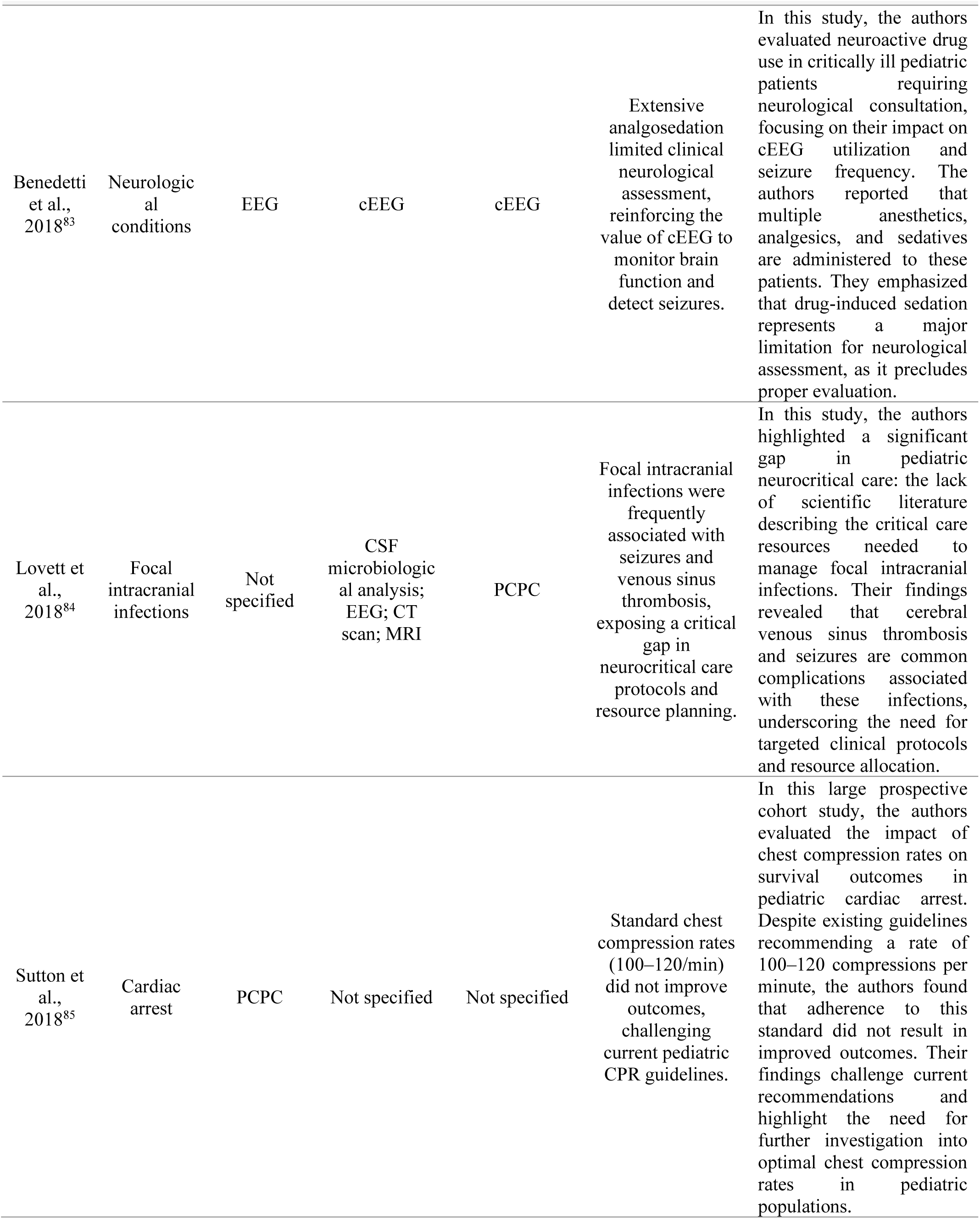
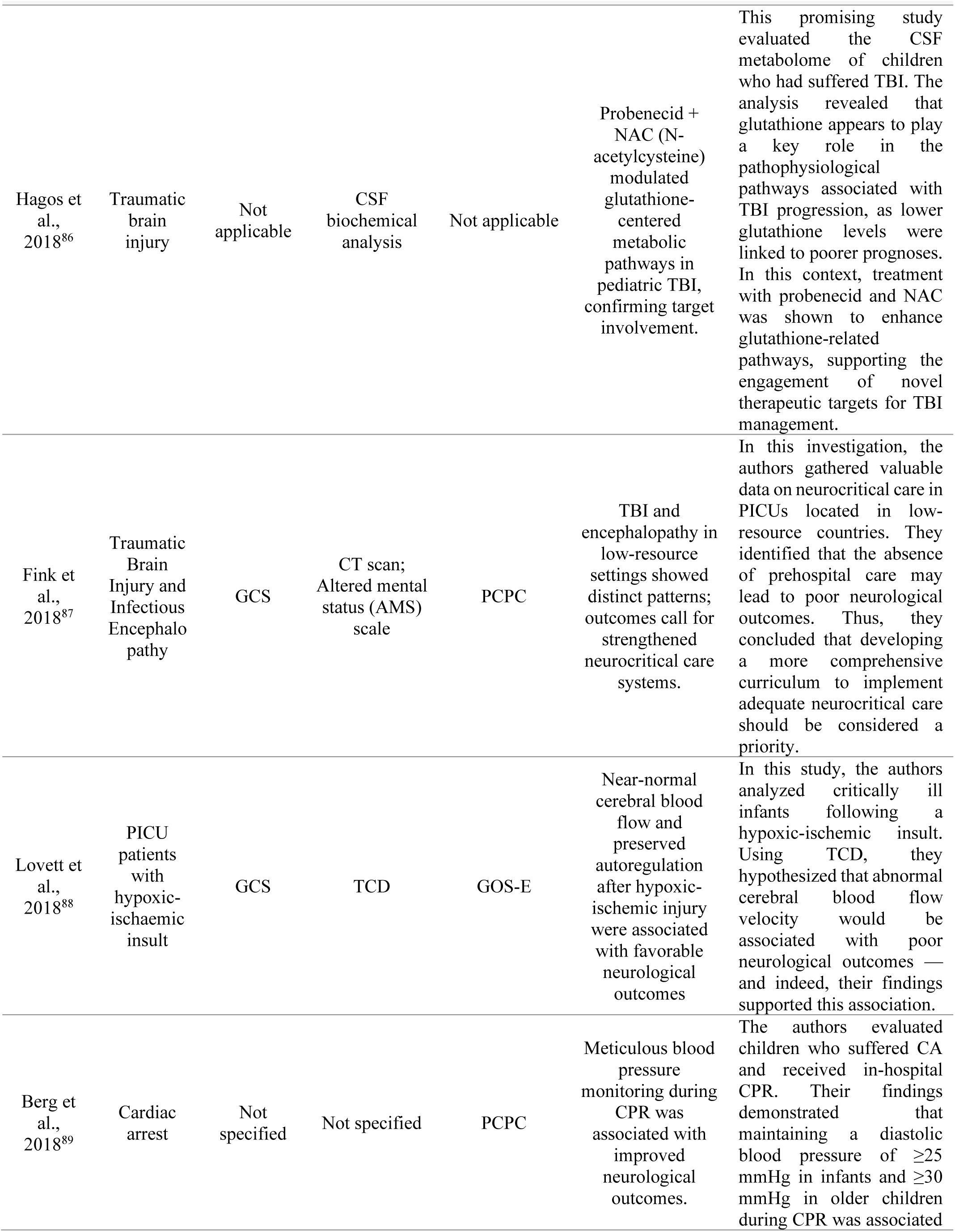
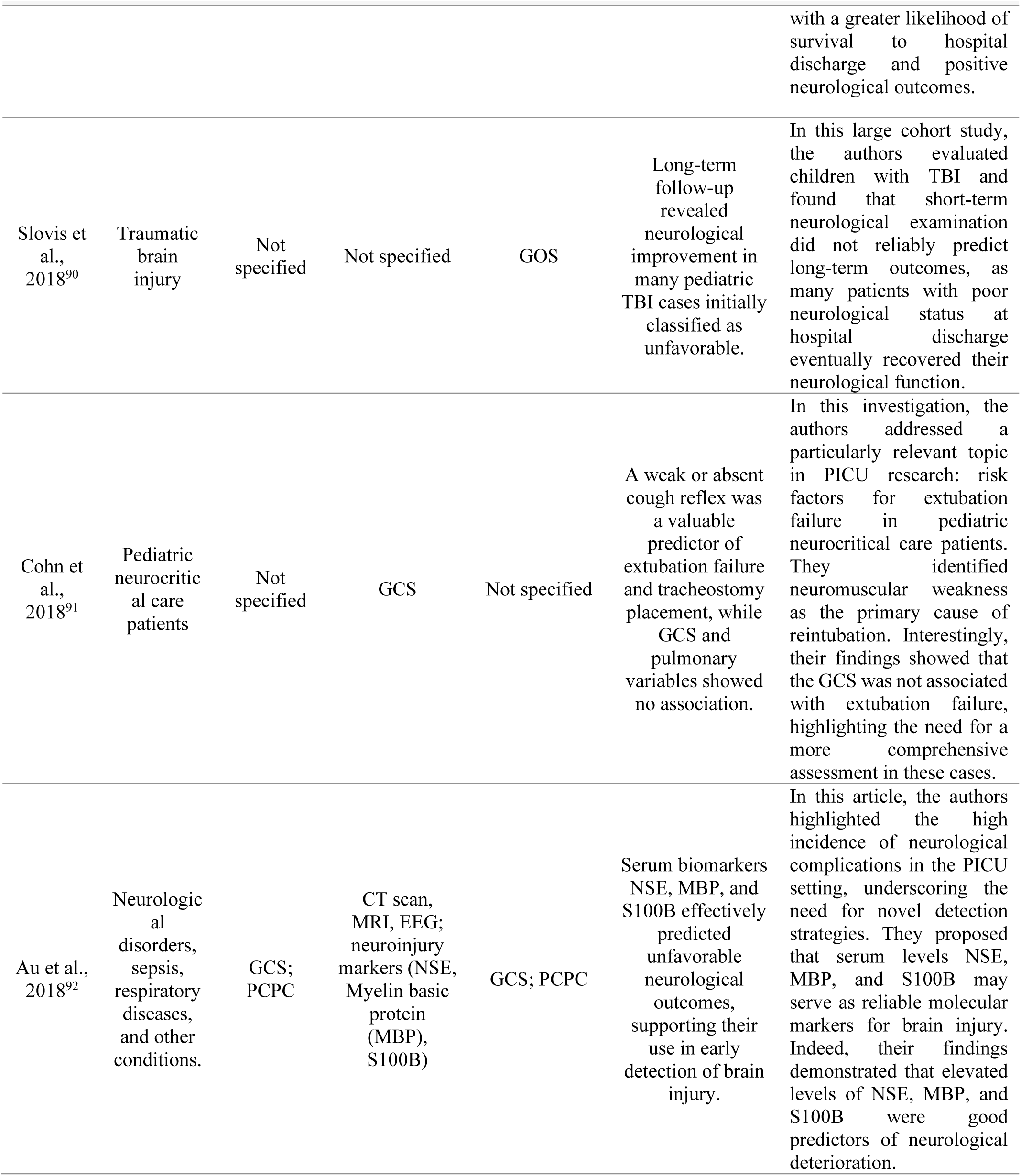
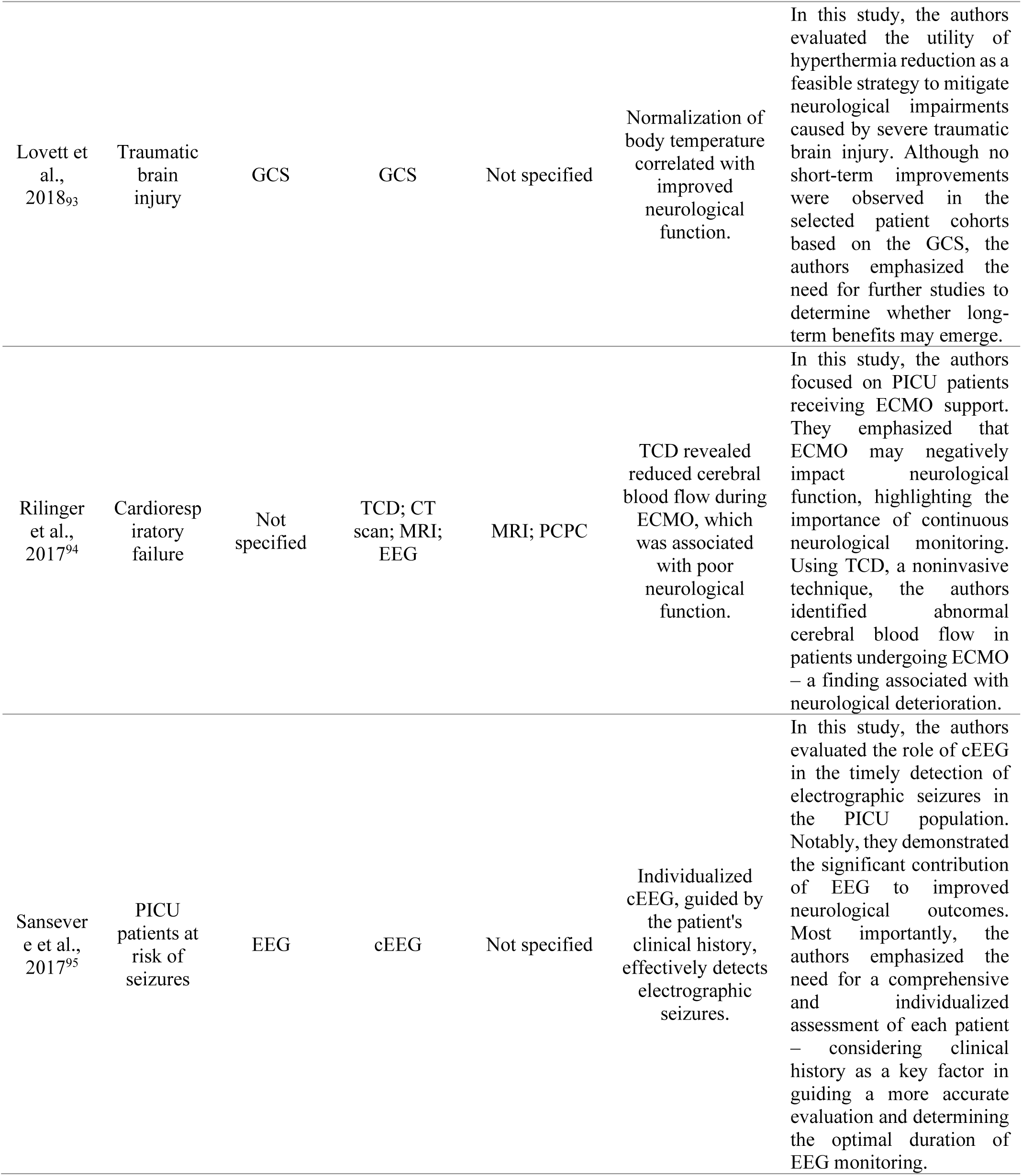
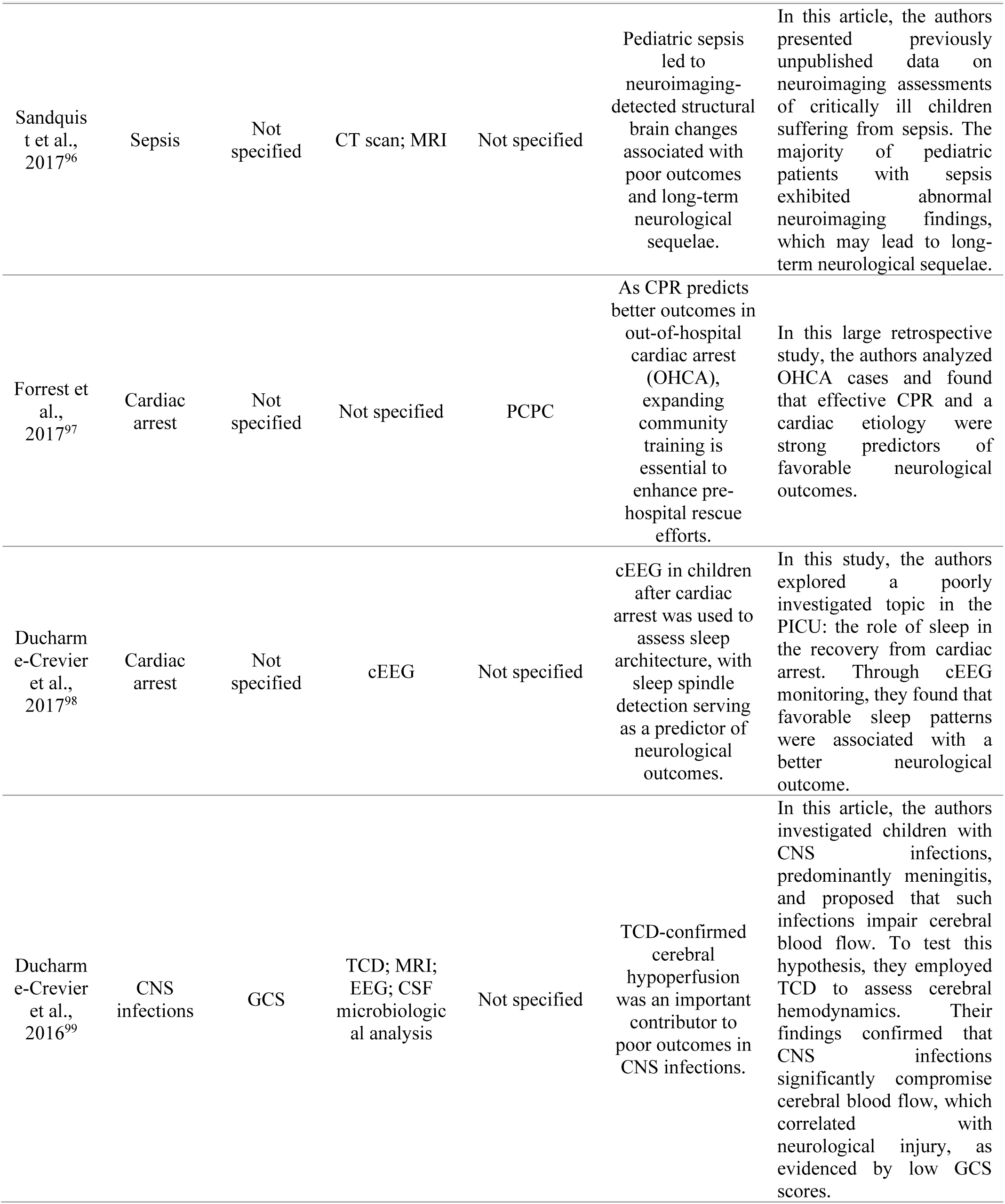
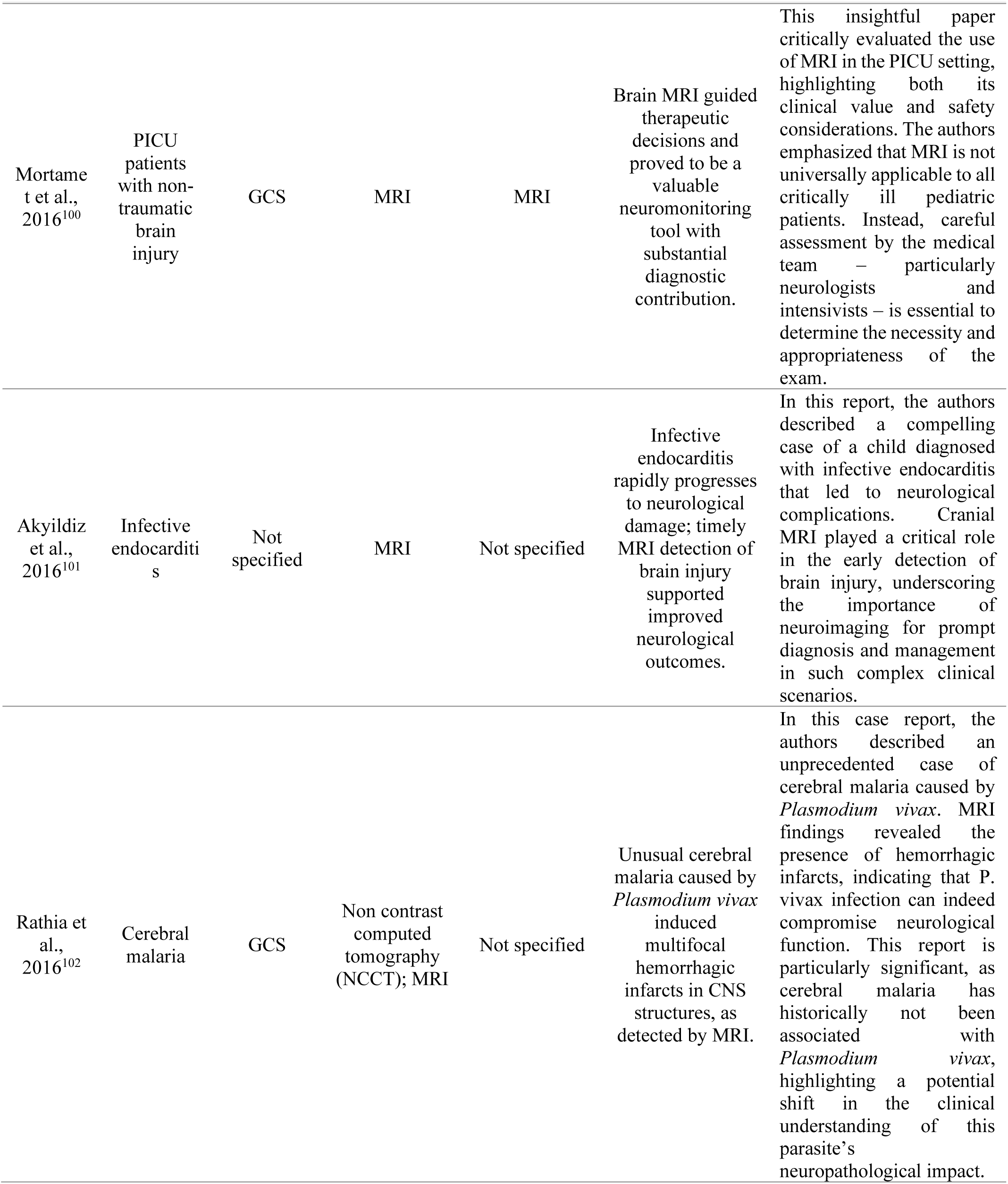
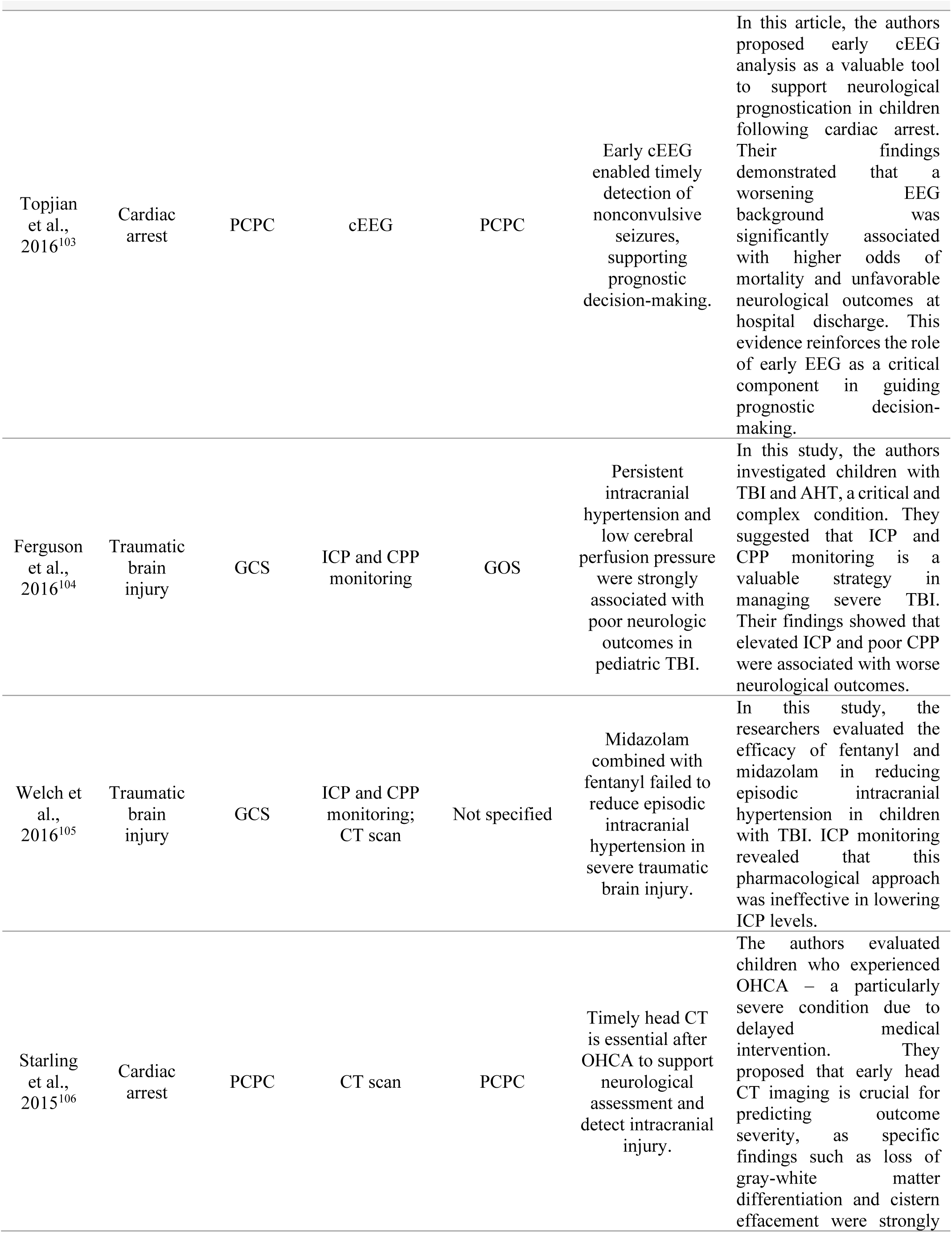
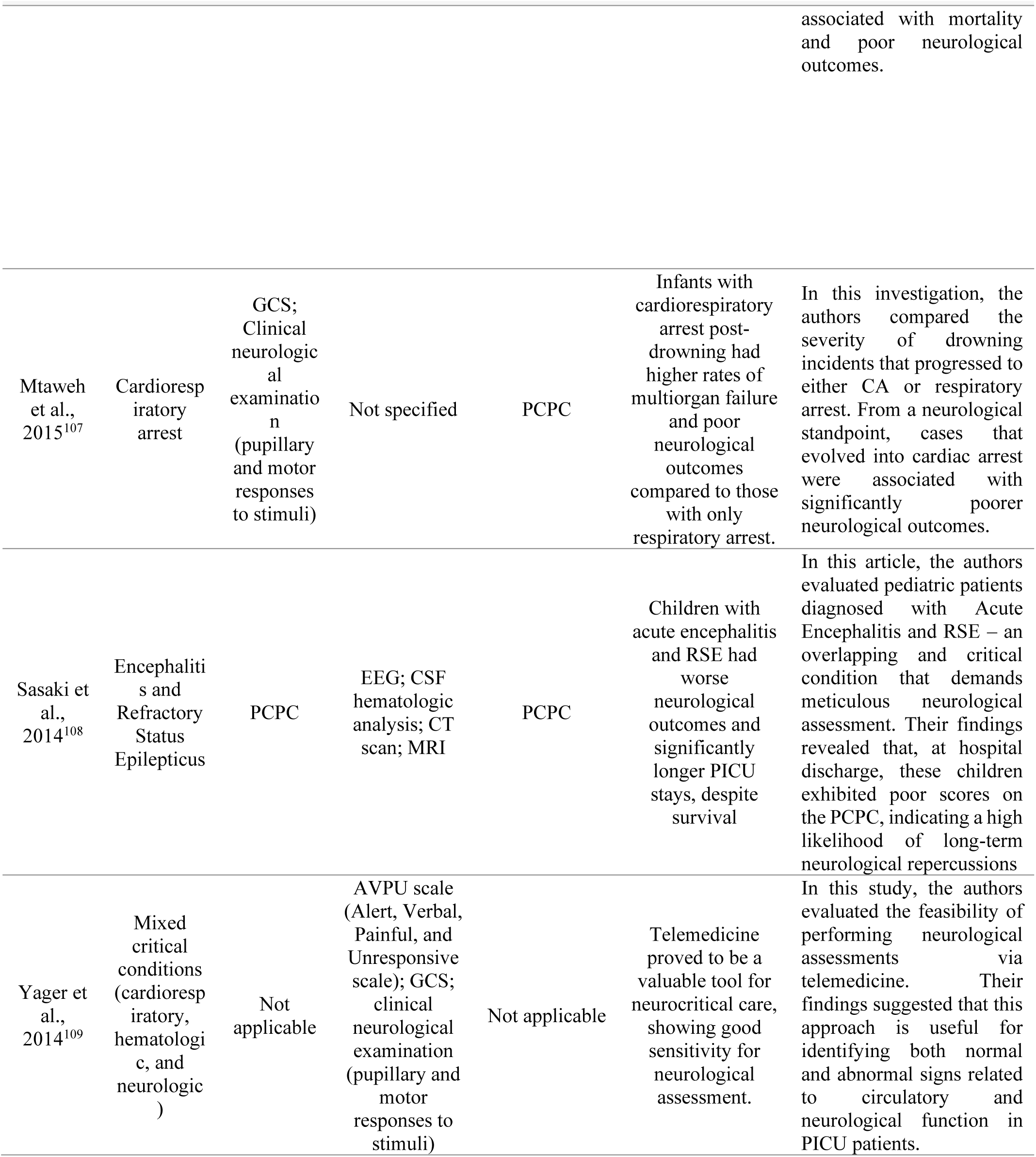
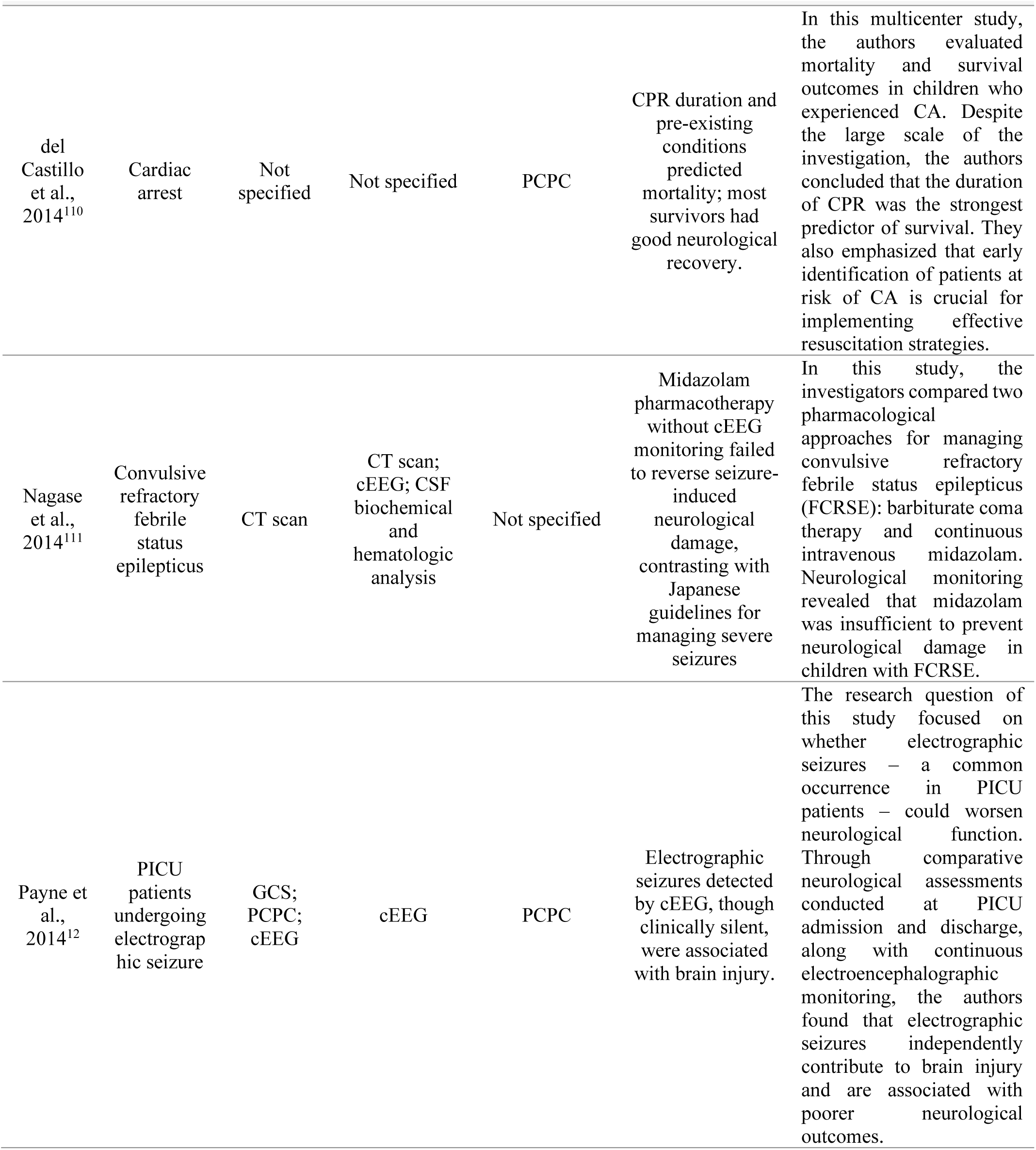
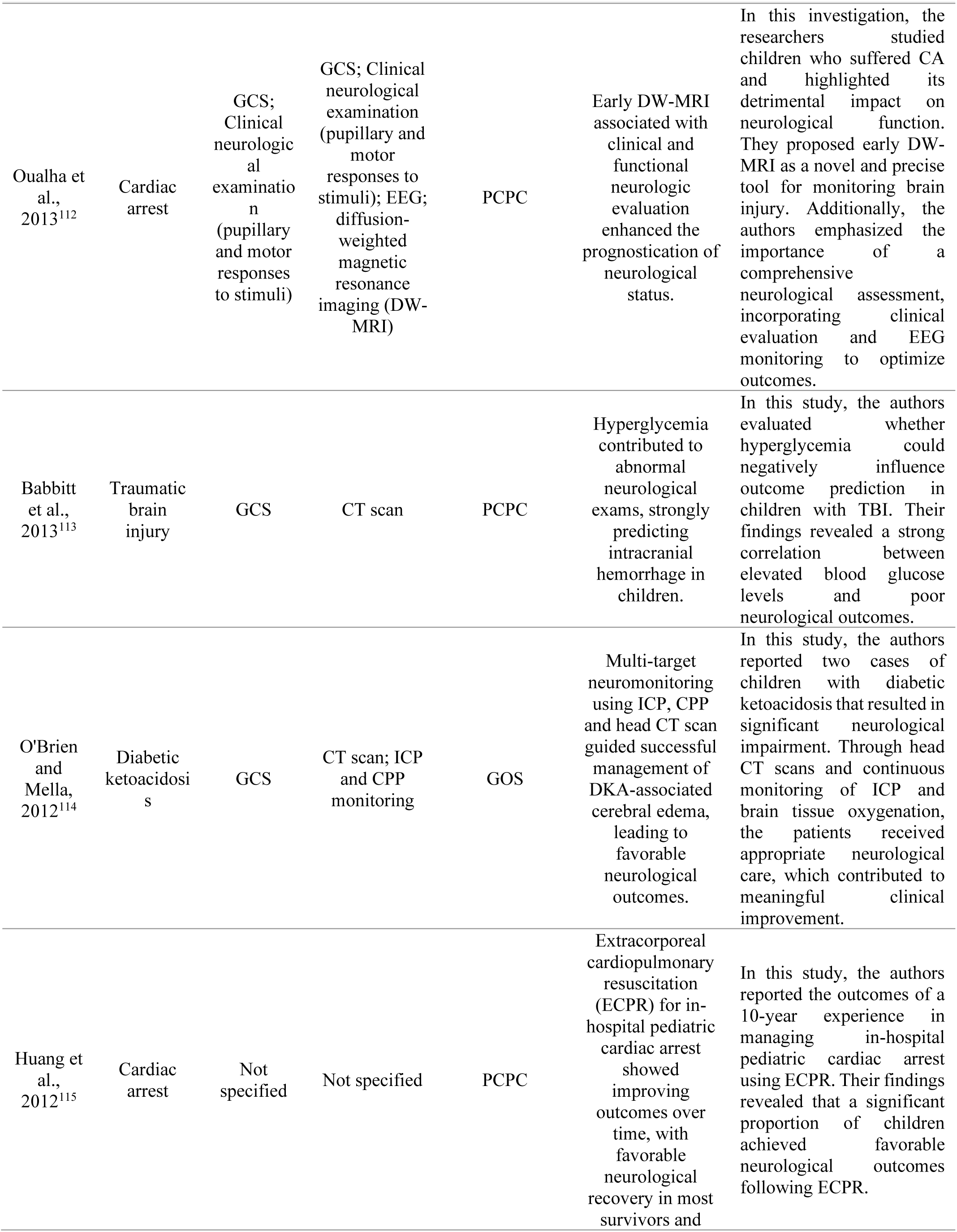
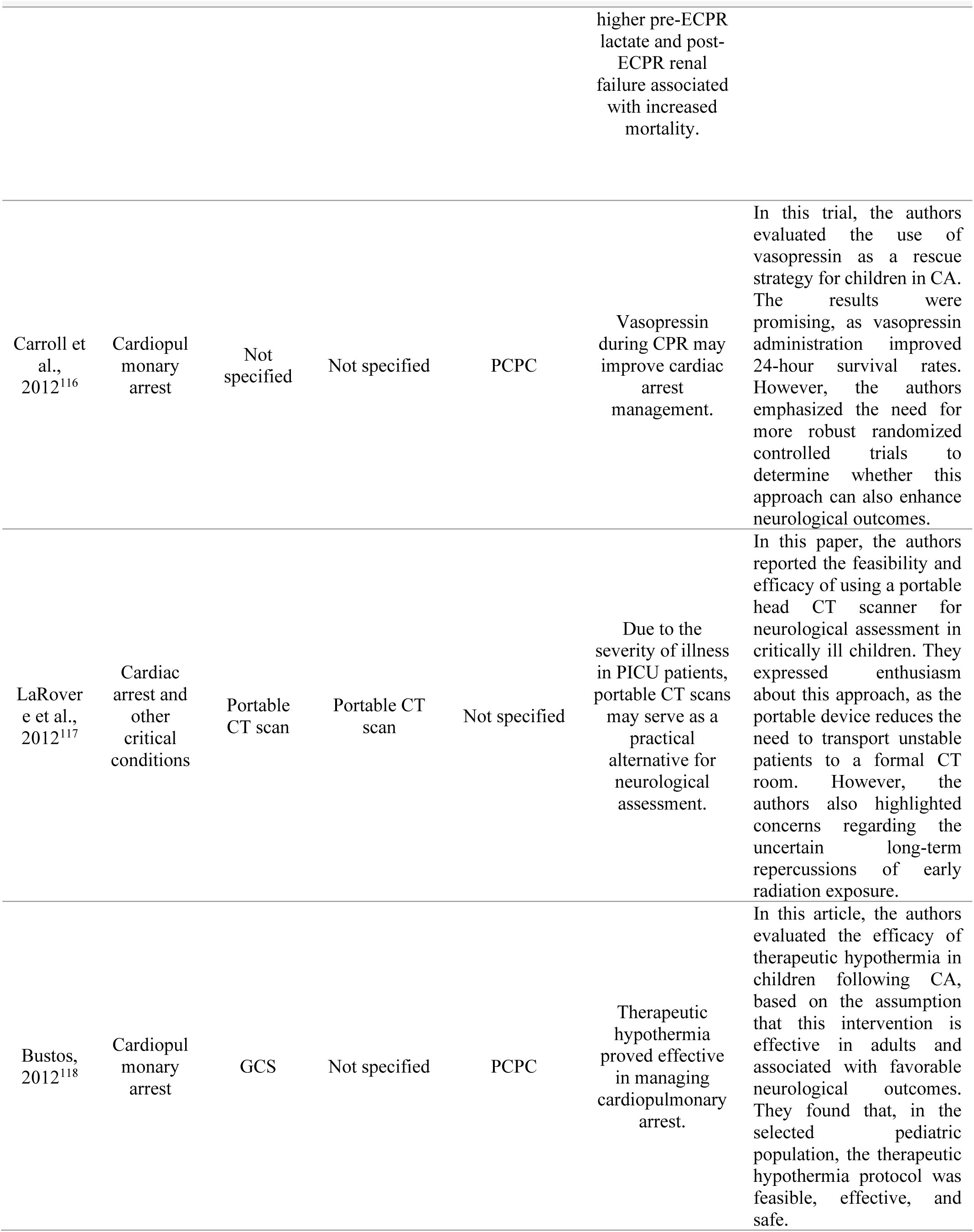
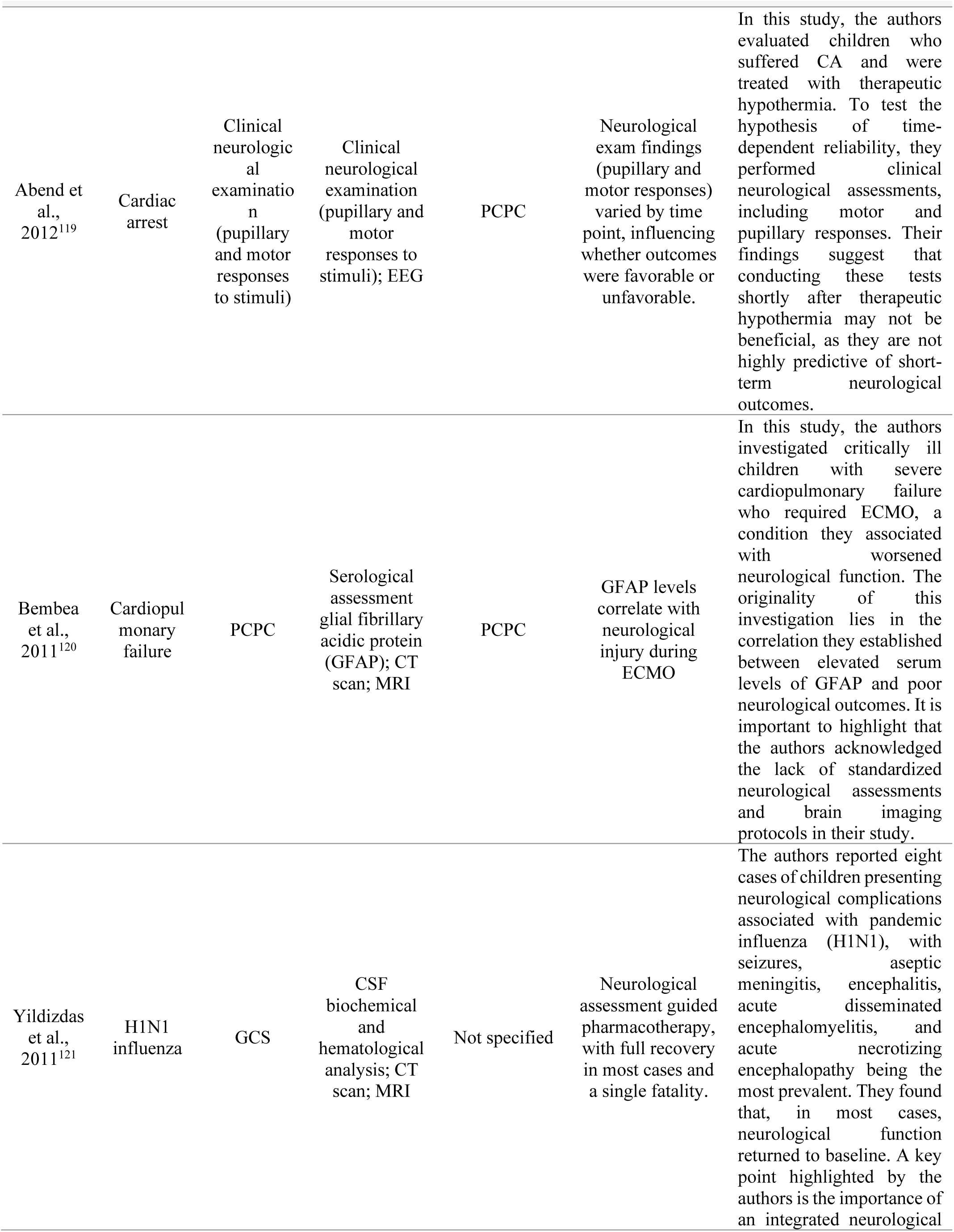
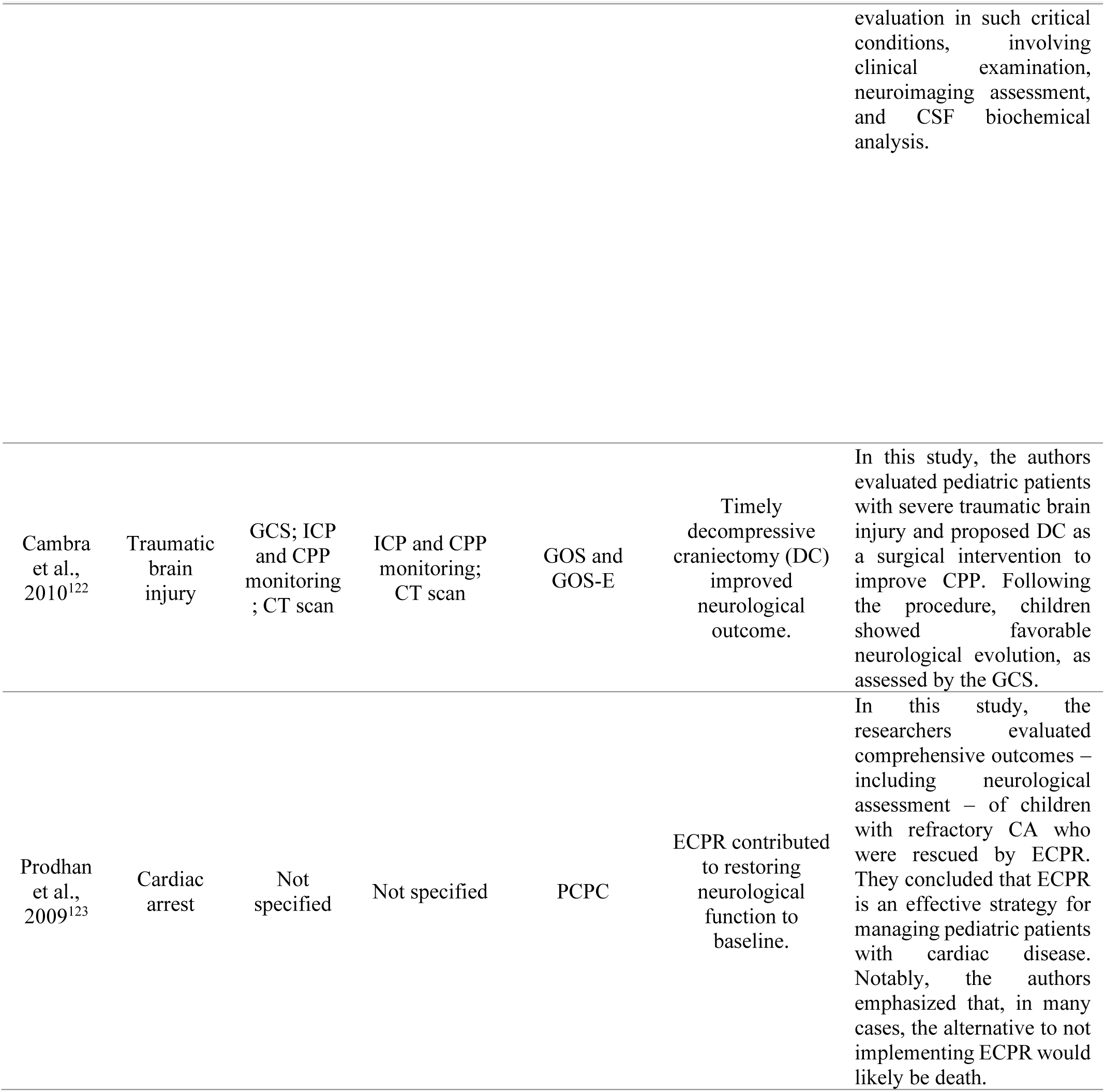
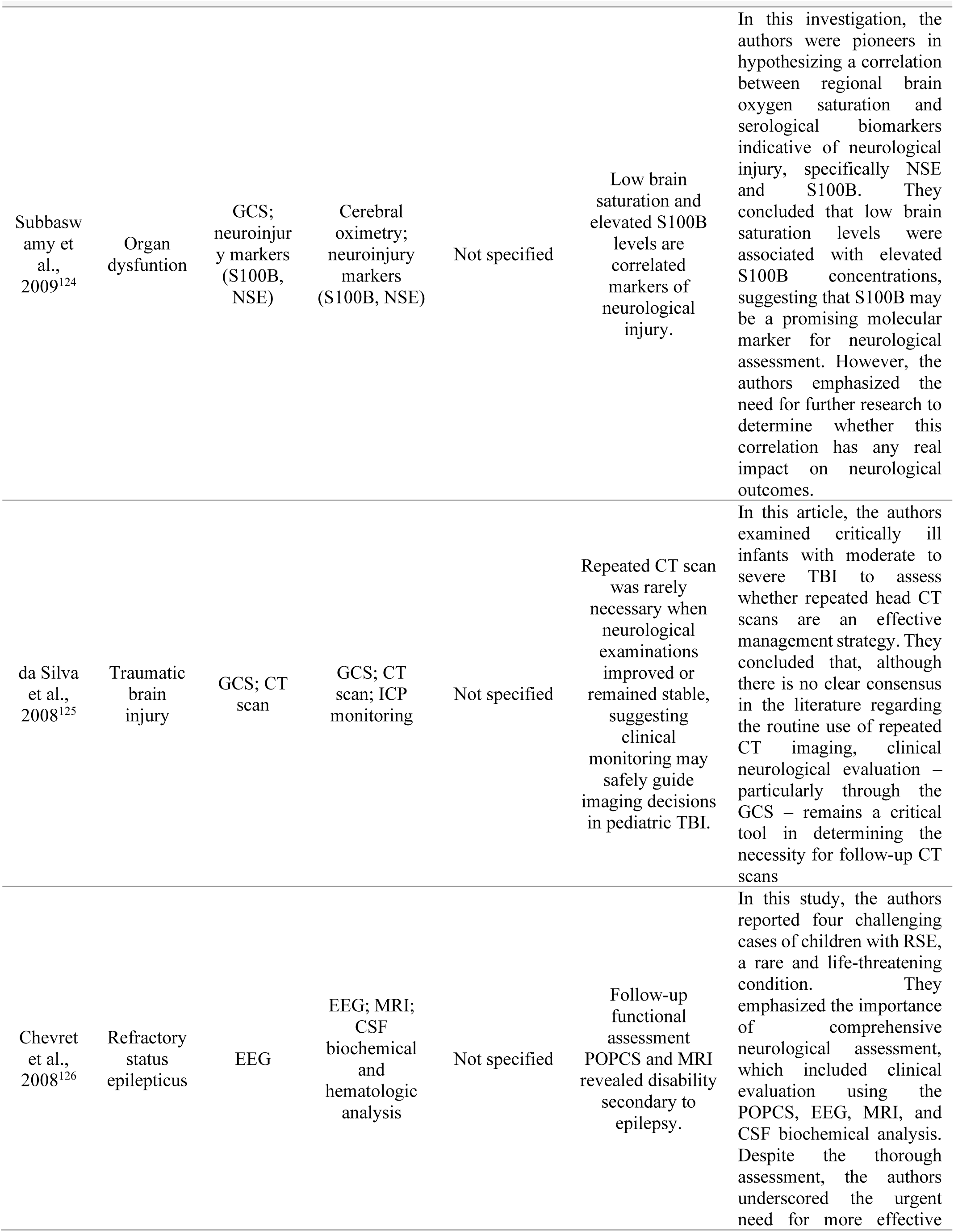
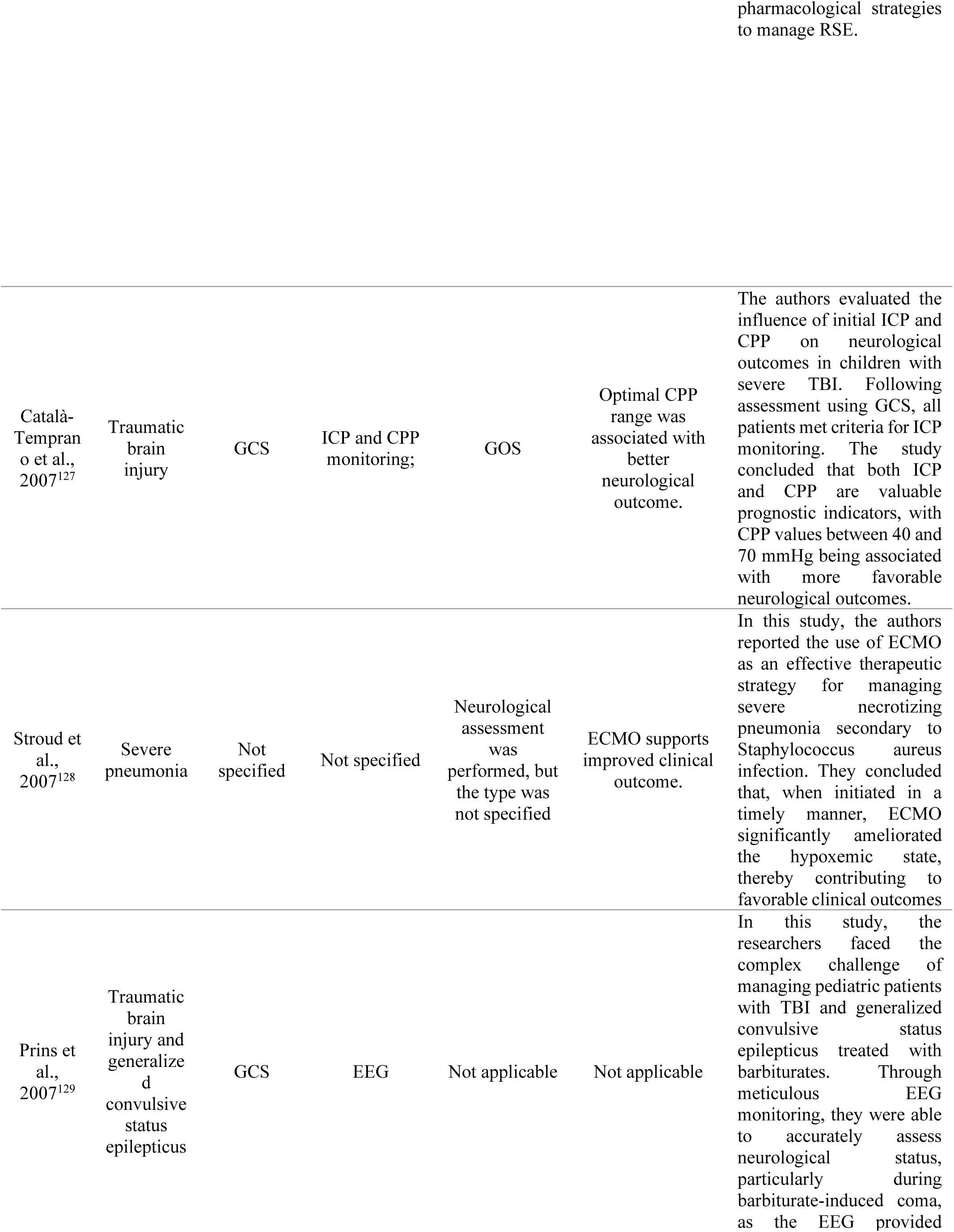
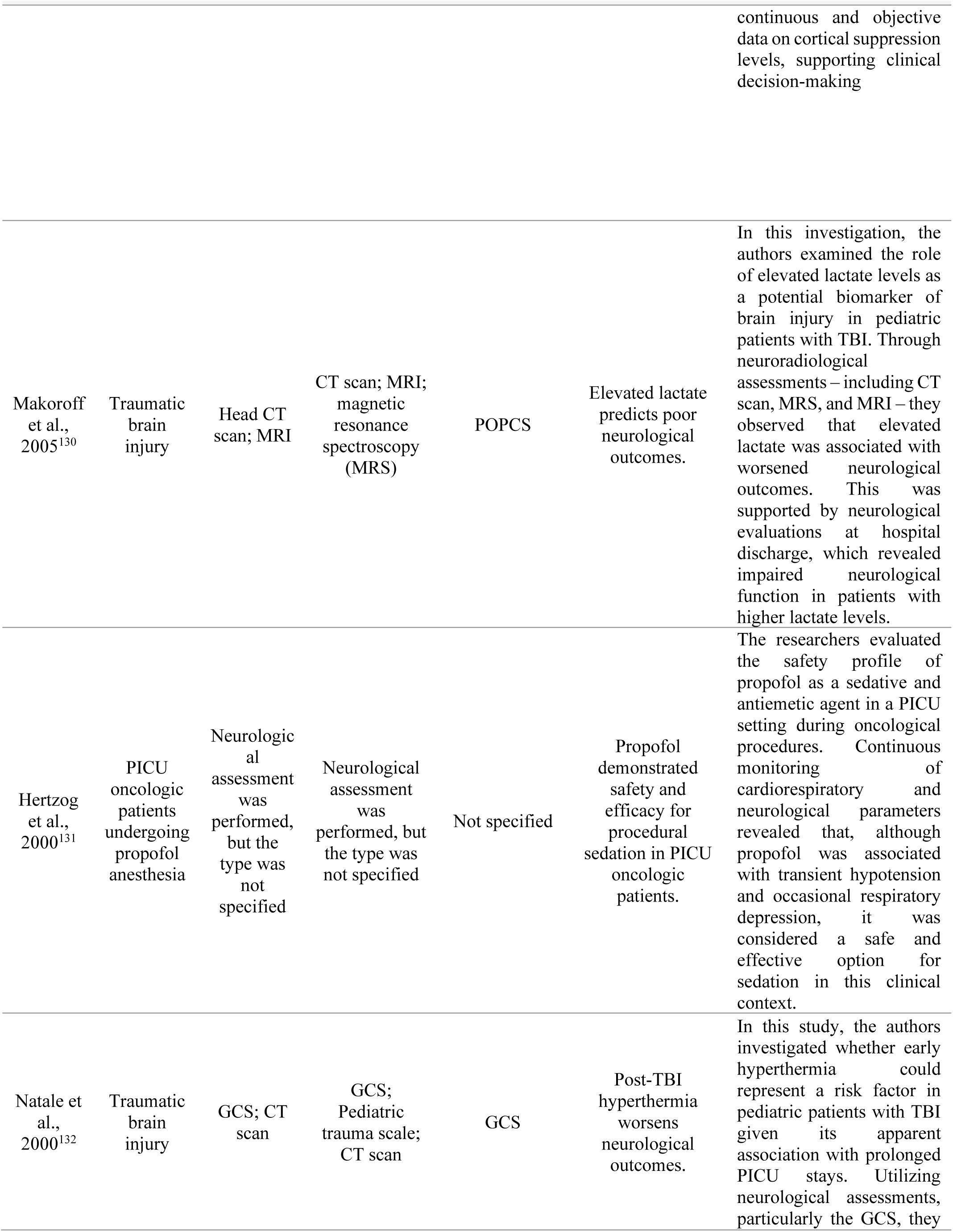
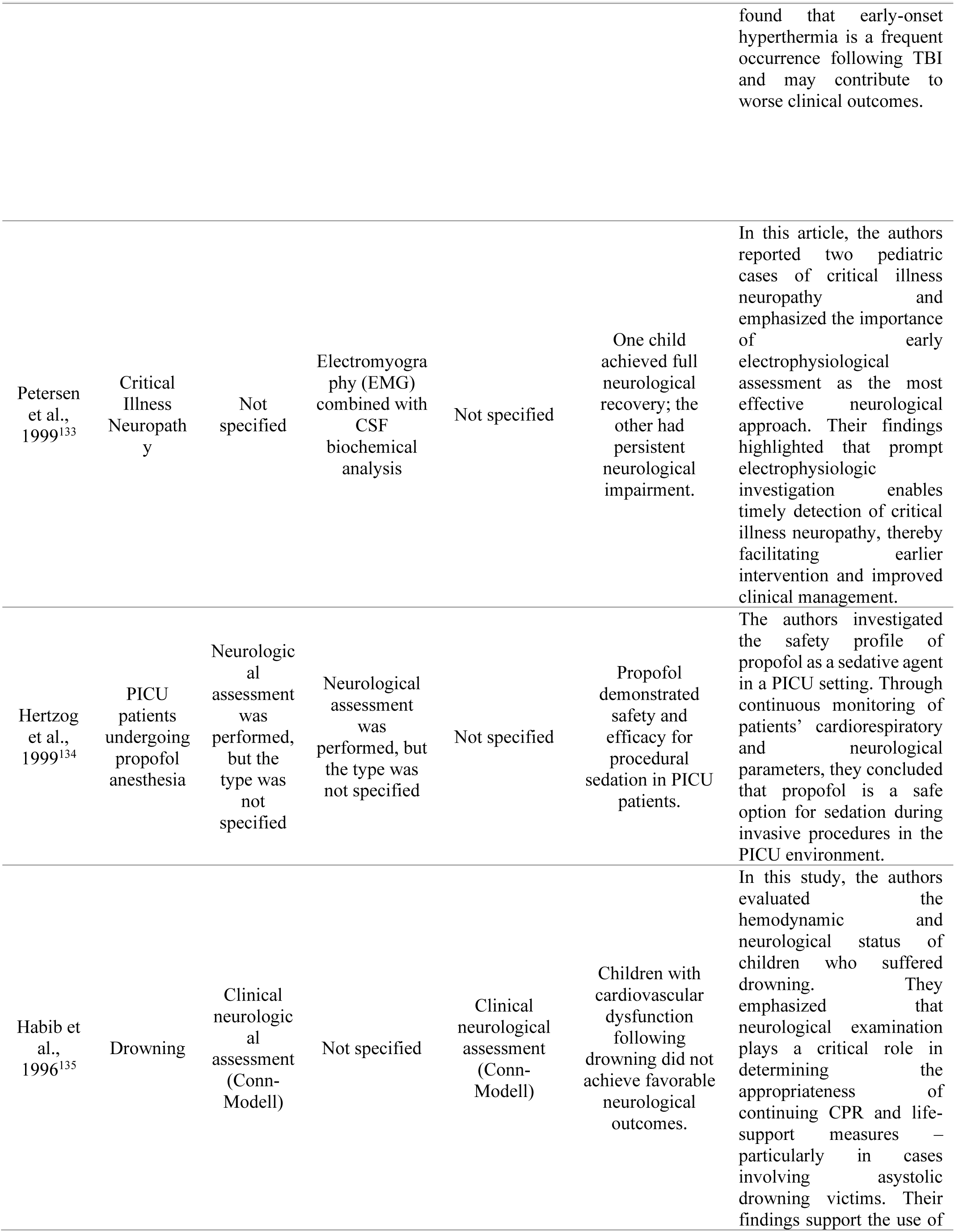
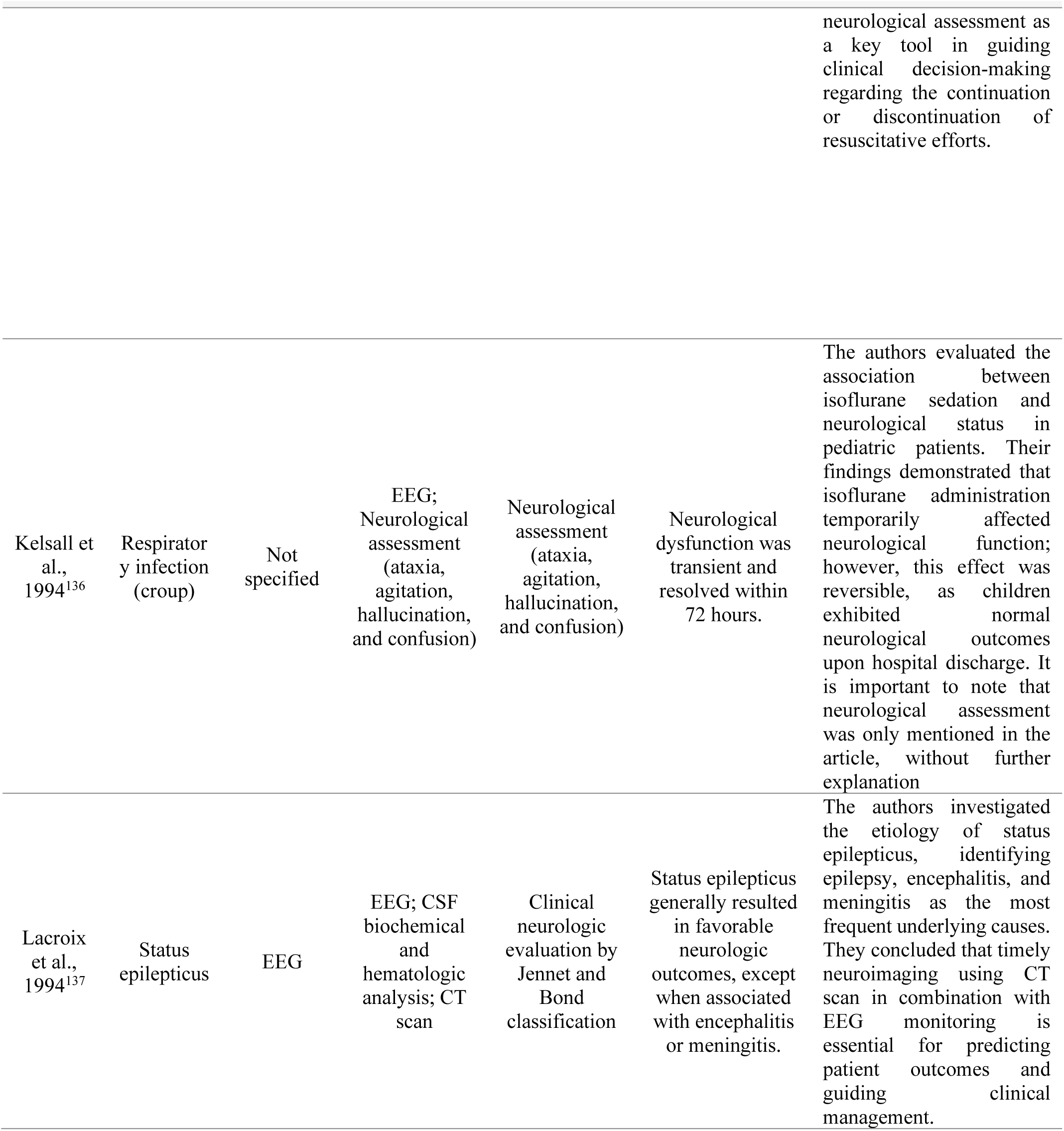
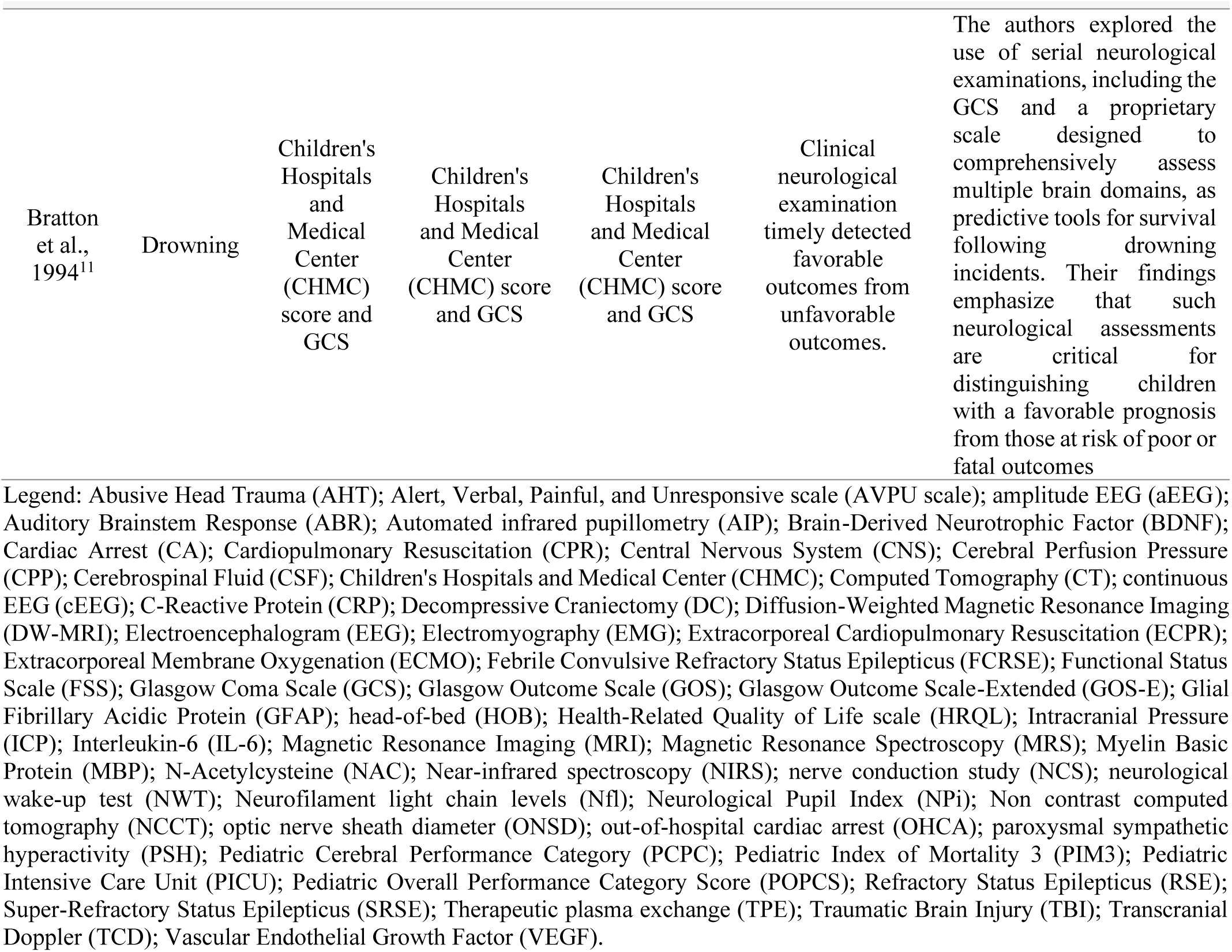
Knowledge mapping of neurological examination approaches across pediatric intensive care scenarios.

Considering the most studied diseases, the temporal distribution of publications shows a progressive increase in research output over the last two decades, with a notable acceleration in the past ten years. Traumatic brain injury and cardiac arrest dominated recent contributions, whereas CNS infections and encephalopathies appeared less frequently despite their clinical relevance (Figure 6).

**Figure 6.**
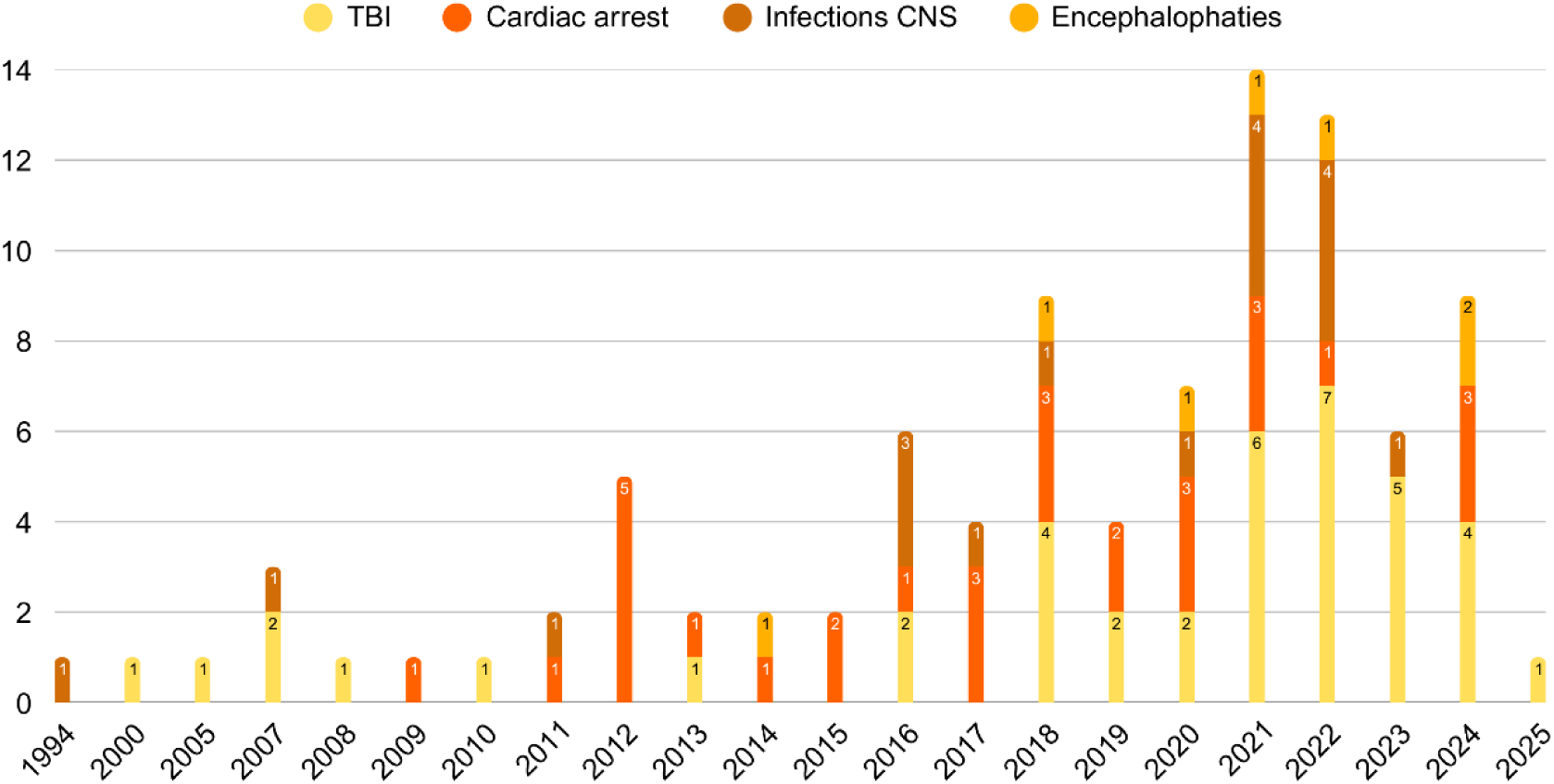
Temporal distribution of publications on neuromonitoring in critically ill children according to clinical conditions (traumatic brain injury, cardiac arrest, central nervous system infections, and encephalopathies) between 1994 and 2025. Each bar indicates the number of studies published per year. TBI = traumatic brain injury; CNS = central nervous system.

Neuromonitoring approaches across major conditions revealed disease-specific patterns. In traumatic brain injury, the GCS dominated admission assessments (n = 32), while ICP monitoring, MRI, and EEG were most frequently applied during the PICU stay. At discharge, outcome scales such as the PCPC and the Glasgow Outcome Scale-Extended (GOS-E) were commonly used, indicating a structured, multimodal approach throughout care. In contrast, cardiac arrest cases were less detailed at admission, where PCPC prevailed (n = 10), but the PICU stay relied heavily on EEG and MRI, with discharge evaluations overwhelmingly based on PCPC (n = 23). For central nervous system infections, GCS was sporadically applied at admission, but MRI (n = 11), EEG (n = 6), and cerebrospinal fluid analysis (n = 6) dominated during PICU care, whereas discharge assessments were rarely reported. Similarly, in encephalopathies, admission relied on GCS and PCPC, while MRI and EEG were most frequent during hospitalization, with limited structured evaluations at discharge (Figure 7).

**Figure 7.**
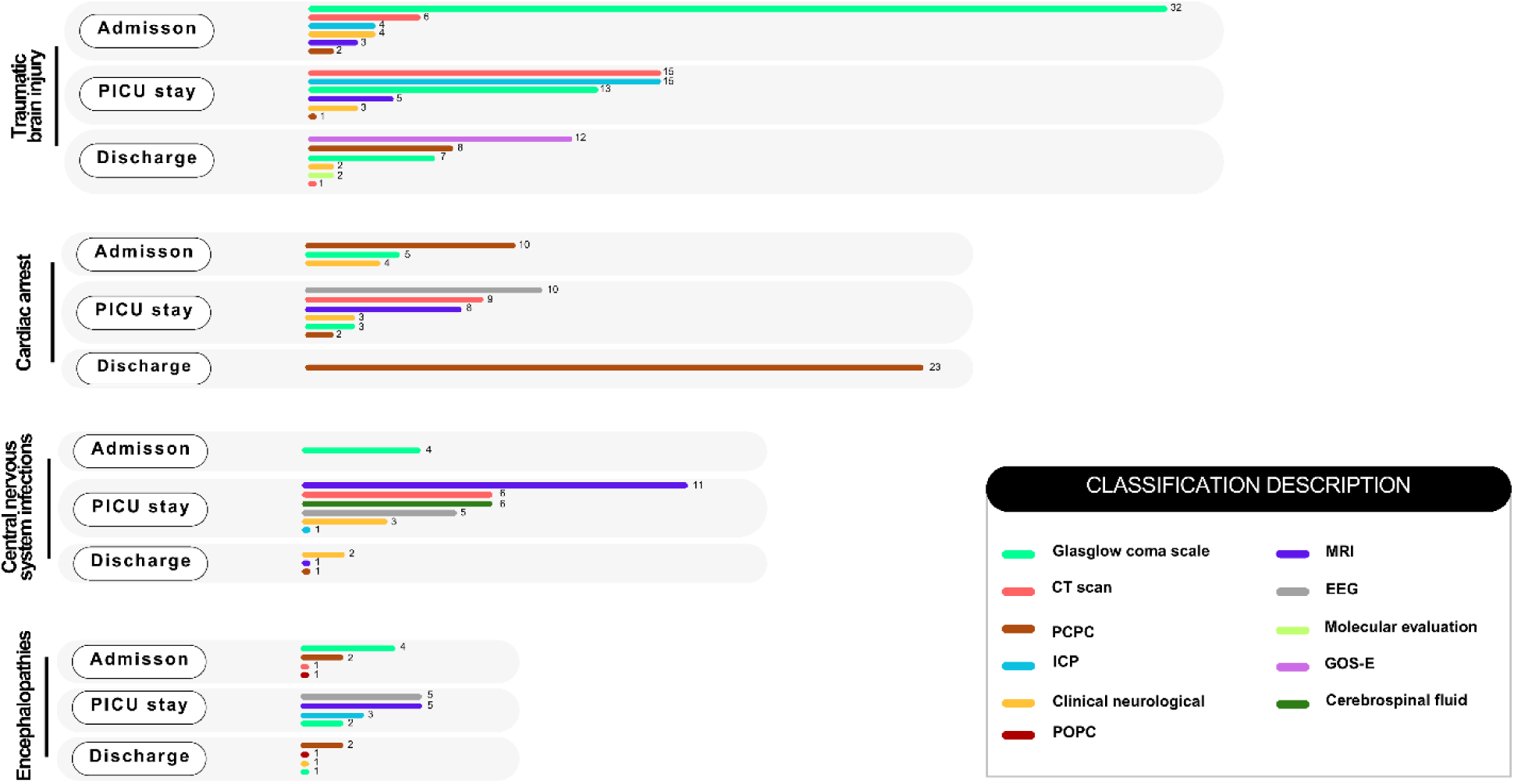
Neuromonitoring strategies reported in studies of critically ill children, stratified by condition (traumatic brain injury, cardiac arrest, central nervous system infections, and encephalopathies) and timing of evaluation (admission, PICU stay, discharge). Bars represent the frequency of each tool across studies. GCS = Glasgow Coma Scale; CT = computed tomography; PCPC = Pediatric Cerebral Performance Category; ICP = intracranial pressure monitoring; POPC = Pediatric Overall Performance Category; MRI = magnetic resonance imaging; EEG = electroencephalography; GOS-E = Glasgow Outcome Scale -Extended; CSF = cerebrospinal fluid analysis.

Biomarker evaluation was limited but promising. Only a minority of studies incorporated neuroinjury markers (n = 7), and these investigations remain underrepresented relative to imaging and electrophysiology. Reported biomarkers included neuron-specific enolase (NSE), S100B, myelin basic protein (MBP), and neurofilament light chain (NFL), with higher levels generally associated with worse short-term neurological status. In recent studies, exploratory work has also investigated glial fibrillary acidic protein (GFAP), vascular endothelial growth factor (VEGF), and brain-derived neurotrophic factor (BDNF).

## 4 Discussion

Here, we performed a comprehensive bibliometric analysis of neurological examination in critically ill infants admitted to PICUs. Our aim was to evaluate how often, and in what contexts, neurological assessments are integrated into pediatric intensive care. Despite the clinical importance of this practice, our search retrieved only 359 publications, of which 128 met the inclusion criteria. This modest number underscores the underrepresentation of neurological evaluation in the PICU literature and suggests that the topic remains undervalued in pediatric critical care research.

A striking finding was the geographic concentration of scientific output. The United States accounted for more than half of the included studies, reflecting the presence of highly specialized centers, robust funding, and established research networks. However, this predominance also raises concerns about global equity and representation. Standards developed in well-resourced contexts may not be scalable to low- and middle-income countries, potentially widening disparities in access to high-quality neurocritical care. While countries such as France, Spain, and India contributed to the literature, their citation impact was markedly lower. This pattern highlights how integration into dominant scientific networks often dictates visibility and recognition, limiting the global dissemination of knowledge generated in peripheral contexts. The imbalance in research leadership has tangible consequences. Guidelines and best practices risk have been shaped primarily by the realities of high-income settings, rather than reflecting the broader diversity of pediatric critical care worldwide. This issue highlights the need for strengthened cross-regional partnerships and greater engagement of underrepresented countries in the research landscape.

The analysis of study designs revealed a field dominated by observational methodologies, particularly retrospective and prospective cohort studies, which together represented more than half of the literature. While these designs are valuable for describing clinical practices and associations, they provide limited evidence for establishing causality or informing standardized neurological protocols. The scarcity of randomized clinical trials and interventional studies demonstrates the lack of high-level evidence to guide clinical decision-making in this domain. Furthermore, inconsistencies in how study types were reported, often relying on author-defined classifications, point to an absence of methodological consensus. This heterogeneity undermines comparability and hampers critical synthesis of results. Reliance on retrospective designs is especially problematic, as it increases susceptibility to selection bias, underreporting of outcomes, and insufficient follow-up on long-term neurodevelopmental sequelae. These limitations underscore the urgent need for prospective, multicenter, methodologically rigorous studies capable of validating assessment tools and capturing the longitudinal impact of critical illness on brain development. Patterns of author productivity followed Lotka’s Law, whereby a small number of prolific authors produce the majority of publications, while most contribute only occasionally. The Berg-Nadkarni-Topjian network emerged as a central hub, shaping much of the research in this field. At the same time, mapping of corresponding authorship highlighted additional leaders, such as Fink EL, Lang SS, Appavu B, Au AK, and Kirschen MP, whose influence extends beyond publication volume through consistent roles in study coordination and editorial responsibility. Notably, many of these authors are based in North American institutions, again emphasizing the geographic concentration of intellectual leadership. While such concentration fosters expertise and standardization, it also raises concerns about inclusivity and the dissemination of context-sensitive care practices across diverse healthcare systems.

Keyword co-occurrence patterns further illustrate the field’s priorities. The prominence of terms such as “traumatic brain injury,” “cardiac arrest,” “neuromonitoring,” and “outcome” highlights a predominant focus on acute neurological emergencies and prognostication. The frequent recurrence of “neurocritical care” and “cardiopulmonary resuscitation” reflects increasing integration of multidisciplinary approaches and the gradual institutionalization of specialized services within PICUs. Together, these trends indicate that researchers are not only concerned with identifying neurological impairment but also with developing tools to monitor, predict, and improve long-term outcomes in critically ill children.

Journal distribution adhered to Bradford’s Law of Scattering, with a small core of journals, such as Pediatric Critical Care Medicine, Neurocritical Care, and Resuscitation, accounting for more than a third of publications. These journals represent the primary venues for disseminating research in this area and highlight the centrality of specialized outlets in consolidating knowledge. At the same time, high-impact interdisciplinary journals, such as Circulation, Brain, and Critical Care, hosted some of the most influential articles, underscoring the translational importance of pediatric neurocritical care beyond the specialized niche. Conversely, the wide dispersion of single-publication journals reflects the emerging and multidisciplinary character of this field, including expertise on pediatrics, neurology, neurosurgery, emergency medicine, and intensive care. While this fragmentation complicates the consolidation of evidence, it also highlights opportunities for cross-disciplinary integration, particularly as pediatric neurocritical care continues to emerge as a distinct subspecialty.

In summary, our bibliometric findings reveal a field that is growing but still limited in scope, geographically imbalanced, and methodologically constrained. Neurological examination in the PICU remains largely underreported in literature, concentrated in a few countries and institutions, and based on observational research.

While our study is primarily bibliometric, we also conducted a mapping of critical knowledge, focusing on the reasons for PICU admission and the modalities and timing of neurological assessments. Among the identified causes, traumatic brain injury, cardiopulmonary complications, encephalopathies, and infections emerged as the most frequent entry diagnoses, followed by a heterogeneous group of other conditions. This narrow range of conditions retrieved prompts a tentative key conclusion that neurological examination is disproportionately explored in select pathologies directly affecting the central nervous system. The data reinforces the notion that neurological assessment across the broader PICU patient population remains an underrepresented and overlooked area in scientific literature.

As expected, traumatic brain injury represents the most common cause of PICU admission, reflecting acute damage of central nervous system function^21^. It is important to highlight that traumatic brain injury represents a highly sensitive condition in the pediatric population, considering the critical period of ongoing neurodevelopment during early childhood^25^. Accordingly, several international guidelines have been established to promote optimal care for pediatric traumatic brain injury, offering structured recommendations for both clinical management and research directions^138,139^. These frameworks served as a benchmark for interpreting our findings, enabling a comparative and critical analysis.

Surprisingly, fewer than half of the studies incorporated ICP monitoring, a key recommendation in international guidelines for the management of moderate to severe traumatic brain injury^22, 29, 44, 57, 66, 73^. This finding suggests a concerning deviation from established neurocritical care standards. On the other hand, despite receiving comparatively less emphasis in protocols, clinical neurological assessment played a central role in evaluating traumatic brain injury severity. Tools such as the GCS, with particular focus on pupillary responses, and assessments of consciousness level were widely applied, especially during initial stabilization in the emergency setting^25, 28, 31, 57^.

To enhance clarity and structure in the critical synthesis, we mapped neurological assessments according to their timing: (i) at admission, (ii) during PICU stay, and (iii) at discharge. This strategy facilitated the identification of trends and gaps across the patient care continuum. For traumatic brain injury, most studies reported initial neurological evaluations upon admission, mainly using the GCS^21,22,23,29,25,27,28^. In contrast, evaluations conducted during PICU stay and especially at discharge were less frequently described^23,53,49,73,81,90^. Notably, when assessments were conducted at discharge, the PCPC scale was the most frequently used tool, offering valuable insights into functional outcomes and persistent cognitive sequelae23,31,30,42,113.

Contrary to expectations, neuroimaging modalities, such as CT and MRI, were not consistently employed across studies. Furthermore, the integration of ICP monitoring and neuroimaging that are widely recognized as the gold standard for pediatric neurocritical care, was poorly reported with solely seven articles^25, 31, 36, 66, 105, 122, 125^. This finding illustrates a lack of standardization in neurological monitoring and suggests a potential underuse of available technologies to guide clinical decisions in traumatic brain injury care.

Another clinical scenario frequently addressed in the included studies involved cardiopulmonary complications, particularly cardiac arrest. At admission, a concerning pattern emerged, which many studies failed to specify the exact clinical neurological evaluation performed^79, 89, 97, 98, 110, 115, 123^. This occurrence may reflect the fact that neurological impairment was often secondary to systemic events, such as hypoxia or circulatory collapse^14^. In the subset of studies that reported neurological assessment at admission, the PCPC scale was the most frequently used tool^14,15,18,62,71,74^. Given that cardiac arrest typically results in global hypoxic-ischemic injury rather than focal neurological deficits, the use of the broad PCPC scale is justified^14^. During the PICU stay, a large proportion of studies either did not specify the neurological approach or emphasized functional monitoring tools, particularly EEG, alongside neuroimaging techniques (CT scan and MRI)^61,62,63,82,112^. At discharge, many studies again omitted details of neurological assessment^61,63,82,85,98,117^. However, among those that reported such evaluations, more than half used the PCPC scale, highlighting an emphasis on functional status and survival quality, rather than real-time neurological monitoring throughout the critical care course^14,15,18,41,117,71,74^.

Central nervous system infections, another significant cause of PICU admission, displayed a similar lack of standardized neurological evaluation at both admission and discharge^40,51,55,96,101^. Most assessments were concentrated during the PICU stay, when patients are typically under closer monitoring. The most commonly employed tools were neuroimaging techniques, such as MRI and CT scans, used to identify structural changes or complications^44,46,54,55,64,67,84^. Functional evaluation with EEG was also reported, though less frequently^40,67,84,92,99^. Strikingly, cerebrospinal fluid analysis, which should be a cornerstone in the diagnostic workup of infectious central nervous system diseases, was poorly reported in less than half of the studies^44,55,64,84,99,121^. This omission raises important questions regarding the completeness and standardization of diagnostic strategies in pediatric critically ill patients.

Though less frequent, encephalopathies also emerged as relevant causes of PICU admission in our dataset. At admission, most studies specified the neurological approach, with GCS and PCPC being the tools of choice^13,24,43,69,108^. During the PICU stay, EEG and MRI were commonly used, suggesting greater emphasis on identifying subclinical seizures or metabolic abnormalities^13,24,108^. Notably, ICP or CPP monitoring was rarely used in encephalopathy cases, despite its potential relevance in conditions associated with cerebral edema^13,24,69^. At discharge, most studies once again failed to specify whether a neurological evaluation was performed, reflecting a broader tendency to prioritize acute management over long-term outcome evaluation^24,39,45,47,59^.

One of the most promising domains, albeit still emerging, is the use of neurological biomarkers. Few studies in our survey explored the application of markers such as S100B, NSE, and GFAP^26,58,124,71,92,120^, which were frequently associated with structural injury, neuroinflammation, or poor outcomes. For instance, serum GFAP and NSE correlated with functional outcomes and could represent early indicators of diffuse injury, even in children without evident focal deficits^124,120,92^. However, the limited number of studies employing biomarkers reflects an insufficiently explored opportunity to advance toward objective, reproducible, and early-detection strategies in pediatric neurocritical care and precision medicine. This is of particular relevance in view of the growing international effort to standardize biological markers panels and validate their clinical applicability.

Thus, our findings reveal that neuromonitoring in pediatric critical care remains concentrated on a few acute neurological emergencies, applied at selected timepoints, and heavily reliant on established tools, such as GCS, CT, EEG, and ICP/CPP. While early findings are promising, biomarker research remains poorly explored. This selective focus highlights both progress in specific domains and substantial gaps, notably the underrepresentation of systematic, multimodal surveillance across the wider PICU population. Over the past decades, neuromonitoring in pediatric intensive care settings has gained growing attention. One of the leading contributors to this field is Dr. LaRovere, whose group has published pivotal studies that robustly contribute to the emerging landscape of pediatric neurocritical care. Notably, a nationwide survey of 328 American institutions revealed that only 45 institutions met the criteria to be considered as neurocritical care centers^140^. Essentially, the authors highlighted a major obstacle to advancing the field, which consists in the absence of a standardized, universally accepted definition of what constitutes a neuromonitoring service. This concern is not isolated. Chang and Rasmussen^141^ discussed the lack of standardization in neuromonitoring protocols in the PICU. Through a retrospective chart review at a single institution, the authors observed that while neuromonitoring was more consistently applied in adult intensive care units, pediatric cases lacked formal protocols and consensus guidelines. Although such surveys were institution-specific, we hypothesized that this reality could reflect a broader international scenario. Our bibliometric approach offers a pioneering contribution by gathering and analyzing global publications, consolidating the available evidence to demonstrate that this is a gap that requires critical attention.

Motivated by this gap, our research group has been investigating the neuropsychiatric consequences of psychotropic drugs in the PICU environment^142,8^, to bring greater scientific attention to the current scenario of pediatric neurocritical care through a comprehensive bibliometric and knowledge-mapping analysis. While most of the existing literature on pediatric neurocritical care focuses on clearly defined neurological conditions, we argue that PICU admission *per se,* regardless of the primary diagnosis, may be considered a potential injury to the central nervous system. This perspective is particularly relevant given the routine use of sedoanalgesia and other invasive procedures, which may lead to long-term neurocognitive repercussions in children^143,8^.

It is important to highlight that our bibliometric review did not aim to evaluate the methodological rigor or quality of individual studies, but rather to synthesize the current state of the literature, identify gaps, and highlight directions for future research. This goal was largely accomplished, though several limitations must be acknowledged. Firstly, the analysis was restricted to a single database (e.g., WoS-CC), which, although widely recognized and comprehensive, may have excluded relevant publications indexed elsewhere. Second, the effectiveness of bibliometric approaches is inherently constrained by how research is framed, labeled, and reported by authors. Neurological assessments are likely performed routinely in PICUs worldwide; however, they are often embedded within broader clinical protocols and seldom reported as standalone outcomes. This underreporting may reflect a perception that such data hold limited scientific novelty or impact, thereby contributing to their relative absence in indexed literature. Thus, our findings should not be interpreted as evidence that neurological evaluation is underused in clinical practice, but rather that it is underrepresented in research publications. Recognizing and addressing this publication gap is essential to ensure that the cumulative scientific record accurately reflects the true scope of clinical practice and informs evidence-based standards of pediatric neurocritical care.

Finally, it is worth noting that delirium, despite its growing recognition as a critical neurological manifestation in critically ill children, was not retrieved in our search. Delirium has been described by many researchers as a *new vital sign,* representing a clinical expression of acute brain dysfunction^144^. Its absence in our dataset underscores the persistent underdiagnosis and underreporting of this neuropsychiatric emergency^145^. Moreover, the inconsistent use of terminology remains a significant barrier, as emphasized by a consensus statement published in 2020 that unified the preferred nomenclature for delirium and acute encephalopathy, endorsed by ten professional societies^146,147^. That declaration highlighted the need for a standardized lexicon across disciplines, emphasizing that delirium – unlike vague terms such as “altered mental status” – is a valid, operationalized diagnostic construct with high reliability and clinical utility. Recognizing and naming delirium explicitly not only enables structured management approaches but also fosters transparent communication with patients and families, a vital step in mitigating the human and ethical burden of neurological dysfunction in pediatric intensive care.

## Conclusion

Together, our findings reveal a significant disconnection between the complex neurological vulnerabilities of critically ill children and the current scientific strategies guiding their care. The predominance of low-evidence study designs and the selective application of neuromonitoring technologies point to a fragmented approach that hinders the development of effective and equitable pediatric neurocritical care. Addressing this gap requires not only more robust and multicenter methodologies, but also a conceptual shift in which every PICU admission should be considered a potential neurological risk. By adopting this broader perspective, clinicians and researchers can begin to design inclusive, predictive, and restorative models of care that better safeguard long-term brain health.

Our central conclusion relies on the scarce literature explicitly addressing neurological examinations in critically ill children. This gap raises important questions as to whether neurological assessments are underutilized in clinical practice or simply underreported and deprioritized in scientific literature. Regardless of the cause, our analysis highlights an urgent need to broaden the scope of neurological evaluations beyond children with overt neurological diagnoses. Standardized, timely, and structured neurological support should become a routine component of PICU care, recognizing the inherent risks that both critical illness and its associated interventions present to the developing brain.

**Conflict of Interest Disclosures (including financial disclosures)**

The authors declare no conflicts of interest. We confirm that this manuscript is original, has not been published before, and is not under consideration elsewhere.

## Data Availability

All data produced in the present work are contained in the manuscript

## Acknowledgments

Dr. Cristiane Maia thanks Conselho Nacional de Desenvolvimento Científico e Tecnológico (CNPq) for recognizing her scientific merit with a research productivity scholarship. Brenda C. da Conceição and Lucas Villar P. S. Pantoja thank Fundação Amazônia de Amparo a Estudos e Pesquisa (FAPESPA, Brazil) for their PhD fellowships.

## Funding/Support

This work was supported by Conselho Nacional de Desenvolvimento Científico e Tecnológico – CNPq/ Brazil for the Researcher Productivity Grant (number 303196/2022-0 to CSFM) and Fundação Amazônia de Amparo a Estudos e Pesquisas-FAPESPA (Pará State Government). This study was financed in part by the Coordenação de Aperfeiçoamento de Pessoal de Nível Superior – Brasil (CAPES) – Finance Code 001.

## Contributors Statement

José Cláudio M. Rodrigues: Conceptualization, Formal analysis, Investigation, Methodology; Brenda C. da Conceição: Conceptualization, Formal analysis, Investigation, Methodology, Writing – original draft; Lucas Villar P. S. Pantoja: Conceptualization, Formal analysis, Investigation, Methodology, Writing – original draft; Kissila M. Machado-Ferraro: Formal analysis, Investigation; Mary L. F. Maia: Formal analysis, Investigation; Fábio J. C. Souza-Junior: Formal analysis, Investigation, Supervision; Rafael Rodrigues Lima: Writing – review & editing; Rodrigo A. Cunha: Writing – original draft, Writing – review & editing; Roberta Esteves Vieira de Castro: Writing – review & editing; Fernando Bezerra: Writing – review & editing; Luanna Melo P. Fernandes: Writing – review & editing; Eneas A. Fontes- Junior: Writing – review & editing; Cristiane S. F. Maia: Conceptualization, Data curation, Funding acquisition, Methodology, Project administration, Writing – original draft, Writing – review & editing. All authors approved the final manuscript as submitted and agreed to be accountable for all aspects of the work.

## Notes

### Competing Interest Statement

The authors have declared no competing interest.

### Funding Statement

This work was supported by Conselho Nacional de Desenvolvimento Cientifico e Tecnologico (CNPq/ Brazil) for the Researcher Productivity Grant (number 303196/2022-0 to CSFM) and Fundacao Amazonia de Amparo a Estudos e Pesquisas (FAPESPA, Para State Government). This study was financed in part by the Coordenacao de Aperfeicoamento de Pessoal de Nivel Superior (Brasil, CAPES, Finance Code 001).

