## Supplementary information for "Under-recognized or Under-reported? A Global Bibliometric Analysis (1994-2025) of Neurological Assessment in Pediatric Intensive Care Units"

#### CONTENTS

**Table S1.** Additional information on selected articles.

| Year | Authors | Corresponding authors | Article Title | Country | University of the corresponding author | DOI Link |
|---|---|---|---|---|---|---|
| 2025 | Horvat, CM; Barda, AJ; Claudio, EP; Au, AK; Bauman, A; Li, QY; Li, RT; Munjal, N; Wainwright, MS; Boonchalermvichien, T; Hochheiser, H; Clark, RSB | Christopher Horvat | Interoperable Models for Identifying Critically Ill Children at Risk of Neurologic Morbidity | USA | UPMC Children's Hospital of Pittsburgh | http://dx.doi.org/10.1001/jamanetworkopen.2024.57469 |
| 2024 | Silverman, A; Sasaki, M; Lima, JEE; Cheronis, C; Lin, GL; Johnson, A; Dahmoush, H; Archer, E; Grekov, K; Larocca, TJ; Van Haren, K | Andrew Silverman | Child Neurology: Remarkable Recovery From Severe Acute Necrotizing Encephalopathy | USA | Stanford Medicine, California | http://dx.doi.org/10.1212/WNL.0000000000209877 |
| 2024 | Liu, R; Majumdar, T; Gardner, MM; Burnett, R; Graham, K; Beaulieu, F; Sutton, RM; Nadkarni, VM; Berg, RA; Morgan, RW; Topjian, AA; Kirschen, MP | Raymond Liu | Association of Postarrest Hypotension Burden With Unfavorable Neurologic Outcome After Pediatric Cardiac Arrest | USA | Perelman School of Medicine at the University of Pennsylvania | http://dx.doi.org/10.1097/CCM.0000000000006339 |
| 2024 | Nishisaki, A; Reeder, RW; McGovern, EL; Ahmed, T; Bell, MJ; Bishop, R; Bochkoris, M; Burns, C; Carcillo, JA; Carpenter, TC; Diddle, W; Federman, M; Fink, EL; Franzon, D; Frazier, AH; Friess, SH; Graham, K; Hall, M; Hehir, DA; Horvat, CM; Huard, LL; Maa, T; Manga, A; McQuillen, P; Meert, KL; Morgan, RW; Mourani, PM; Nadkarni, VM; Naim, MY; Notterman, D; Palmer, CA; Sapru, A; Schneiter, C; | Akira Nishisaki | Brief report: incidence and outcomes of pediatric tracheal intubation-associated cardiac arrests in the ICU-RESUS clinical trial | USA | The Children's Hospital of Philadelphia | http://dx.doi.org/10.1186/s13054-024-05065-0 |

#### CONTENTS

**Table S1.** Additional information on selected articles.

| | | | | | | |
|---|---|---|---|---|---|---|
| | Sharron, MP; Srivastava, N; Viteri, S; Wessel, D; Wolfe, HA; Yates, AR; Zuppa, AF; Sutton, RM; Berg, RA | | | | | |
| 2024 | Sen, K; Harrar, D; Pariseau, N; Tucker, K; Keenan, J; Zhang, AQ; Gropman, A | Kuntal Sen | Seizure Characteristics and EEG Features in Intoxication Type and Energy Deficiency Neurometabolic Disorders in the Pediatric Intensive Care Unit: Single-Center Experience Over 10 Years | USA | School of Medicine and Health Sciences, Children's National Hospital | http://dx.doi.org/10.1007/s12028-024-02073-4 |
| 2024 | Tanna, R; Amorim, E; Caffarelli, M | Mauro Caffarelli | A Bedside Screening Tool for Acute Intracranial Hemorrhages in Intubated Children using Continuous Quantitative Electroencephalography Monitoring | USA | School of Medicine and Health Sciences, Children's National Hospital | http://dx.doi.org/10.1055/s-0044-1788052 |
| 2024 | Mazzio, EL; Topjian, AA; Reeder, RW; Sutton, RM; Morgan, RW; Berg, RA; Nadkarni, VM; Wolfe, HA; Graham, K; Naim, MY; Friess, SH; Abend, NS; Press, CA | Emma L. Mazzio | Association of EEG characteristics with outcomes following pediatric ICU cardiac arrest: A secondary analysis of the ICU-RESUScitation trial | USA | The Children's Hospital of Philadelphia, Civic Center Blvd | http://dx.doi.org/10.1016/j.resuscitation.2024.110271 |
| 2024 | Ari, HF; Anik, A; Demir, S; Çelik, SF | Hatice Feray Ari | Severe myxedema coma and pericardial effusion in a child with Down syndrome: the importance of adherence to levothyroxine therapy | Turkey | Aydın Adnan Menderes University | http://dx.doi.org/10.24953/turkjpediatr.2024.4587 |

### CONTENTS

**Table S1.** Additional information on selected articles.

| | | | | | | |
|---|---|---|---|---|---|---|
| 2024 | Calero, PG; García, ML; Sastre, AS; Sharluyan, A; Vicente, JCD | Paula Greciano Calero | Home alkaline solution poisoning - Do we know the risks of oral rehydration solutions? | Spain | Hospital Universitario Son Espases | http://dx.doi.org/10.3306/AJHS.2024.39.03.154 |
| 2024 | Sahin, S; Botan, E; Gün, E; Yüksel, MF; Süt, NY; Kartal, AT; Gurbanov, A; Kahveci, F; Özen, H; Havan, M; Yildirim, M; Sahap, SK; Bektas, Ö; Teber, S; Fitoz, S; Kendirli, T | Süleyman Şahin | Correlation between early computed tomography findings and neurological outcome in pediatric traumatic brain injury patients | Turkey | Ankara University Medical School | http://dx.doi.org/10.1007/s10072-024-07511-x |
| 2024 | Mulder, HD; Helfferich, J; Kneyber, MCJ | Hilde D. Mulder | The neurological wake-up test in severe pediatric traumatic brain injury: a long term, single-center experience | Netherlands | University of Groningen | http://dx.doi.org/10.3389/fped.2024.1367337 |
| 2024 | Yock-Corrales, A; Lee, JH; Domínguez-Rojas, JA; Caporal, P; Roa, JD; Fernandez-Sarmiento, J; González-Dambrauskas, S; Zhu, YN; Abbas, Q; Kazzaz, Y; Dewi, DS; Chong, SL | Adriana Yock-Corrales | A Multicenter Study on the Clinical Characteristics and Outcomes Among Children With Moderate to Severe Abusive Head Trauma | Costa Rica | Hospital Nacional de Niños | http://dx.doi.org/10.1016/j.jpedsurg.2023.09.038 |
| 2024 | Miragaia, P; Grangeia, A; Rodrigues, E; Sousa, R; Ribeiro, A | Pedro Miragaia | Acute Encephalopathy in a 10-Year-Old Patient With Maple Syrup Urine Disease: A Challenging Diagnosis | Portugal | Centro Hospitalar Universitário de São João | http://dx.doi.org/10.7759/cureus.53043 |
| 2024 | Janas, AM; Miller, KR; Stence, NV; Wyrwa, JM; Ruzas, CM; Messer, R; Mourani, PM; Fink, EL; Maddux, AB | Anna M. Janas | Utility of Early Magnetic Resonance Imaging to Enhance Outcome Prediction in Critically Ill Children with Severe Traumatic Brain Injury | USA | University of Colorado Anschutz Medical Campus | http://dx.doi.org/10.1007/s12028-023-01898-9 |

#### CONTENTS

**Table S1.** Additional information on selected articles.

| | | | | | | |
|---|---|---|---|---|---|---|
| 2024 | Bhadani, KH; Sankar, J; Datta, SK; Tungal, S; Jat, KR; Kabra, SK; Lodha, R | Jhuma Sankar | Validation of a Clinical Tool to Predict Neurological Outcomes in Critically Ill Children-A Prospective Observational Study | India | All India Institute of Medical Sciences | http://dx.doi.org/10.1007/s12098-023-04482-3 |
| 2023 | Misirlioglu, M; Ekinci, F; Yildizdas, D; Horoz, OO; Yilmaz, HL; Incecik, F; Ozsoy, M; Yontem, A; Bilen, S; Silay, S | Merve Misirlioglu | A Retrospective Cohort Study of Traumatic Brain Injury in Children: A Single-Institution Experience and Determinants of Neurologic Outcome | Turkey | Mersin University Faculty of Medicine | http://dx.doi.org/10.2478/jccm-2023-0027 |
| 2023 | LaRovere, KL; Luchette, M; Akhondi-Asl, A; DeSouza, BJ; Tasker, RC; Mehta, NM; Geva, A | Kerri L. LaRovere, | Heart Rate Change as a Potential Digital Biomarker of Brain Death in Critically Ill Children With Acute Catastrophic Brain Injury | USA | Boston Children's Hospital and Harvard Medical School | http://dx.doi.org/10.1097/CCE.0000000000000908 |
| 2023 | Nakip, OS; Pektezel, MY; Terzi, K; Kesici, S; Bayrakci, B | Ozlem Saritas Nakip | Optic nerve sheath diameter and pulsatility index for the diagnosis and follow-up in pediatric traumatic brain injury: a prospective observational cohort study | Turkey | Hacettepe University | http://dx.doi.org/10.1007/s00381-023-05959-4 |
| 2023 | Yao, S; Chong, SL; Allen, JC; Dang, HX; Ming, MX; Chan, LCN; Gan, CS; Ji, J; Fan, LJ; Kurosawa, H; Lee, JH | Shu-Ling Chong | Early metabolic derangements and unfavorable outcomes in pediatric traumatic brain injury: a retrospective multi-center cohort study | Singapora | KK Women's and Children's Hospital | http://dx.doi.org/10.21037/tp-22-443 |

#### CONTENTS

**Table S1.** Additional information on selected articles.

| | | | | | | |
|---|---|---|---|---|---|---|
| 2023 | Ketharanathan, N; Hunfeld, MAW; de Jong, MC; van der Zanden, LJ; Spoor, JKH; Wildschut, ED; de Hoog, M; Tibboel, D; Buysse, CMP | Naomi Ketharanathan | Withdrawal of Life-Sustaining Therapies in Children with Severe Traumatic Brain Injury | Netherlands | Sophia Children's Hospital | http://dx.doi.org/10.1089/neu.2022.0321 |
| 2023 | Renner, T; Tobias, JD | J. D. Tobias | Sugammadex to reverse residual neuromuscular blockade and facilitate neurologic examination in an adolescent trauma patient in the Pediatric ICU setting | USA | The Ohio State University College of Medicine | http://dx.doi.org/10.14587/paccj.2023.14 |
| 2023 | Devi, AK; Randhawa, MS; Bansal, A; Angurana, SK; Malhi, P; Nallasamy, K; Jayashree, M | Arun Bansal | Long-Term Neurological, Behavioral, Functional, Quality of Life, and School Performance Outcomes in Children With Guillain-Barre Syndrome Admitted to PICU | USA | Institute of Medical Education and Research | http://dx.doi.org/10.1016/j.pediatrneurol.2022.11.002 |
| 2022 | Martin, S; Du Pont-Thibodeau, G; Seely, AJE; Emeriaud, G; Herry, CL; Recher, M; Lacroix, J; Ducharme-Crevier, L | Sophie Martin | Heart Rate Variability in Children with Moderate and Severe Traumatic Brain Injury: A Prospective Observational Study | Canada | University of Montreal | http://dx.doi.org/10.1055/s-0042-1759877 |
| 2022 | Agrwal, S; Pallavi; Jhamb, U; Saxena, R | Pallavi | Paroxysmal Sympathetic Hyperactivity in Neurocritical Children: A Pilot Study | India | Maulana Azad Medical College | 10.5005/jp-journals-10071-24346 |
| 2022 | Tsitsipanis, C; Miliaraki, M; Ntotsikas, K; Baldounis, D; Kokkinakis, E; Briassoulis, G; Venihaki, M; Vakis, A; Ilia, S | Marianna Miliaraki | Impact of Intracranial Hypertension on Outcome of Severe Traumatic Brain Injury Pediatric Patients: | Greece | University of Crete | http://dx.doi.org/10.3390/pediatric14030042 |

#### CONTENTS

**Table S1.** Additional information on selected articles.

| | | | | | | |
|---|---|---|---|---|---|---|
| | | | A 15-Year Single Center Experience | | | |
| 2022 | Bova, SM; Serafini, L; Capetti, P; Dallapiccola, AR; Doneda, C; Gadda, A; Lonoce, L; Vittorini, A; Mannarino, S; Veggiotti, P | Stefania Maria Bova | Neurological Involvement in Multisystem Inflammatory Syndrome in Children: Clinical, Electroencephalographic and Magnetic Resonance Imaging Peculiarities and Therapeutic Implications. An Italian Single-Center Experience | Italy | V. Buzzi Children's Hospital | http://dx.doi.org/10.3389/fped.2022.932208 |
| 2022 | Regeffe, F; Chevignard, M; Millet, A; Bellier, A; Wroblewski, I; Patural, H; Javouhey, E; Mortamet, G | Guillaume Mortamet | Factors associated with poor neurological outcome in children after abusive head trauma: A multicenter retrospective study | France | Grenoble-Alpes University Hospital | http://dx.doi.org/10.1016/j.chiabu.2022.105779 |
| 2022 | Atay, G; Yazar, H; Erdogan, S; Tugrul, HC; Iscan, H; Kutlubay, B | Gürkan Atay | Therapeutic Plasma Exchange for Treating Pediatric Neurological Diseases | Turkey | University of Health Sciences Türkiye | http://dx.doi.org/10.4274/TP.galenos.2022.83997 |
| 2022 | Beck, J; Grosjean, C; Bednarek, N; Loron, G | Gauthier Loron | Amplitude-Integrated EEG Monitoring in Pediatric Intensive Care: Prognostic Value in Meningitis before One Year of Age | France | University of Reims Champagne Ardenne | http://dx.doi.org/10.3390/children9050668 |

#### CONTENTS

**Table S1.** Additional information on selected articles.

| | | | | | | |
|---|---|---|---|---|---|---|
| 2022 | Ding, X; Liu, G; Qian, SY; Zeng, JS; Wang, Y; Chu, JP; Chen, Q; Chen, JL; Duan, YY; Jin, DQ; Huang, JT; Lu, XL; Guo, YM; Shi, XA; Huo, XM; Su, J; Cheng, YB; Yin, Y; Xin, XW; Sun, ZY; Zhao, SD; Miao, HJ; Lou, ZX; Li, J; Jiang, JH; Dong, SY | Suyun Qian | Epidemiology of Cardiopulmonary Arrest and Outcome of Resuscitation in PICU Across China: A Prospective Multicenter Cohort Study | China | Capital Medical University | http://dx.doi.org/10.3389/fped.2022.811819 |
| 2022 | Özcan, S; Gunes, MSA; Havan, M; Perk, O; Azapagasi, E; Gün, E; Botan, E; Ergun, E; Ates, U; Kahilogullari, G; Kendirli, T | Serhan Özcan | Comparison of pre-PICU and per-PICU interventions, clinical features and neurologic outcomes of motor vehicle collision trauma and other mechanisms of trauma in children | Turkey | Ankara University Faculty of Medicine | http://dx.doi.org/10.14744/tjtes.2022.86617 |
| 2022 | Hui, WF; Leung, KKY; Au, CC; Fung, CW; Cheng, FWT; Kan, E; Hon, KLE | Kam Lun Ellis Hon | Clinical Characteristics and Outcomes of Acute Childhood Encephalopathy in a Tertiary Pediatric Intensive Care Unit | China | Hong Kong Children's Hospital | http://dx.doi.org/10.1097/PEC.0000000000002571 |
| 2022 | Lang, SS; Gajjar, AA; Tucker, AM; Storm, PB; Rahman, RK; Madsen, PJ; O'Brien, A; Chiotos, K; Kilbaugh, TJ; Huh, JW | Shih-Shan Lang | Urgent Neurosurgical Interventions in the COVID-19-Positive Pediatric Population | USA | University of Pennsylvania | http://dx.doi.org/10.1016/j.wneu.2021.10.155 |
| 2022 | Caputo, D; Santarone, ME; Serino, D; Pietrafusa, N; Vigevano, F; Fusco, L | Lucia Fusco | Super-refractory status epilepticus (SRSE): A case series of 22 pediatric patients | Italy | Bambino Gesù Children’s Hospital | http://dx.doi.org/10.1016/j.ejpn.2022.01.006 |

#### CONTENTS

**Table S1.** Additional information on selected articles.

| | | | | | | |
|---|---|---|---|---|---|---|
| 2022 | Becker, AE; Teixeira, SR; Lunig, NA; Mondal, A; Fitzgerald, JC; Topjian, AA; Weiss, SL; Griffis, H; Schramm, SE; Traynor, DM; Vossough, A; Kirschen, MP | Andrew Becker | Sepsis-Related Brain MRI Abnormalities Are Associated With Mortality and Poor Neurological Outcome in Pediatric Sepsis | USA | Children's Hospital of Philadelphia | http://dx.doi.org/10.1016/j.pediatrneurol.2021.12.001 |
| 2022 | Munjal, NK; Bergman, I; Scheuer, ML; Genovese, CR; Simon, DW; Patterson, CM | Christina Patterson | Quantitative Electroencephalography (EEG) Predicting Acute Neurologic Deterioration in the Pediatric Intensive Care Unit: A Case Series | USA | UPMC Children's Hospital of Pittsburgh | http://dx.doi.org/10.1177/08830738211053908 |
| 2022 | Hess, V; Zhu, F; Miguel, J; Girard, F; Toussaint, M; Klein, O; Anxionnat, R; Wiedemann, A | Arnaud Wiedemann | Vasodilator-Stimulated Phosphoprotein to Monitor Clopidogrel Posology in a 7-Year-Old Child Stented for a Post-Traumatic Intracranial Internal Carotid Artery Aneurysm | France | University of Lorraine | http://dx.doi.org/10.1055/s-0041-1726456 |
| 2022 | Deana, C; Vetrugno, L; Stefani, F; Bassi, F; Bove, T | Cristian Deana | Transcranial Doppler in a child: A most valuable imaging modality | Italy | Azienda Sanitaria Universitaria Friuli Centrale | http://dx.doi.org/10.1177/1742271X21998059 |
| 2021 | Winkler, GA; Minns, AB; Kreshak, AA | Garret A. Winkler | Severe Perampanel Toxicity in a Pediatric Patient With Prolonged Symptoms | USA | University of California at San Diego Medical Center | http://dx.doi.org/10.1016/j.jemermed.2021.07.011 |
| 2021 | Renji, S; Solomon, R; Wankhade, D; Udani, S; Udani, V | Rekha Solomon | Profile of Children With Tuberculosis in a Pediatric Intensive Care Unit in Mumbai | India | BJ Wadia Hospital for Children | http://dx.doi.org/10.1007/s13312-021-2394-0 |

#### CONTENTS

**Table S1.** Additional information on selected articles.

| | | | | | | |
|---|---|---|---|---|---|---|
| 2021 | Lang, SS; Kumar, NK; Zhao, C; Zhang, DY; Tucker, AM; Storm, PB; Heuer, GG; Gajjar, AA; Kim, CT; Yuan, I; Sotardi, S; Kilbaugh, TJ; Huh, JW | Shih-Shan Lang | Invasive brain tissue oxygen and intracranial pressure (ICP) monitoring versus ICP-only monitoring in pediatric severe traumatic brain injury | USA | University of Pennsylvania Perelman School of Medicine | http://dx.doi.org/10.3171/2022.4.PEDS21568 |
| 2021 | Rollet-Cohen, V; Sachs, P; Léger, PL; Merchaoui, Z; Rambaud, J; Berteloot, L; Kossorotoff, M; Mortamet, G; Dauger, S; Tissieres, P; Renolleau, S; Oualha, M | Virginie Rollet-Cohen | Transcranial Doppler Use in Non-traumatic Critically Ill Children: A Multicentre Descriptive Study | France | Necker-Enfants Malades University Hospital | http://dx.doi.org/10.3389/fped.2021.609175 |
| 2021 | Al-Doory, SA; Magzoub, A; Pawar, N; Radaideh, M; Saleh, SM; Al Sabbah, MA; Mir, F | Sura Ahmed Al-Doory | Complicated Pneumococcal Meningitis: A Diagnostic and Therapeutic Challenge | United Arab Emirates | Latifa Women and Children Hospital | http://dx.doi.org/10.1159/000516806 |
| 2021 | Annamalai, MR; Bhalala, U | U Bhalala | Multiple extubation failures following a rhino-enteroviral infection: A unique case report in a pediatric patient | USA | Children's Hospital of San Antonio | http://dx.doi.org/10.4103/jpgm.JPGM_883_20 |
| 2021 | Ballestín, AS; Vicente, JCD; Juan, GF; Petrosyan, AS; Ferragut, CMR; Calvar, AG; Rubio, MDC; de la Ballina, AF | Alberto Salas Ballestín | Prognostic Factors of Children Admitted to a Pediatric Intensive Care Unit After an Episode of Drowning | Spain | Son Espases University Hospital | http://dx.doi.org/10.1097/PEC.0000000000001554 |
| 2021 | Appavu, B; Burrows, BT; Nickoles, T; Boerwinkle, V; Willyerd, A; Gunnala, V; Mangum, T; Marku, I; Adelson, PD | Brian Appavu | Implementation of Multimodality Neurologic Monitoring Reporting in Pediatric Traumatic Brain Injury Management | USA | University Arizona College of Medicine | http://dx.doi.org/10.1007/s12028-021-01190-8 |

**CONTENTS**

**Table S1.** Additional information on selected articles.

| | | | | | | |
|---|---|---|---|---|---|---|
| 2021 | Madurski, C; Jarvis, JM; Beers, SR; Houtrow, AJ; Wagner, AK; Fabio, A; Wang, CY; Smith, CM; Doughty, L; Janesko-Feldman, K; Rubin, P; Pollon, D; Treble-Barna, A; Kochanek, PM; Fink, EL | Ericka L. Fink | Serum Biomarkers of Regeneration and Plasticity are Associated with Functional Outcome in Pediatric Neurocritical Illness: An Exploratory Study | USA | University of Pittsburgh School of Medicine | http://dx.doi.org/10.1007/s12028-021-01199-z |
| 2021 | Maruyama, A; Tokumoto, S; Yamaguchi, H; Ishida, Y; Tanaka, T; Tomioka, K; Nishiyama, M; Fujita, K; Toyoshima, D; Nagase, H | Hiroaki Nagase | Early non-convulsive seizures are associated with the development of acute encephalopathy with biphasic seizures and late reduced diffusion | Japan | Kobe University Graduate School of Medicine | http://dx.doi.org/10.1016/j.brainde v.2020.11.012 |
| 2021 | Woods, KS; Horvat, CM; Kantawala, S; Simon, DW; Rakkar, J; Kochanek, PM; Clark, RSB; Au, AK | Alicia K. Au | Intracranial and Cerebral Perfusion Pressure Thresholds Associated With Inhospital Mortality Across Pediatric Neurocritical Care* | USA | University of Pittsburgh School of Medicine | http://dx.doi.org/10.1097/PCC.0000000000002618 |
| 2021 | McGetrick, ME; Schneider, N; Olson, DM; Aiyagari, V; Miles, D | Molly E. McGetrick | Automated Infrared Pupillometer Use in Assessing the Neurological Status in Pediatric Neurocritical Care Patients: Case Reports and Literature Review | USA | The University of Texas Southwestern Medical Center | http://dx.doi.org/10.1055/s-0041-1731074 |
| 2021 | Hunfeld, M; Nadkarni, VM; Topjian, A; Harpman, J; Tibboel, D; van Rosmalen, J; de Hoog, M; Catsman-Berrevoets, CE; Buysse, CMP | Maayke Hunfeld | Timing and Cause of Death in Children Following Return of Circulation After Out-of-Hospital Cardiac Arrest: A Single-Center | Netherlands | Sophia Children’s Hospital | http://dx.doi.org/10.1097/PCC.0000000000002577 |

#### CONTENTS

**Table S1.** Additional information on selected articles.

| | | | | | | |
|---|---|---|---|---|---|---|
| | | | Retrospective Cohort Study* | | | |
| 2021 | Joram, N; Beqiri, E; Pezzato, S; Andrea, M; Robba, C; Liet, JM; Chenouard, A; Bourgoin, P; Czosnyka, M; Léger, PL; Smielewski, P | Nicolas Joram | Continuous Monitoring of Cerebral Autoregulation in Children Supported by Extracorporeal Membrane Oxygenation: A Pilot Study | France | University Paris | http://dx.doi.org/10.1007/s12028-020-01111-1 |
| 2021 | Turel, O; Abdillah, FK; Yozgat, CY; Uzuner, S; Duramaz, BB; Dundar, TT; Seyithanoglu, MH; Yesilbas, O; Kutlu, NO | Can Yilmaz Yozgat | A Rare Presentation of Neurobrucellosis in a 6-Year-Old Pediatric Patient with Sagittal Sinus Thrombosis | Turkey | Bezmialem Vakif University | http://dx.doi.org/10.1055/s-0040-1715482 |
| 2021 | Biggs, A; Lovett, M; Moore-Clingenpeel, M; O'Brien, N | Austin Biggs | Optic nerve sheath diameter does not correlate with intracranial pressure in pediatric neurocritical care patients | USA | Medical University of South Carolina | http://dx.doi.org/10.1007/s00381-020-04910-1 |
| 2020 | Lang, SS; Valeri, A; Zhang, BQ; Storm, PB; Heuer, GG; Leavesley, L; Bellah, R; Kim, CT; Griffis, H; Kilbaugh, TJ; Huh, JW | Shih-Shan Lang | Head of bed elevation in pediatric patients with severe traumatic brain injury | USA | Children’s Hospital of Philadelphia | http://dx.doi.org/10.3171/2020.4.PEDS20102 |

#### CONTENTS

**Table S1.** Additional information on selected articles.

| | | | | | | |
|---|---|---|---|---|---|---|
| 2020 | Meert, KL; Reeder, R; Maddux, AB; Banks, R; Berg, RA; Zuppa, A; Newth, CJ; Wessel, D; Pollack, MM; Hall, MW; Quasney, M; Sapru, A; Carcillo, JA; McQuillen, PS; Mourani, PM; Chima, RS; Holubkov, R; Sorenson, S; Varni, JW; McGalliard, J; Haaland, W; Whitlock, KB; Dean, JM; Zimmerman, JJ | Kathleen L. Meert | Trajectories and Risk Factors for Altered Physical and Psychosocial Health-Related Quality of Life After Pediatric Community-Acquired Septic Shock* | USA | Wayne State University | http://dx.doi.org/10.1097/PCC.0000000000002374 |
| 2020 | Tsou, PY; Garcia, AV; Yiu, A; Vaidya, DM; Bembea, MM | Melania M. Bembea | Association of Cerebral Oximetry with Outcomes after Extracorporeal Membrane Oxygenation | USA | Johns Hopkins University School of Medicine | http://dx.doi.org/10.1007/s12028-019-00892-4 |
| 2020 | Takia, L; Patra, N; Nallasamy, K; Saini, L; Suthar, R; Angurana, SK; Jayashree, M | Suresh K. Angurana | Acute Necrotizing Encephalopathy of Childhood with H1N1 Infection | India | Institute of Medical Education and Research | http://dx.doi.org/10.1055/s-0040-1705182 |
| 2020 | Samprathi, M; Agarwal, A; Jayashree, M; Bansal, A; Baranwal, A; Nallasamy, K; Angurana, SK | Muralidharan Jayashree | Better Groundwork Can Avoid Troubled Waters: A Developing Country Perspective on Drowning | India | Institute of Medical Education and Research | http://dx.doi.org/10.1093/tropej/fmz074 |
| 2020 | Kirschen, MP; Yehya, N; Graham, K; Kilbaugh, T; Berg, RA; Topjian, A; Diaz-Arrastia, R | Matthew P. Kirschen | Circulating Neurofilament Light Chain Is Associated With Survival After Pediatric Cardiac Arrest* | USA | University of Pennsylvania | http://dx.doi.org/10.1097/PCC.0000000000002294 |
| 2020 | Williams, V; Bansal, A; Jayashree, M; Ismail, J; Aggarwal, A; Gupta, SK; Singhi, S; Singhi, P; Baranwal, AK; Nallasamy, K | Arun Bansal | Decompressive craniectomy in pediatric non-traumatic intracranial hypertension: a single center experience | India | Institute of Medical Education and Research | http://dx.doi.org/10.1080/02688697.2020.1740648 |

#### CONTENTS

**Table S1.** Additional information on selected articles.

| | | | | | | |
|---|---|---|---|---|---|---|
| 2020 | Appavu, B; Foldes, S; Temkit, M; Jacobson, A; Burrows, BT; Brown, D; Boerwinkle, V; Marku, I; Adelson, PD | Brian Appavu | Intracranial Electroencephalography in Pediatric Severe Traumatic Brain Injury | USA | University Arizona College of Medicine | http://dx.doi.org/10.1097/PCC.0000000000002136 |
| 2020 | Bourgoin, P; Barrault, V; Joram, N; Visonneau, LL; Toulgoat, F; Anthoine, E; Loron, G; Chenouard, A | Pierre Bourgoin | The Prognostic Value of Early Amplitude-Integrated Electroencephalography Monitoring After Pediatric Cardiac Arrest* | France | University Hospital, Nantes | http://dx.doi.org/10.1097/PCC.0000000000002171 |
| 2020 | Amonkar, P; Revathi, N; Gavhane, J | Priyanka Amonkar | A study of critically ill children presenting with seizures regardless of seizure duration admitted in the PICU of a tertiary hospital in India | India | MGM Hospital and Medical College | http://dx.doi.org/10.1016/j.ebr.2020.100382 |
| 2019 | Piastra, M; De Luca, D; Genovese, O; Tosi, F; Caliandro, F; Zorzi, G; Massimi, L; Visconti, F; Pizza, A; Biasucci, DG; Conti, G | Alessandro Pizza | Clinical Outcomes and Prognostic Factors for Spontaneous Intracerebral Hemorrhage in Pediatric ICU: A 12-Year Experience | Italy | Catholic University Medical School | http://dx.doi.org/10.1177/0885066617726049 |
| 2019 | Bruns, N; Sanchez-Albisua, I; Weiss, C; Tschiedel, E; Dohna-Schwake, C; Felderhoff-Müser, U; Müller, H | Nora Bruns | Amplitude-Integrated EEG for Neurological Assessment and Seizure Detection in a German Pediatric Intensive Care Unit | Germany | University Hospital Essen | http://dx.doi.org/10.3389/fped.2019.00358 |
| 2019 | Hungerford, JL; O'Brien, N; Moore-Clingenpeel, M; Sribnick, EA; Sargel, C; Hall, M; Leonard, JR; Tobias, JD | Nicole O’Brien | Remifentanil for Sedation of Children With Traumatic Brain Injury | USA | Ohio State University | http://dx.doi.org/10.1177/0885066617704390 |

#### CONTENTS

**Table S1.** Additional information on selected articles.

| | | | | | | |
|---|---|---|---|---|---|---|
| 2019 | Lee, S; Zhao, XL; Davis, KA; Topjian, AA; Litt, B; Abend, NS | Nicholas S. Abend | Quantitative EEG predicts outcomes in children after cardiac arrest | USA | University of Pennsylvania | http://dx.doi.org/10.1212/WNL.0000000000007504 |
| 2019 | Lin, YJ; Liu, HY; Kuo, HC; Huang, CF; Hsu, MH; Cheng, MC; Chien, SJ; Lin, IC; Lo, MH; Sheus, JJ | Ying-Jui Lin | Left Ventricle Decompression Strategies in Pediatric Peripheral Extracorporeal Membrane Oxygenation | USA | Chang Gung University College of Medicine | http://dx.doi.org/10.6515/ACS.201905_35(3).20181125A |
| 2019 | Piantino, JA; Lin, A; Crowder, D; Williams, CN; Perez-Alday, E; Tereshchenko, LG; Newgard, CD | Juan A. Piantino | Early Heart Rate Variability and Electroencephalographic Abnormalities in Acutely Brain-Injured Children Who Progress to Brain Death | USA | Oregon Health and Science University | http://dx.doi.org/10.1097/PCC.0000000000001759 |
| 2018 | Okochi, S; Shakoor, A; Barton, S; Zenilman, AR; Street, C; Streltsova, S; Cheung, EW; Middlesworth, W; Bain, JM | Shunpei Okochi | Prevalence of Seizures in Pediatric Extracorporeal Membrane Oxygenation Patients as Measured by Continuous Electroencephalography | USA | Columbia University Herbert | http://dx.doi.org/10.1097/PCC.0000000000001730 |
| 2018 | Benedetti, GM; Silverstein, FS; Rau, SM; Lester, SG; Benedetti, MH; Shellhaas, RA | Renee A. Shellhaas | Sedation and Analgesia Influence Electroencephalography Monitoring in Pediatric Neurocritical Care | USA | University of Michigan | http://dx.doi.org/10.1016/j.pediatrneurol.2018.05.001 |
| 2018 | Lovett, ME; Shah, ZS; Moore-Clingenpeel, M; Sribnick, E; Ostendorf, A; Chung, MG; Leonard, J; O'Brien, NF | Marlina E. Lovett | Intensive care resources required to care for critically ill children with focal intracranial infections | USA | Ohio State University College of Medicine | http://dx.doi.org/10.3171/2018.4.PEDS17715 |

#### CONTENTS

**Table S1.** Additional information on selected articles.

| | | | | | | |
|---|---|---|---|---|---|---|
| 2018 | Sutton, RM; Reeder, RW; Landis, W; Meert, KL; Yates, AR; Berger, JT; Newth, CJ; Carcillo, JA; McQuillen, PS; Harrison, RE; Moler, FW; Pollack, MM; Carpenter, TC; Notterman, DA; Holubkov, R; Dean, JM; Nadkarni, VM; Berg, RA; Zuppa, AF; Graham, K; Twelves, C; Diliberto, MA; Tomanio, E; Kwok, J; Bell, MJ; Abraham, A; Sapru, A; Alkhouli, MF; Heidemann, S; Pawluszka, A; Hall, MW; Steele, L; Shanley, TP; Weber, M; Dalton, HJ; La Bell, A; Mourani, PM; Malone, K; Telford, R; Locandro, C; Coleman, W; Peterson, A; Thelen, J; Doctor, A | Robert M. Sutton | Chest compression rates and pediatric in-hospital cardiac arrest survival outcomes | USA | University of Pennsylvania | http://dx.doi.org/10.1016/j.resuscitation.2018.07.015 |
| 2018 | Hagos, FT; Empey, PE; Wang, PC; Ma, XC; Poloyac, SM; Bayir, H; Kochanek, PM; Bell, MJ; Clark, RSB | Robert S. B. Clark | Exploratory Application of Neuropharmacometabolomics in Severe Childhood Traumatic Brain Injury | USA | University of Pittsburgh | http://dx.doi.org/10.1097/CCM.0000000000003203 |
| 2018 | Fink, EL; Andre-von Arnim, AV; Kumar, R; Wilson, PT; Bacha, T; Aklilu, AT; Teklemariam, TL; Hooli, S; Tuyisenge, L; Otupiri, E; Fabio, A; Gianakas, J; Kochanek, PM; Angus, DC; Tasker, RC | Ericka L. Fink | Traumatic Brain Injury and Infectious Encephalopathy in Children From Four Resource-Limited Settings in Africa | USA | University of Pittsburgh Medical Center | http://dx.doi.org/10.1097/PCC.0000000000001554 |

#### CONTENTS

**Table S1.** Additional information on selected articles.

| | | | | | | |
|---|---|---|---|---|---|---|
| 2018 | Lovett, ME; Maa, T; Chung, MG; O'Brien, NF | Marlina E. Lovett | Cerebral blood flow velocity and autoregulation in paediatric patients following a global hypoxic-ischaemic insult | USA | Ohio State University College of Medicine | http://dx.doi.org/10.1016/j.resuscitation.2018.02.005 |
| 2018 | Berg, RA; Sutton, RM; Reeder, RW; Berger, JT; Newth, CJ; Carcillo, JA; McQuillen, PS; Meert, KL; Yates, AR; Harrison, RE; Moler, FW; Pollack, MM; Carpenter, TC; Wessel, DL; Jenkins, TL; Notterman, DA; Holubkov, R; Tamburro, RF; Dean, JM; Nadkarni, VM | Robert A. Berg | Association Between Diastolic Blood Pressure During Pediatric In-Hospital Cardiopulmonary Resuscitation and Survival | USA | University of Pennsylvania | http://dx.doi.org/10.1161/CIRCULATIONAHA.117.032270 |
| 2018 | Slovis, JC; Gupta, N; Li, NY; Kernie, SG; Miles, DK | Darryl K. Miles | Assessment of Recovery Following Pediatric Traumatic Brain Injury | USA | University of Texas Southwestern Medical Center | http://dx.doi.org/10.1097/PCC.0000000000001490 |
| 2018 | Cohn, EC; Robertson, TS; Scott, SA; Finley, AM; Huang, R; Miles, DK | Darryl K. Miles | Extubation Failure and Tracheostomy Placement in Children with Acute Neurocritical Illness | USA | University of Texas Southwestern Medical Center | http://dx.doi.org/10.1007/s12028-017-0429-0 |
| 2018 | Au, AK; Bell, MJ; Fink, EL; Aneja, RK; Kochanek, PM; Clark, RSB | Alicia K. Au | Brain-Specific Serum Biomarkers Predict Neurological Morbidity in Diagnostically Diverse Pediatric Intensive Care Unit Patients | USA | University of Pittsburgh Medical Center | http://dx.doi.org/10.1007/s12028-017-0414-7 |
| 2018 | Lovett, ME; Moore-Clingenpeel, M; Ayad, O; O'Brien, N | Marlina E. Lovett | Reduction of hyperthermia in pediatric patients with severe traumatic brain injury: a quality improvement initiative | USA | Ohio State University College of Medicine | http://dx.doi.org/10.3171/2017.8.PEDS17104 |

#### CONTENTS

**Table S1.** Additional information on selected articles.

| | | | | | | |
|---|---|---|---|---|---|---|
| 2017 | Rilinger, JF; Smith, CM; deRegnier, RAO; Goldstein, JL; Mills, MG; Reynolds, M; Backer, CL; Burrowes, DM; Mehta, P; Piantino, J; Wainwright, MS | Mark S. Wainwright | Transcranial Doppler Identification of Neurologic Injury during Pediatric Extracorporeal Membrane Oxygenation Therapy | USA | University Feinberg School of Medicine | http://dx.doi.org/10.1016/j.jstrokecerebrovasdis.2017.05.022 |
| 2017 | Sansevere, AJ; Duncan, ED; Libenson, MH; Loddenkemper, T; Pearl, PL; Tasker, RC | Arnold J. Sansevere | Continuous EEG in Pediatric Critical Care: Yield and Efficiency of Seizure Detection | USA | Boston Children's Hospital | http://dx.doi.org/10.1097/WNP.0000000000000379 |
| 2017 | Sandquist, MK; Clee, MS; Patel, SK; Howard, KA; Yunger, T; Nagaraj, UD; Jones, BV; Fei, L; Vadivelu, S; Wong, HR | Mary Sandquist | High Frequency of Neuroimaging Abnormalities Among Pediatric Patients With Sepsis Who Undergo Neuroimaging | USA | University of Louisville | http://dx.doi.org/10.1097/PCC.0000000000001173 |
| 2017 | Forrest, A; Butt, WW; Namachivayam, SP | Siva P Namachivayam | Outcomes of children admitted to intensive care after out-of-hospital cardiac arrest in Victoria, Australia | Australia | University of Melbourne | |
| 2017 | Ducharme-Crevier, L; Press, CA; Kurz, JE; Mills, MG; Goldstein, JL; Wainwright, MS | Mark S. Wainwright | Early Presence of Sleep Spindles on Electroencephalography Is Associated With Good Outcome After Pediatric Cardiac Arrest | USA | Northwestern University Feinberg School of Medicine | http://dx.doi.org/10.1097/PCC.0000000000001137 |
| 2016 | Ducharme-Crevier, L; Mills, MG; Mehta, PM; Smith, CM; Wainwright, MS | Mark S. Wainwright | Use of Transcranial Doppler for Management of Central Nervous System Infections in Critically Ill Children | USA | Northwestern University Feinberg School of Medicine | http://dx.doi.org/10.1016/j.pediatrneurol.2016.08.027 |

#### CONTENTS

**Table S1.** Additional information on selected articles.

| | | | | | | |
|---|---|---|---|---|---|---|
| 2016 | Mortamet, G; Kossorotoff, M; Baptiste, A; Boddaert, N; Castelle, M; Hubert, P; Lesage, F; Renolleau, S; Oualha, M | Guillaume Mortamet | Description and Contribution of Brain Magnetic Resonance Imaging in Nontraumatic Critically Ill Children | France | University of Montreal | http://dx.doi.org/10.1177/0883073816666737 |
| 2016 | Akyildiz, B; Ülgen-Tekerek, N; Özyurt, A; Pamukçu, Ö; Narin, N | Başak Akyıldız | Cerebrovascular complication of infective endocarditis complicated with abdominal trauma | Turkey | Erciyes University Faculty of Medicine | http://dx.doi.org/10.24953/turkjped.2016.05.017 |
| 2016 | Rathia, SK; Sankar, J; Kandasamy, D; Lodha, R | Rakesh Lodha | Plasmodium vivax Malaria Presenting with Multifocal Hemorrhagic Brain Infarcts in a School-going Child | India | All India Institute of Medical Sciences | http://dx.doi.org/10.1093/tropej/fmw007 |
| 2016 | Topjian, AA; Sánchez, SM; Shults, J; Berg, RA; Dlugos, DJ; Abend, NS | Topjian et al., 2016 | Early Electroencephalographic Background Features Predict Outcomes in Children Resuscitated From Cardiac Arrest | USA | University of Pennsylvania | http://dx.doi.org/10.1097/PCC.0000000000000740 |
| 2016 | Ferguson, NM; Shein, SL; Kochanek, PM; Luther, J; Wisniewski, SR; Clark, RSB; Tyler-Kabara, EC; Adelson, PD; Bell, MJ | Michael J. Bell | Intracranial Hypertension and Cerebral Hypoperfusion in Children With Severe Traumatic Brain Injury: Thresholds and Burden in Accidental and Abusive Insults | USA | University of Pittsburgh School of Medicine | http://dx.doi.org/10.1097/PCC.0000000000000709 |
| 2016 | Welch, TP; Wallendorf, MJ; Kharasch, ED; Leonard, JR; Doctor, A; Pineda, JA | Jose A. Pineda | Fentanyl and Midazolam Are Ineffective in Reducing Episodic Intracranial Hypertension in Severe Pediatric Traumatic Brain Injury | USA | Washington University School of Medicine | http://dx.doi.org/10.1097/CCM.0000000000001558 |

#### CONTENTS

**Table S1.** Additional information on selected articles.

| | | | | | | |
|---|---|---|---|---|---|---|
| 2015 | Starling, RM; Shekdar, K; Licht, D; Nadkarni, VM; Berg, RA; Topjian, AA | Alexis A. Topjian | Early Head CT Findings Are Associated With Outcomes After Pediatric Out-of-Hospital Cardiac Arrest | USA | University of Pennsylvania Perelman School of Medicine | http://dx.doi.org/10.1097/PCC.0000000000000404 |
| 2015 | Mtaweh, H; Kochanek, PM; Carcillo, JA; Bell, MJ; Fink, EL | Mtaweh et al., 2015 | Patterns of multiorgan dysfunction after pediatric drowning | USA | University of Pittsburgh | http://dx.doi.org/10.1016/j.resuscitation.2015.02.005 |
| 2014 | Sasaki, J; Chegondi, M; Raszynski, A; Totapally, BR | Jun Sasaki | Outcome of Children With Acute Encephalitis and Refractory Status Epilepticus | USA | Miami Children's Hospital | http://dx.doi.org/10.1177/0883073813513069 |
| 2014 | Yager, PH; Clark, ME; Dapul, HR; Murphy, S; Zheng, H; Noviski, N | Natan Noviski | Reliability of Circulatory and Neurologic Examination by Telemedicine in a Pediatric Intensive Care Unit | USA | Massachusetts General Hospital | http://dx.doi.org/10.1016/j.jpeds.2014.07.002 |
| 2014 | del Castillo, J; López-Herce, J; Cañadas, S; Matamoros, M; Rodríguez-Núnez, A; Rodríguez-Calvo, A; Carrillo, A | Jesus Lopez-Herce | Cardiac arrest and resuscitation in the pediatric intensive care unit: A prospective multicenter multinational study | Spain | Gregorio Marañón University General Hospital | http://dx.doi.org/10.1016/j.resuscitation.2014.06.024 |
| 2014 | Nagase, H; Nishiyama, M; Nakagawa, T; Fujita, K; Saji, Y; Maruyama, A | Hiroaki Nagase | Midazolam Fails to Prevent Neurological Damage in Children With Convulsive Refractory Febrile Status Epilepticus | Japan | Hyogo Prefectural Kobe Children's Hospital | http://dx.doi.org/10.1016/j.pediatrneurol.2014.02.021 |
| 2014 | Payne, ET; Zhao, XY; Frndova, H; McBain, K; Sharma, R; Hutchison, JS; Hahn, CD | Cecil D. Hahn | Seizure burden is independently associated with short term outcome in critically ill children | Canada | University Avenue | http://dx.doi.org/10.1093/brain/awu042 |

**CONTENTS**

**Table S1.** Additional information on selected articles.

| | | | | | | |
|---|---|---|---|---|---|---|
| 2013 | Oualha, M; Gatterre, P; Boddaert, N; Dupic, L; De Saint Blanquat, L; Hubert, P; Lesage, F; Desguerre, I | Mehdi Oualha | Early diffusion-weighted magnetic resonance imaging in children after cardiac arrest may provide valuable prognostic information on clinical outcome | France | Paris-Descartes University | http://dx.doi.org/10.1007/s00134-013-2930-z |
| 2013 | Babbitt, CJ; Halpern, R; Liao, E; Lai, K | Christopher J. Babbitt | Hyperglycemia Is Associated With Intracranial Injury in Children Younger Than 3 Years of Age | USA | University of California at Irvine | http://dx.doi.org/10.1097/PEC.0b013e3182850409 |
| 2012 | O'Brien, NF; Mella, C | O'Brien and Mella, 2012 | Brain tissue oxygenation-guided management of diabetic ketoacidosis induced cerebral edema | USA | Ohio State University College of Medicine | http://dx.doi.org/10.1097/PCC.0b013e3182601132 |
| 2012 | Huang, SC; Wu, ET; Wang, CC; Chen, YS; Chang, CI; Chiu, IS; Ko, WJ; Wang, SS | Shoei-Shen Wang | Eleven years of experience with extracorporeal cardiopulmonary resuscitation for paediatric patients with in-hospital cardiac arrest | Taiwan | National Taiwan University Hospital | http://dx.doi.org/10.1016/j.resuscitation.2012.01.031 |
| 2012 | Carroll, TG; Dimas, VV; Raymond, TT | Tia Tortoriello Raymond | Vasopressin rescue for in-pediatric intensive care unit cardiopulmonary arrest refractory to initial epinephrine dosing: A prospective feasibility pilot trial | USA | University of Texas Southwestern Medical Center | http://dx.doi.org/10.1097/PCC.0b013e31822f1569 |
| 2012 | LaRovere, KL; Brett, MS; Tasker, RC; Strauss, KJ; Burns, JP | Kerri L. LaRovere | Head Computed Tomography Scanning During Pediatric Neurocritical Care: | USA | Children’s Hospital Boston | http://dx.doi.org/10.1007/s12028-011-9627-3 |

**CONTENTS**

**Table S1.** Additional information on selected articles.

| | | | | | | |
|---|---|---|---|---|---|---|
| | | | Diagnostic Yield and the Utility of Portable Studies | | | |
| 2012 | Bustos, R | R. Bustos | Therapeutic hypothermia after pediatric cardiac arrest | Chile | Hospital Guillermo Grant Benavente | http://dx.doi.org/10.1016/j.anpedi.2011.07.017 |
| 2012 | Abend, NS; Topjian, AA; Kessler, SK; Gutierrez-Colina, AM; Berg, RA; Nadkarni, V; Dlugos, DJ; Clancy, RR; Ichord, RN | Nicholas S. Abend | Outcome prediction by motor and pupillary responses in children treated with therapeutic hypothermia after cardiac arrest | USA | University of Pennsylvania School of Medicine | http://dx.doi.org/10.1097/PCC.0b013e3182196a7b |
| 2011 | Bembea, MM; Savage, W; Strouse, JJ; Schwartz, JM; Graham, E; Thompson, CB; Everett, A | Melania M. Bembea | Glial fibrillary acidic protein as a brain injury biomarker in children undergoing extracorporeal membrane oxygenation | USA | Johns Hopkins University | http://dx.doi.org/10.1097/PCC.0b013e3181fe3ec7 |
| 2011 | Yildizdas, D; Kendirli, T; Arslanköylü, AE; Horoz, ÖÖ; Incecik, F; Ince, E; Ciftci, E | T. Kendirli | Neurological complications of pandemic influenza (H1N1) in children | Turkey | Ankara University School of Medicine | http://dx.doi.org/10.1007/s00431-010-1352-y |
| 2010 | Cambra, FJ; Palomeque, A; Muñoz-Santanach, D; Matute, SS; Balbuena, RN; Fructuoso, GG | F.J. Cambra | Use of decompressive craniectomy in the management of refractory intracranial hypertension in paediatric patients | Spain | Esplugues de Llobregat Pediatric Intensive Care Unit | http://dx.doi.org/10.1016/j.anpedi.2010.02.011 |
| 2009 | Prodhan, P; Fiser, RT; Dyamenahalli, U; Gossett, J; Imamura, M; Jaquiss, RDB; Bhutta, AT | Parthak Prodhan | Outcomes after extracorporeal cardiopulmonary resuscitation (ECPR) following refractory pediatric cardiac arrest in the intensive care unit | USA | University of Arkansas Medical Sciences | http://dx.doi.org/10.1016/j.resuscitation.2009.07.004 |

#### CONTENTS

**Table S1.** Additional information on selected articles.

| | | | | | | |
|---|---|---|---|---|---|---|
| 2009 | Subbaswamy, A; Hsu, AA; Weinstein, S; Bell, MJ | M. J. Bell | Correlation of Cerebral Near-Infrared Spectroscopy (cNIRS) and Neurological Markers in Critically Ill Children | USA | University of Pittsburgh School of Medicine | http://dx.doi.org/10.1007/s12028-008-9122-7 |
| 2008 | da Silva, PSL; Reis, ME; Aguiar, VE | Paulo Sérgio Lucas da Silva | Value of Repeat Cranial Computed Tomography in Pediatric Patients Sustaining Moderate to Severe Traumatic Brain Injury | Brazil | Hospital for Municipal Public Servants of São Paulo | http://dx.doi.org/10.1097/TA.0b013e318156866c |
| 2008 | Chevret, L; Husson, B; Nguefack, S; Nehlig, A; Bouilleret, V | Laurent Chevret | Prolonged refractory status epilepticus with early and persistent restricted hippocampal signal MRI abnormality | France | Bicêtre Hospital | http://dx.doi.org/10.1007/s00415-008-0713-1 |
| 2007 | Català-Temprano, A; Teruel, GC; Lasaosa, FJC; Odena, MP; Julian, AN; Rico, A | Albert Català-Temprano | Intracranial pressure and cerebral perfusion pressure as risk factors in children with traumatic brain injuries | Spain | University of Barcelona | http://dx.doi.org/10.3171/ped.2007.106.6.463 |
| 2007 | Stroud, MH; Okhuysen-Cawley, R; Jaquiss, R; Berlinski, A; Fiser, RT | Michael H. Stroud | Successful use of extracorporeal membrane oxygenation in severe necrotizing pneumonia caused by Staphylocloccus aureus | USA | University of Arkansas for Medical Sciences | http://dx.doi.org/10.1097/01.PCC.0000262795.11598.56 |
| 2007 | Prins, SA; De Hoog, M; Blok, JH; Tibboel, D; Visser, GH | Dick Tibboel | Continuous noninvasive monitoring of barbiturate coma in critically ill children using the Bispectral™ index monitor | Netherlands | University Medical Center | http://dx.doi.org/10.1186/cc6138 |

#### CONTENTS

**Table S1.** Additional information on selected articles.

| | | | | | | |
|---|---|---|---|---|---|---|
| 2005 | Makoroff, KL; Cecil, LM; Care, M; Ball, WS | Kathi L. Makoroff | Elevated lactate as an early marker of brain injury in inflicted traumatic brain injury | USA | University of Cincinnati College of Medicine | http://dx.doi.org/10.1007/s00247-005-1441-7 |
| 2000 | Hertzog, JH; Dalton, HJ; Anderson, BD; Shad, AT; Gootenberg, JE; Hauser, GJ | James H. Hertzog | Prospective evaluation of propofol anesthesia in the pediatric intensive care unit for elective oncology procedures in ambulatory and hospitalized children | USA | AI duPont Hospital for Children | http://dx.doi.org/10.1542/peds.106.4.742 |
| 2000 | Natale, JE; Joseph, JG; Helfaer, MA; Shaffner, DH | JoAnne E. Natale | Early hyperthermia after traumatic brain injury in children: Risk factors, influence on length of stay, and effect on short-term neurologic status | USA | Johns Hopkins School of Medicine | http://dx.doi.org/10.1097/00003246-200007000-00071 |
| 1999 | Petersen, B; Schneider, C; Strassburg, HM; Schrod, L | Birgit Petersen | Critical illness neuropathy in pediatric intensive care patients | Germany | Christian-Albrechts-University Kiel | http://dx.doi.org/10.1016/S0887-8994(99)00076-4 |
| 1999 | Hertzog, JH; Campbell, JK; Dalton, HJ; Hauser, GJ | James H. Hertzog | Propofol anesthesia for invasive procedures in ambulatory and hospitalized children: Experience in the pediatric intensive care unit | USA | Georgetown University Medical Center | http://dx.doi.org/10.1542/peds.103.3.e30 |
| 1996 | Habib, DM; Tecklenburg, FW; Webb, SA; Anas, NG; Perkin, RM | D M Habib | Prediction of childhood drowning and near-drowning morbidity and mortality | USA | Medical University of South Carolina | http://dx.doi.org/10.1097/00006565-199608000-00005 |
| 1994 | KELSALL, AWR; ROSSRUSSELL, R; HERRICK, MJ | A W Kelsall | Reversible neurologic dysfunction following - isoflurane sedation in pediatric intensive-care | United Kingdom | Addenbrookes Hospital | http://dx.doi.org/10.1097/00003246-199406000-00024 |

#### CONTENTS

**Table S1.** Additional information on selected articles.

| | | | | | | |
|---|---|---|---|---|---|---|
| 1994 | LACROIX, J; DEAL, C; GAUTHIER, M; ROUSSEAU, E; FARRELL, CA | J Lacroix | Admissions to a pediatric intensive-care unit for status epilepticus - a 10-year experience | Canada | University of Montreal | http://dx.doi.org/10.1097/00003246-199405000-00019 |
| 1994 | BRATTON, SL; JARDINE, DS; MORRAY, JP | S L Bratton | Serial neurologic examinations after near-drowning and outcome | USA | University of Washington School of Medicine | http://dx.doi.org/10.1001/archpedi.1994.02170020053008 |
